# SNX13 and SNX14 interorganelle tethers suppress lysosome membrane damage and neurodegeneration

**DOI:** 10.64898/2026.10.22.26365257

**Authors:** Kaitlynn Gov, Vanessa B. Sanchez, Emma Reynolds, Caroline Bickerton, Teresa Jimenez, Sanghee Yun, Ipsita Chakraborty, Raunaq Deo, Daniel Smith, Thomas Roule, Blessy Paul, Sonakshi Srivastava, Amelia J. Eisch, Ophir Shalem, Xilma Ortiz-Gonzalez, Clementina Mesaros, Joseph G. Gleeson, Uner Tan, Antoine Pouzet, Caroline Nava, Brett M. Collins, William A. Prinz, Jeffrey McDonald, Jonathan Levy, Boris Keren, Alexandra Afenjar, Joan Font-Burgada, W. Mike Henne, Naiara Akizu

**Author notes:** co-first authors with the agreement that they may invert the author order on their respective CVs to reflect their equal contributions (Kaitlynn Gov, Vanessa B. Sanchez). co-corresponding authors with the agreement they may invert the author order on their respective CVs to reflect their equal contributions.

## Abstract

Lysosome membrane permeabilization (LMP) is an emerging signature of neurodegenerative diseases, yet the mechanisms that enable an effective repair are largely undefined, particularly in neurons. Here, we reveal sorting nexin (SNX) proteins SNX13 and SNX14 maintain lysosome membrane lipid composition to support efficient LMP repair and neuronal survival. We identify loss of function mutations in SNX13 in children manifesting a previously unrecognized cerebellar neurodegenerative disorder. Consistently, *Snx13* ablation in mice causes cerebellar Purkinje cell death and signatures of ataxia-like motor deficits. We show loss of SNX13 or its ataxia-linked paralog SNX14 perturbs lysosome morphology, positioning, and acidity and causes increased susceptibility to lysosome damage. Using lysosome immunoprecipitation and proteomics, we comprehensively map lysosome proteome remodeling during membrane damage and repair, finding that SNX13 or SNX14 deficiency blunts recruitment of repair machinery, including ESCRTs and PITT pathway proteins, to damaged lysosomes. Mechanistically, we determine SNX13 and SNX14 support lysosome integrity through interorganelle lipid crosstalk. We define SNX13 and SNX14 as lipid binding proteins that localize to ER-lysosome junctions, where they support lysosomal phosphoinositide levels in response to membrane damage. In line with this, loss of SNX13 or SNX14 impairs lysosome PtdIns4P signaling. Altogether, our work reveals a conserved mechanism of interorganelle lipid crosstalk by SNX13/14 family proteins that safeguards lysosome membranes, maintaining them competent for repair, and is defective in cerebellar neurodegeneration.

## Introduction

Lysosomes are dynamic digestive organelles that mediate macromolecule turnover, coordinate cellular signaling, and supply metabolites for *de novo* biosynthesis pathways^1, 2^. Their limiting membrane consists of a phospholipid bilayer that serves as a barrier to maintain the acidic lumen and segregate hydrolytic enzymes from the cellular environment. Throughout their lifetime, lysosomes undergo frequent membrane remodeling, accumulation of undegraded substrates, including membrane-reactive metabolic byproducts that continually challenge membrane integrity that can culminate in lysosome membrane permeabilization (LMP). LMP compromises lysosome function and exposes the cytosol to hydrolytic enzymes capable of activating cell death pathways^3^. Notably, LMP is emerging as a key pathogenic mechanism of neurodegeneration, underscoring the critical need for neurons to preserve, repair, and restore lysosomal membrane integrity^4–6^.

Recent studies have identified specialized pathways that suppress or repair LMP through the recruitment of specific proteins to damaged lysosome membranes, including components of the endosomal sorting complexes required for transport (ESCRT) pathway^7–9^ and the phosphoinositide-initiated membrane tethering and lipid transport (PITT) pathway^10^. Whereas ESCRT recruitment mediates membrane remodeling to seal LMP nano-pores^7, 8^, the PITT pathway, triggered by local accumulation of the phosphoinositide lipid PtdIns4P (PI4P), promotes the formation of endoplasmic reticulum (ER)-lysosome contacts that are implicated in lysosomal membrane repair^10, 11^. In line with this, ER-lysosome junctions accumulate lipid transport proteins (LTPs), supporting a model in which lipid delivery from the ER replenishes damaged lysosome membranes^12, 13^. However, what LTPs at these ER-lysosome interfaces maintain a repair-responsive lysosome membrane lipid composition, and how they mechanistically contribute to LMP repair, remains to be cataloged and dissected.

Loss of function mutations in sorting nexin 14 (*SNX14)* cause the cerebellar neurodegenerative disorder known as spinocerebellar ataxia recessive 20 (SCAR20)^14–16^. Our previous work established that SNX14 deficiency alters the ER, lysosomes, and lipid droplets and disrupts cellular lipid metabolism^15, 17–20^. SNX14, along with its paralogs, SNX13, SNX19, and SNX25, belong to the SNX-RGS subfamily of SNX proteins^21^. Consistent with their role in maintaining organelle homeostasis, numerous studies have identified SNX-RGS proteins as molecular tethers that connect the ER network to other cellular compartments^18, 22–24^. A key feature of SNX-RGS proteins is the presence of N-terminal transmembrane domains that anchor them to the ER^22^ and a Phox homology (PX) domain, that enables binding to phosphoinositide-rich endosomes or lysosomes, thus facilitating ER-endolysosomal tethering^25, 26^. Although several SNX-RGS proteins contain a regulator of G-protein signaling (RGS) domain, recent studies indicate that these domains do not exhibit GAP activity in SNX-RGS proteins, suggesting they have evolved other non-canonical functions^27–30^. The main distinction of SNX-RGS proteins is the presence of two unique domains called the PX-associated (PXA) and PXC domains localized to the N and C-termini, respectively. Structural studies combining crystallography with AlphaFold-guided modeling have predicted that PXA and PXC domains interact intramolecularly to form a large hydrophobic cavity with putative lipid binding and transport functions^31^, though these have not been tested experimentally.

Here, we identify a cohort of patients with cerebellar neurodegeneration and loss of function mutations in the SNX-RGS protein SNX13. We generate a mouse model to demonstrate that *Snx13* deletion causes neurodegeneration of cerebellar Purkinje Cells (PCs) cell-autonomously. Mechanistically, we show that SNX13 or SNX14 are required to maintain lysosome membrane integrity and sustained PtdIns4P signaling in cultured human cells and human pluripotent stem cell (hPSC)-derived cortical neurons. Through lysosome immunoprecipitation (Lyso-IP) proteomics we find loss of SNX13 or SNX14 blunts LMP-induced recruitment of ESCRT machinery to lysosomes. Finally, using the yeast homolog of SNX13/14 called NVJ3, we find that the PXA:PXC module binds phospholipids *in vitro*. In line with this, we show that yeast NVJ3 maintains lysosome membrane lipid composition through its PXA:PXC module *in vivo*. Together, our findings reveal LMP as a pathogenic mechanism underlying neurodegeneration in SNX14 and novel SNX13 deficiencies, and support a model in which these multiorganelle tethers preserve lysosomal membrane homeostasis by regulating ER–lysosome lipid crosstalk.

## Results

### Identification of *SNX13* variants in individuals with a novel cerebellar ataxia syndrome

Given the structural and architectural similarities of SNX-RGS subfamily proteins, we hypothesized that, in addition to SNX14, which is associated with spinocerebellar ataxia SCAR20, other subfamily members may be genetically linked to cerebellar neurodegenerative disease. Thus, we examined exome and genome sequencing of cerebellar ataxia cohorts worldwide and identified two siblings harboring a homozygous *SNX13* nonsense variant (*SNX13* (NM001350862.1) c.1048C>T; p.R350*) (Figure 1A-C; Patient (P)1-2)). Sanger sequencing of all family members confirmed the variant was inherited from parents (Figure S1A), who were both healthy carriers. The siblings presented clinical features closely resembling SCAR20, including ataxic gait, amyotrophy, severe to moderate intellectual disability, delayed speech and social development and coarse facies characterized by thick lips, macroglossia, and a prominent forehead (Figure 1C, S1B-C, Table S1). Brain magnetic resonance imaging (MRI) of P2 at perinatal age, showed only mild cerebellar hypoplasia (Figure 1B). However, follow-up MRIs at older ages revealed cerebellar atrophy with enlarged interfolial spaces predominantly affecting anterior lobes (Figure 1B). This age dependent cerebellar tissue loss is consistent with progressive postnatal neurodegeneration.

**Figure 1.**
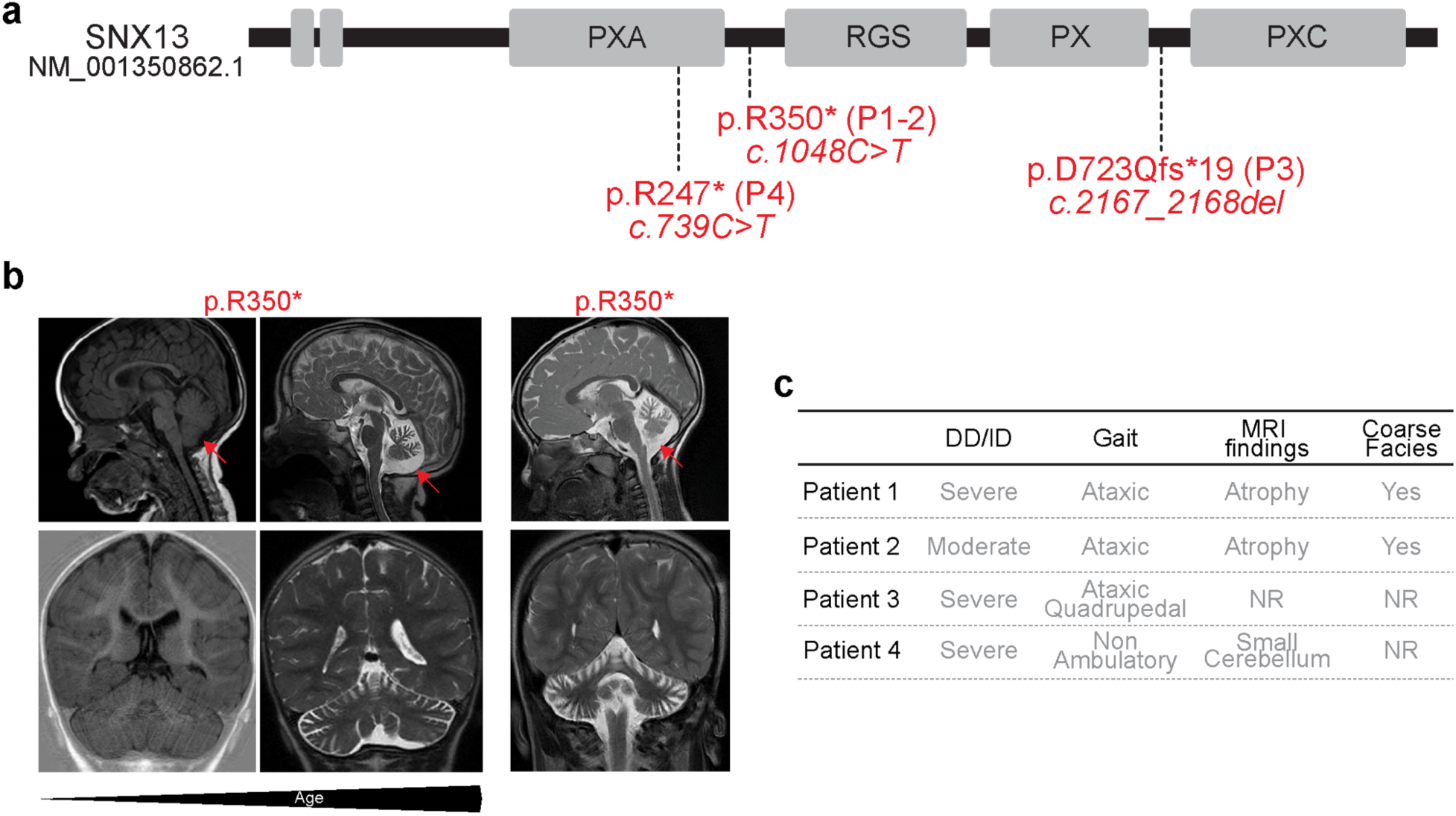
*SNX13* mutations cause a syndromic form of cerebellar atrophy with coarse facial features. **a.** Schematic representation of SNX13 protein domain architecture showing localization of homozygous truncating variants and corresponding patient (P) IDs. **b.** Midline sagittal (top) and coronal (bottom) magnetic resonance imaging (MRI) of the two siblings with homozygous SNX13 p.R350* variant at increasing ages showing progressive cerebellar atrophy (red arrow). **c.** Summary of clinical features in patients harboring homozygous loss of function variants in SNX13. DD; developmental delay; ID: intellectual disability; NR: not reported.

Subsequent analyses in independent cohorts identified two unrelated children with homozygous loss of function variants in *SNX13* (P3: c.2167_2168del; p.D723Qfs*19 and P4: c.739C>T, p.(R247*) (Figure 1A). P3 presented with developmental delay, absent speech and ataxic gait with broad-based stance, and adopted quadrupedal locomotion for rapid movement at older ages (Figure S1C, Table S1). P4 was diagnosed with developmental delay he was unable to sit or walk independently, moved by crawling and mainly communicated through sounds. P4 also showed skeletal defects, including broad and short metacarpals and phalanges and delayed bone age. Similar to P1-2, MRI of P4 revealed a small cerebellum (Table S1).

Consistent with a potential pathogenic role, these variants are either ultrarare (p.R350* allele frequency=1.25e-6) or absent from gnomAD, which contains genomic data from more than 800,000 individuals (gnomAD v4.1.1^32^). Furthermore, *SNX13* is constrained against loss-of-function variation as indicated by a pLI=1 and LOEUF=0.43 (gnomAD v4.1.1^32^). Together, these findings suggest that SNX13 loss results in an early-onset cerebellar neurodegenerative syndrome.

### *Snx13* deletion in Purkinje Cells drives cerebellar neurodegeneration

To establish a causal link between SNX13 loss and cerebellar neurodegeneration, we generated a mouse model and assessed motor behavioral and cerebellar histology. Because germline *Snx13* knockout causes embryonic lethality in mice^33^ and *Snx13* is predominantly expressed in Purkinje Cells (PCs) within the cerebellum (Figure S2A)^34^, we selectively ablated *Snx13* in PCs (*Snx13^PCKO^*) by crossing conditional *Snx13* mice with *Pcp2:Cre* transgenic mice (Figure S2B-C)^35–37^. Notably, *Snx13^PCKO^* survived to adulthood and were indistinguishable from their control littermates based on their body size (Figure 2A-B).

**Fig. 2.**
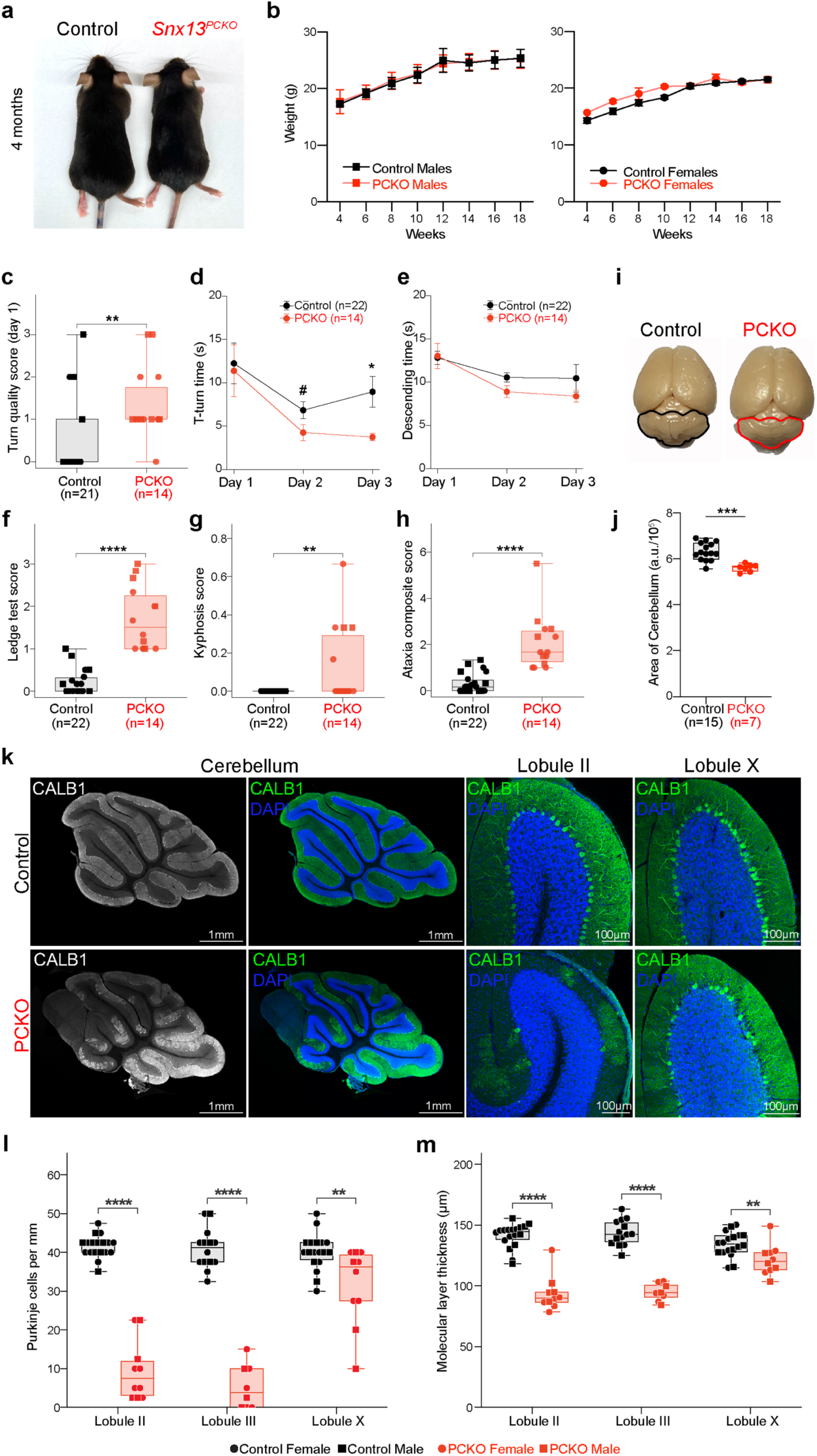
Purkinje cell-specific *Snx13* deletion causes cerebellar neurodegeneration. **a.** Photographs of 4-month-old (mo) Control and *Snx13^PCKO^* (PCKO from herein) littermates. **b.** Line plots of body weights shown as mean ±S.E.M for males (left; n=4-8 Control, n=2-4 PCKO) and females (right; n=3-7 Control, n=1-3 PCKO) across postnatal weeks 4-18. Statistical analysis was performed using two-way ANOVA followed by Sidak’s correction for multiple comparisons across eight families. **c-e.** Vertical pole test. Box plot of T-turn quality score (c) on day 1 for n=21 Control (11 females, 10 males) and n=14 PCKO (8 females, 6 males) mice. Statistical analysis was performed with two-sided Mann-Whitney U test (c). Line plots of T-turn time (d) and descending time (e), shown as mean ±S.E.M for n=22 Control (12 females, 10 males) and n=14 PCKO mice (8 females, 6 males). Statistical analysis was performed using omnibus fixed effects with Wald χ^2^ tests in linear mixed models (genotype, sex and interaction fixed; cohort random intercept; animal added for repeated measures). One control mouse did not complete the T-turn within the 2 min cutoff and was therefore excluded from the turn-quality analysis (Control n = 21 in C, n = 22 in all other panels). **f-h.** Box plots of Ledge test scores (f), Kyphosis scores (g) and Ataxia composite scores (h) shown for n=22 Control (12 females, 10 males) and n=14 PCKO mice (8 females, 6 males). Statistical analysis was performed with two-sided Mann-Whitney U test (f, g) and omnibus fixed effects with Wald χ^2^ tests in linear mixed models (genotype, sex and interaction fixed; cohort random intercept; animal added for repeated measures) (h). See Methods for details in scoring system. **i.** Photographs of mouse brains with cerebellum outlined. **j.** Box plots of cerebellar area quantifications for n=15 Control and n=7 PCKO mice. Statistical analysis was performed with two-tailed unpaired Student’s *t*-test. **k.** Representative images of sagittal cerebellar sections immunostained with PC-specific anti-CALB1 antibody (green) and DAPI for DNA (blue). Right images show a representative image of cerebellar lobules II and X. **l-m.** Box plots of PC linear density (l) and molecular layer thickness (m) in cerebellar lobules II, III and X; each symbol represents the value in one mouse (mean of two sections), n=18 Control (10 females, 8 males) and n=10 PCKO mice (6 females, 4 males) for lobules II and X; n=14 Control and n=8 PCKO mice for lobule III, which was not quantified in one cohort. Statistical analysis was performed using omnibus fixed effects with Wald χ^2^ tests in linear mixed models (genotype, lobule and their interaction fixed; cohort random intercept; animal added for repeated measures). If not otherwise specified, box plots show the median and interquartile range with whiskers from minimum to maximum; each symbol is one mouse (black, control; red, PCKO; circles, females; squares, males). Non significant values are not shown. # = near significant, * p<0.05, ** p<0.01, *** p<0.001, and **** p<0.0001. Scale bar: 100 μm and 1 mm. Additional statistics are available in Table S2.

Cerebellar neurodegeneration in mice often manifests as kyphosis (hunched back) and impaired balance and coordination even before neuronal loss is detectable^38, 39^. Thus, we assessed *Snx13^PCKO^*mice for the degree of kyphosis and performance on three motor tasks. We tested mice at 4 months of age to allow time for *Snx13* deletion after *Pcp2* promoter-driven Cre expression, which occurs around postnatal day 7^35^, and included sex as a factor in our analyses. When challenged to maintain balance on the accelerating rotarod, a rotating rod that gradually increases in speed, *Snx13^PCKO^* mice showed a tendency to fall sooner than controls, while both genotypes improved across trials indicating preserved motor learning (Fig S2D-F). Although differences between genotypes were not statistically significant, performance and rate of improvement across trials differed by sex (Table S2). In the vertical pole test, mice were placed head-up at the top of a vertical pole, and we recorded the time to turn downward (T-turn) and to descend (Figure 2C-E, S2G-H). We also scored the quality of the turn of the first trial, before mice adapted to the task, on a scale of 0 to 3 (0, smooth complete turn; 1, clumsy or hesitant turn; 2, slip during the turn; 3, fall or no turn). The clearest deficit was in the quality of the T-turn where most *Snx13^PCKO^* mice turned clumsily, often slipping or falling, whereas most controls turned smoothly and spent more time exploring the top of the pole (Figure 2C). Furthermore, *Snx13^PCKO^*mice turned faster than controls on day 3 (Figure 2D, S2G), but their poorer turn quality complicates interpretation of this shorter time. Slipping during the turn also reduced the remaining descent distance, potentially masking deficits in descent time (Figure 2E, S2H). In the ledge test, all *Snx13^PCKO^* mice scored between 1 and 3, reflecting impaired balance and coordination while walking on a narrow cage rim, whereas most controls scored 0 and none exceeded 1 (Figure 2F). This difference was significant in both sexes. *Snx13^PCKO^* mice also had higher kyphosis scores overall, with hunched posture observed in 5 of 14 mutants and no controls (Figure 2G). These deficits were major contributors to the significantly higher ataxia composite score in *Snx13^PCKO^* mice (Figure 2H), although the magnitude of the effect differed between sexes (Table S2). Together these data are consistent with cerebellar dysfunction in *Snx13^PCKO^* mice.

To determine whether ataxic phenotypes were associated with loss of cerebellar neurons, we conducted a series of anatomic and histologic analyses. While brain weights were comparable between genotypes (Figure S2I), the cerebellar area, both absolute and relative to whole-brain area, was smaller in *Snx13^PCKO^* mice than in control littermates (Figure 2I-J and S2J-K). Immunostainings for the PC marker Calbindin 1 (CALB1) revealed patches of PC loss and reduced molecular layer (ML) thickness in *Snx13*^PCKO^ cerebella, whereas control mice showed uniformly aligned soma with dendrites extending into the ML (Figure 2K-M). Regions of PC loss also showed increased GFAP and CD68 staining, consistent with an accompanying glial response (Figure S2L-M). These changes were most pronounced in the anterior cerebellar lobules II and III, where PC density was reduced to 25% and 16% of control levels, respectively, whereas lobule X was relatively spared (74% of control) (Figure 2K-M and S2L-M), resembling the regional pattern of cerebellar degeneration observed in P1-2 patients and many cerebellar neurodegenerative conditions^40^. Across all mice with both measures, PC density in lobule II was inversely correlated with the composite ataxia score and the ledge test score (r=-0.68 and -0.83, respectively; Figure S2N-O), consistent with a contribution of PC loss to the ataxic phenotype. Together, these findings support SNX13 deficiency as a previously unrecognized cause of cerebellar degeneration and ataxia.

### Loss of SNX13 perturbs lysosome homeostasis

We previously identified enlarged lysosomes with abnormal lipid storage in SCAR20 patients-derived cells and SNX14-deficient mice cerebellar PC neurons^15, 20^. The homology between SNX13 and SNX14 proteins, and the similar clinical presentations of patients with either protein deficiency, prompted us to investigate whether loss of SNX13 causes lysosomal abnormalities comparable to those resulting from SNX14 loss. We therefore assessed several lysosomal properties following individual siRNA-mediated knockdown (KD) of *SNX13* or *SNX14* in HeLa cells, with a non-targeting siRNA serving as the reference control (denoted as Control). After confirming that *SNX13* and *SNX14* siRNAs reduced their expression (Figure S3A,B), we analyzed the distribution and intensity of LAMP2 positive lysosome puncta, which is a readout of lysosome maturity and integrity. Both *SNX13-*KD and *SNX14-*KDs exhibited reduced LAMP2 staining intensity relative to Control cells (Figure S3C,D). *SNX14*-KD additionally showed increased clustering of lysosomes near the cell nucleus, whereas the spatial distribution of LAMP2 staining appeared largely unchanged upon *SNX13-*KD (Figure S3C,E,F). To assess whether these changes were accompanied by altered lysosome acidification, we measured the response of the fluorometric pH biosensor LysoSensor Yellow/Blue DND160 in live cells. This analysis revealed that both *SNX13*-KD and *SNX14*-KD lysosomes were less acidic than Control lysosomes (Figure 3A,B).

**Fig 3.**
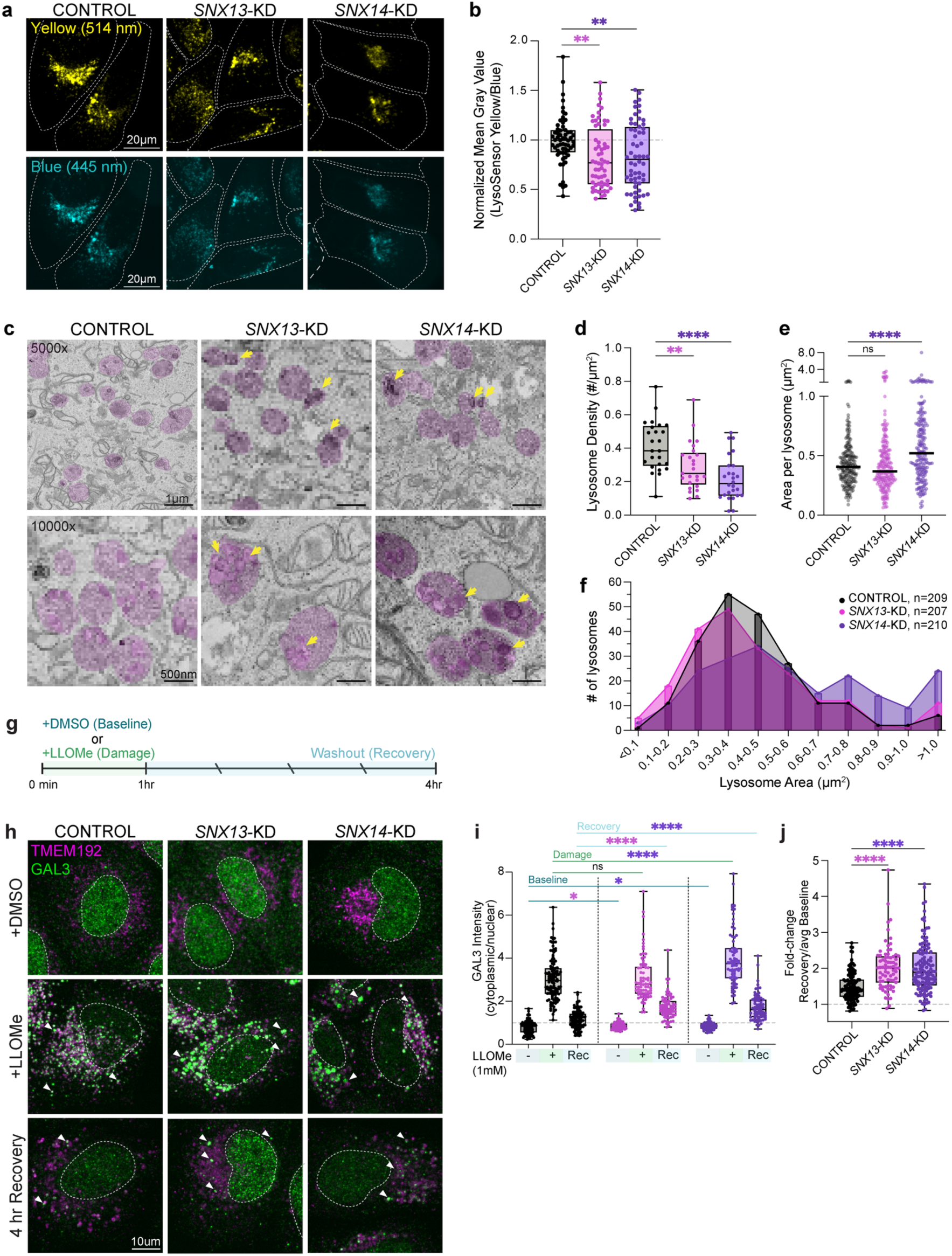
Loss of SNX13 or SNX14 perturbs lysosome acidity, morphology, and repair. **a.** Representative images of Lysosensor Yellow/Blue in live HeLa cells after siRNA knockdown. Blue signal (445nm) is higher at neutral pH and yellow signal (514nm) is higher at acidic pH. Scale bar: 20µm. **b.** Box plots of lysosomal acidity per cell, as calculated by corrected yellow/blue signal ratio, relative to average Control values. n=60 cells from three independent experiments for Control and each KD, analyzed by one-way ANOVA followed by Dunnett’s test for correction of multiple comparisons. **c.** Representative electron micrograph images of lysosomal bodies at 5000x or 10,000x, marked in pink. Yellow arrows denote large undigested membranes within KD lysosomes. 5000x scale bar: 1µm. 10,000x scale bar: 500nm. **d.** Box plots of lysosome density, calculated as the number of lysosomes per unit area from TEM images. n=25 images from at least 10 cells, analyzed by one-way ANOVA followed by Dunnett’s test for correction of multiple comparisons. **e.** Violin plots of single lysosome areas manually identified in TEM. n=207-210 lysosomes from at least 10 cells, analyzed by one-way ANOVA followed by Dunnett’s test for correction of multiple comparisons. **f.** Frequency distribution of lysosomes identified in panel E, binned by area with size set to 0.1µm2. **g.** Experimental scheme for lysosome damage and recovery monitoring with anti-GAL3. **h.** Representative images of HeLa cells with staining of lysosomes (TMEM192) and damage marker (GAL3), treated as outlined in Panel (g) after siRNA knockdown. White dotted lines were drawn around individual nuclei. Arrows denote points of TMEM192-GAL3 colocalization. Scale bar: 10µm. **i.** Box plots of cytoplasmic/nuclear GAL3 intensity per cell, over experimental time, with higher ratios consistent with lysosome damage. n=125 cells from 5 independent experiments for control and n=75 from 3 independent experiments for KDs, analyzed by one-way ANOVA followed by Dunnett’s test for correction of multiple comparisons. **j.** Box plots of fold change in cytoplasmic/nuclear GAL3 intensity per cell from baseline to 4hr recovery. n=125 cells from 5 independent experiments for Control, n=75 from 3 independent experiments for KDs. Statistical significance was determined by one-way ANOVA followed by Dunnett’s test for correction of multiple comparisons. Statistical significance is noted as: ns or not shown=not significant (p>0.05), *p<0.05, **p<0.01, and **** p<0.0001

Because reduced lysosome acidity is often associated with the accumulation of undigested material, a hallmark of lysosomal storage disorders such as Niemann-Pick Type C disease^41^, we next analyzed whether *SNX13*-KD and *SNX14*-KD lysosomes contained abnormal intraluminal structures. Using thin-section transmission electron microscopy we identified lysosomes by the presence of intraluminal vesicles and electron dense debris within the lumen (Figure 3C, highlighted in pink). Consistent with the diminished lysosome fluorescence signal observed by light microscopy, both *SNX13*-KD and *SNX14*-KD lysosomes appeared less abundant by TEM compared to Control lysosomes (Figure 3C,D). Moreover, lysosomes were morphologically distinct exhibiting less circularity (e.g. with a ruffled perimeter) and less roundness across their minor and major axes in both KDs compared with Control cells (Figure S3G-K). *SNX14-*KD lysosomes were also slightly enlarged (Figure 3E,F). Interestingly, both *SNX13*-KD and *SNX14*-KD lysosomes exhibited internal whorl-like electron-dense debris consistent with lysosome dysfunction (Figure 3C, yellow arrows, S3L-O). In all, these findings further support that acute loss of SNX13 or SNX14 directly affects lysosome function.

### Loss of SNX13 or SNX14 reduces ability to repair lysosome membranes

An intact lysosomal membrane is essential for preserving the acidic lysosome environment and degradative function^5, 42^. To determine whether compromised membrane integrity might contribute to the impaired lysosomal properties observed in *SNX13-*KD and *SNX14-*KD cells, we immunostained cells for galectin-3 (Gal3), a lectin that translocates to the cytoplasm and binds to glycans exposed upon lysosomal membrane damage^42, 43^. Results revealed that cytoplasmic Gal3 signal was modestly but significantly higher in *SNX13*-KD and *SNX14*-KD cells compared to Controls (Figure 3H,I).

Equipped with Gal3 immunostaining, we next analyzed the response of *SNX13-*KD and *SNX14-*KD cells to experimentally induced lysosome damage. We exposed cells to the lysosomotropic compound L-leucyl-L-leucine methyl ester (LLOMe), which accumulates within lysosomes and forms amyloid structures that rupture lysosome membranes^44, 45^. We then assessed Gal3 localization immediately after 1h of LLOMe treatment and 4h washout (Figure 3G). As expected, LLOMe treatment significantly increased cytoplasmic Gal3 signal in Control cells, coinciding with TMEM192 lysosome staining (Figure 3H,I). By 4h after LLOMe washout, cytoplasmic Gal3 signal returned to near-baseline levels, indicating efficient LMP repair (Figure 3H-J). In contrast, Gal3 remained significantly elevated in both *SNX13*-KD and *SNX14*-KD cytoplasm after the 4h recovery period, suggesting impaired lysosome repair (Figure 3H-J). We also calculated and compared the fold-change of cytoplasmic Gal3 signal of cells in 4h recovery over the average baseline per condition. Indeed, both KDs retained a greater relative Gal3 signal than Control cells (Figure 3J), indicating that the post-washout Gal3 difference was not explained solely by their higher baseline Gal3 signal. Overall, these data suggest loss of lysosome membrane integrity following acute SNX13 or SNX14 depletion and indicate defective repair as a mechanism of increased LMP.

### Loss of SNX13 or SNX14 mimics a lysosomal proteomic signature of membrane damage

To define how SNX13 and SNX14 loss leads to compromised lysosome membrane integrity, we analyzed the effects of acute SNX13 or SNX14 depletion on lysosomal protein composition at baseline, immediately after 1h LLOMe induced LMP, and after washout followed by a 4h recovery period. Using established methods^46, 47^, we immunoprecipitated lysosomes (Lyso-IP) with anti-HA magnetic beads from Control, *SNX13*-KD, and *SNX14*-KD HEK293T cells stably expressing the endolysosomal protein TMEM192 tagged with HA (TMEM192-RFP-3xHA) (Figures 4A, S4A,B)^44^. Anti-HA immunoprecipitates from cells expressing TMEM192-RFP-FLAG served as a mock-IP. After assessing lysosomal protein enrichment and validating the purity of the Lyso-IP purity by Western blot (Figure S4C, D), we unbiasedly analyzed the isolated fractions by liquid chromatography coupled with tandem mass spectrometry (LC-MS/MS).

**Fig. 4.**
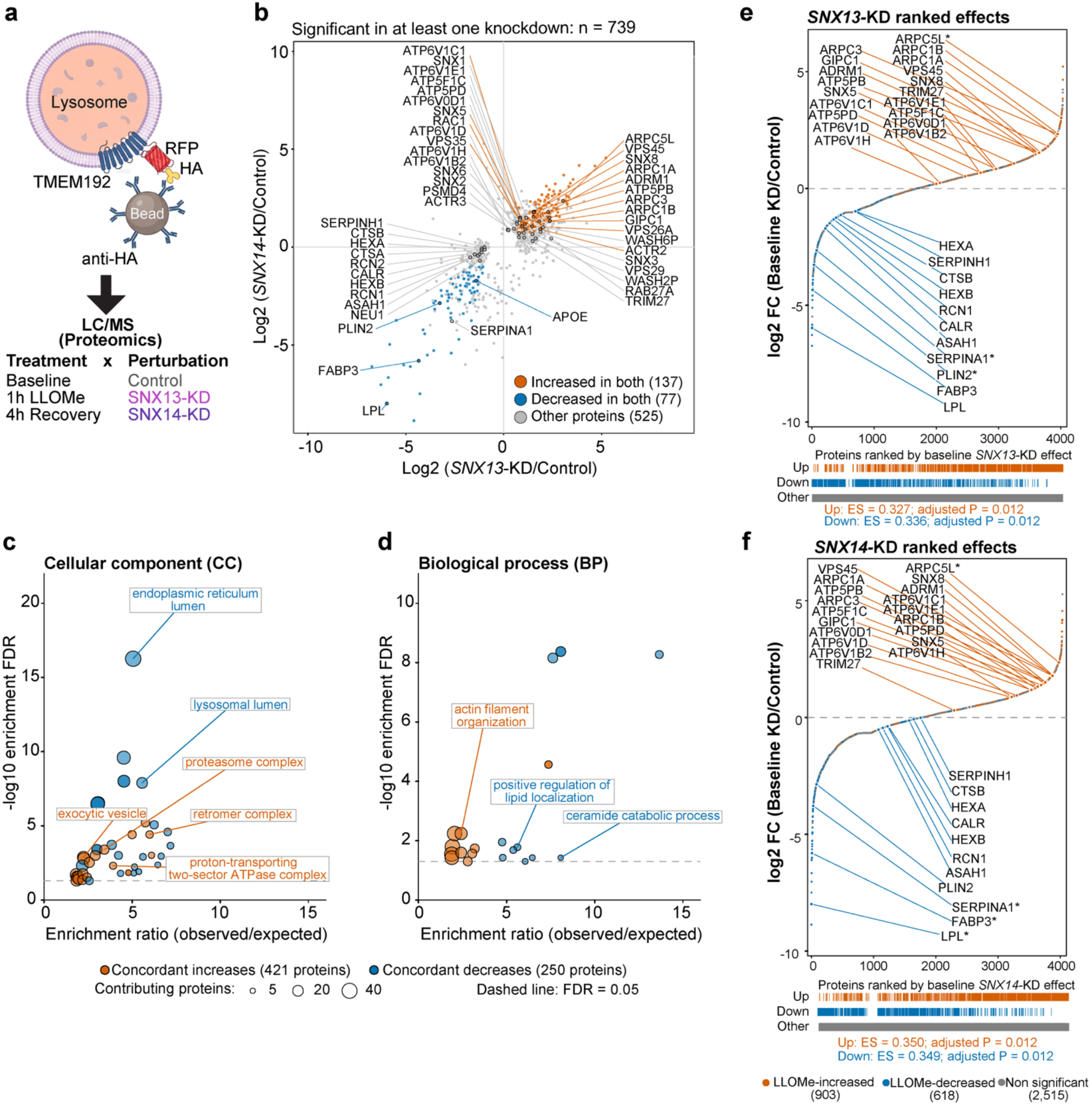
Acute SNX13 or SNX14 depletion induces a shared lysosomal proteomic signature associated with membrane damage. **a.** Experimental scheme of lysosome immunoprecipitation (lyso-IP) followed by LC/MS proteomics in HEK293T cells expressing lysosomal protein TMEM192-mRFP-3xHA under indicated treatments and perturbations. Lysosomes were isolated with anti-HA magnetic beads. Control denotes non-targeting siRNA; baseline denotes the vehicle condition. LLOMe, 1mM L-leucyl-L-leucine methyl ester; KD, knockdown. Artwork created with BioRender.com. **b.** Scatter plot showing comparison of SNX13-KD/Control and SNX14-KD/Control log2 fold changes for the 739 proteins significantly altered in at least one knockdown (FDR < 0.05) at baseline condition. Orange and blue indicate proteins significantly increased (137) or decreased (77), respectively, in both KDs; gray indicates the remaining 525 proteins. Selected proteins are labelled. **c-d.** Bubble plots showing gene ontology (GO) of cellular component (c) and biological process (d) enrichment of concordantly increased (421; orange) and decreased (250; blue) proteins, defined by changes in the same direction in both knockdowns and significance in at least one. Bubble positions indicate enrichment ratio (observed/expected) and -log10 FDR; bubble area represents the number of contributing proteins. Enrichment was assessed using hypergeometric tests with Benjamini–Hochberg correction (see Methods). Dashed lines mark FDR = 0.05. Selected terms are labelled. **e-f.** Rank-ordered plots showing all detected 4,036 proteins ranked by basal SNX13-KD/Control or SNX14-KD/control log₂ fold change. Colors indicate the response to LLOMe in control cells: increased (orange; 903 proteins), decreased (blue; 618 proteins), or not significantly altered (gray; 2,515 proteins). Tracks beneath each plot show the positions of these groups along the ranking. Selected proteins are labelled; an asterisk indicates fewer than two detected baseline replicates in control or the corresponding knockdown. Directional enrichment scores (ES) quantify enrichment of LLOMe-increased and LLOMe-decreased proteins toward the positive and negative ends of the ranking, respectively. Empirical P values were calibrated using a simulation-based null model accounting for shared controls and Holm-adjusted across the four directional tests (see Methods).

Proteomics revealed 4,036 unique proteins present across all samples, including 156 from a previously published set of 213 core lysosomal proteins^48^. To assess specificity, we compared Lyso-IP and mock-IP samples from Control cells at baseline. This comparison included 2,466 proteins detected in at least two replicates of each group, among which 113 were core lysosomal proteins (Figure S4E). To account for global differences in the amount of material recovered, we calculated a compositional enrichment score for each protein, measuring its relative abundance in Lyso-IP versus mock-IP samples after median centering. As expected, core lysosomal proteins had significantly higher scores than the other 2,353 proteins (median, 2.81 vs 0.139; Wilcoxon p=1.03×10^-30^) (Figure S4E), supporting selective recovery of the lysosomal proteome in Lyso-IP samples.

Having established specificity, we proceeded to analyze the effects of LLOMe treatment and recovery on lysosome proteomes. Principal component analysis (PCA) revealed distinct clustering by treatment and perturbation, with PC1 (34.7%) and PC2 (26%) accounting for most of the total variance (Figure S4F). Acute LLOMe treatment was associated with the dominant separation along the PC1, particularly in Control samples, whereas *SNX13-*KD and *SNX14-*KD produced a secondary, treatment-dependent shift (Figure S4F). *SNX13-*KD and *SNX14-*KD samples were also distinct from Controls at baseline, suggesting that depletion of either protein alters lysosomal proteome even in the absence of LLOMe-induced damage (Figure S4F).

To identify baseline changes in the lysosomal proteome associated with SNX13 and SNX14 depletion, we performed a differential analysis of normalized protein abundances. At a false discovery rate (FDR)<0.05, *SNX13-*KD lysosomes exhibited increased abundance of 384 proteins and decreased abundance of 218 relative to Control cells (Table S3). SNX14 depletion affected fewer proteins at the same threshold, with 212 increased and 140 decreased (Table S3). However, despite this difference, protein-level abundance changes were strongly correlated between *SNX13*-KD and *SNX14*-KD lysosomes (Pearson’s r =0.803; Figure 4B). Of the 739 proteins that changed significantly in at least one of the KDs, 137 were increased and 77 were decreased significantly in both KDs relative to Controls (Figure 4B). An additional 457 proteins changed in the same direction in both KDs but reached statistical significance in only one, further supporting broadly concordant effects of SNX13 and SNX14 depletion on the lysosome proteome (Figure 4B).

We next performed a gene ontology (GO) overrepresentation analysis of proteins that change concordantly in *SNX13-*KD and *SNX14-*KD. The 250 proteins with decreased lysosomal abundance were enriched for GO terms related to the lysosome lumen, endoplasmic reticulum lumen, lipid localization and ceramide catabolic processes (Figure 4C, D and Table S3). In contrast, the 421 proteins with increased lysosome abundance were enriched for terms related to exocytic vesicle, proton transporting ATPase complexes, proteasome complex, retromer complex and actin filament organization (Figure 4C, D and Table S3). While the depletion of such lysosomal proteins may be an expected consequence of general lysosome dysfunction, the significance of the remaining proteomic changes in this context was less readily apparent. We speculate that increases in proteasome components and V-ATPase subunits may represent a cellular response to lysosome dysfunction. In these knockdown conditions, lysosomes may upregulate alternative degradative pathways (e.g. proteasomal degradation) and increase expression of lysosome acidification machinery to try to compensate for lysosome deacidification.

To assess whether the baseline lysosomal proteomic changes in *SNX13-*KD or *SNX14-*KD cells resembled those induced by membrane damage, we compared these changes with a proteomic signature of experimentally induced LMP. We defined this signature as the changes in lysosomal protein abundance at 1h LLOMe treatment relative to baseline in Control cells. LLOMe significantly increased the lysosomal abundance of 903 proteins and decreased 618 in Control cells (Figure S4G-I Table S5). Of note, ESCRT proteins were elevated with LLOMe treatment, consistent with damage induction (Figure S4H, I and Table S6). These changes included proteins that also changed in the same direction in *SNX13*-KD and *SNX14*-KD lysosomes compared to Controls at baseline. For example, luminal digestive hexosaminidase enzymes HEXA and HEXB, and the lysosomal digestive enzyme cathepsin B (CTSB) were reduced in damaged lysosomes and in both KDs compared to Controls at baseline (Figure 4B and S4G). Other changes included less abundant ER luminal proteins such as calreticulin (CALR), reticulocalbin-1 (RCN1), and SERPINH1 (Figure 4B and S4G). In contrast, components of the lysosome ATPase proton pump were increased, including ATP6V1H and ATP6V0D1, as well as exocytic vesicle regulators such as GIPC1 (Figure 4B and S4G).

To determine how broadly this resemblance extended beyond individual proteins, we analyzed the effects of *SNX13-*KD and *SNX14-*KD on the entire LLOMe-induced lysosomal proteome signature. Compared with proteins that showed no significant response to LLOMe (2,515), the 903 proteins that increased in lysosomes upon LLOMe shifted toward higher abundance in both KDs, whereas the 618 that decreased in LLOMe shifted toward lower abundance (Figure S4J, K and Table S5). Consistent with these shifts, ranking all proteins by their abundance change upon *SNX13-*KD or *SNX14-*KD revealed enrichment of LLOMe-increased lysosomal proteins toward the positive end and LLOMe-decreased proteins toward the negative end of both rankings (Figure 4E, F). This indicates SNX13 or SNX14 loss itself may partially mimic the broad directional changes of LLOMe-induced lysosome damage. Indeed, a calibrated simulation test accounting for measurement uncertainty and shared baseline control supported all four directional enrichments (Holm-adjusted p = 0.012 for each comparison). Thus, under baseline conditions, SNX13 or SNX14 depletion reproduces, at least partially, the directional changes induced by lysosome membrane damage in the lysosome proteome.

### Loss of SNX13 or SNX14 alters damage induced lysosome proteome remodeling

Since Lyso-IP proteomics revealed signatures of lysosome dysfunction with SNX13 or SNX14 loss, we hypothesized these may be linked to alterations in the recruitment of lysosome repair machinery, such as the ESCRT and autophagosome-associated proteins that are recruited to lysosomes upon damage^8, 10^. Indeed, we detected significant elevations of ESCRT proteins including ESCRT-III subunit CHMP4B and CHMP2B upon LLOMe treatment in Control Lyso-IP samples (Figure S4G). However, these proteins were not significantly more abundant in KDs compared to Controls (Figure S5A-C). Thus, SNX13 or SNX14 depletion reproduced several features of LMP, such as the loss of lysosome luminal proteins, but lacked the accumulation of repair proteins recruited after acute LMP.

To test whether these differences could reflect an impaired engagement of damage-sensing and repair proteins in *SNX13-*KD and *SNX14-*KD lysosomes, we next analyzed how these cells responded to LLOMe-induced lysosome damage and washout. We first performed pairwise comparisons of Control and *SNX13-*KD or *SNX14-*KD Lyso-IPs at each treatment condition independently (Fig 5A-C Table S7-S9). Like at baseline conditions, lysosome protein-level abundance changes were strongly correlated between *SNX13*-KD and *SNX14*-KD cells at LLOMe and recovery (Pearson’s r=0.763 and r=0.779 respectively; Figure 5A, C). At LLOMe 1,045 proteins changed significantly in at least one knockdown compared to Controls, whereas only 366 were different at recovery (Figure 5A, C). Proteins with increased abundance in KD relative to Control lysosomes at LLOMe were enriched for proteasome components (PSMD2, PSMD8, PSMC4, PSMC1, PSMC6 and PSMA4) and protein folding chaperones (HSP90B1, HSPA1B, PDIA4, HSP90AA1, HSP90AB1), while at recovery, retromer components and associated trafficking proteins (RAB7A, VPS35, VPS26A, SNX27, VPS29 and VPS26B) were the most enriched functional associations. We speculate that these changes may reflect alterations in proteostasis pathways, which may be boosted or altered as cells adapt to lysosome dysfunction. In support of this idea, several individual proteins involved in autophagy regulation, translation and metabolism (e.g. SPG21, EEG1G, CDK4, ENO1 and DENR) remained elevated under both treatment conditions (Figure 5A-C).

**Fig. 5.**
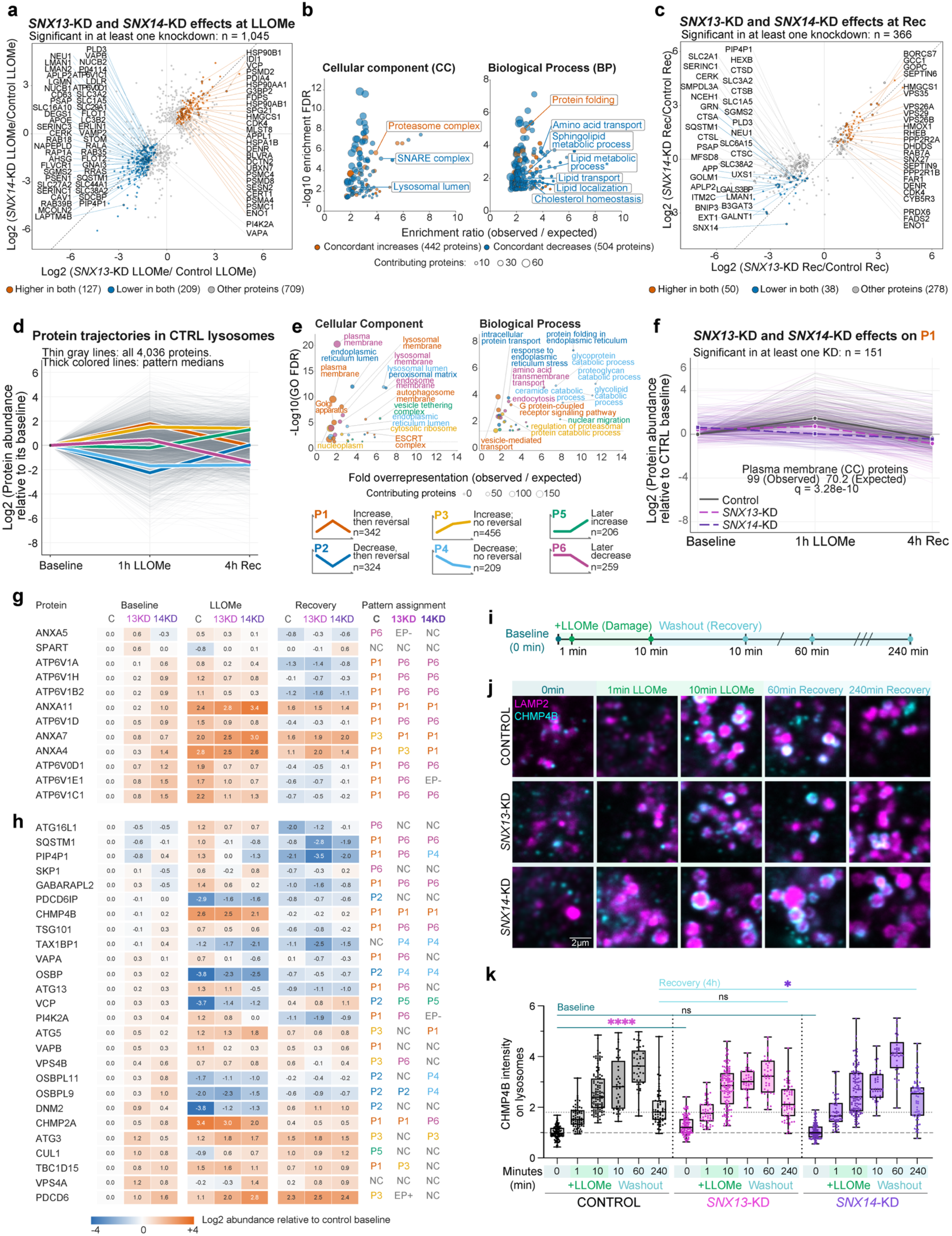
SNX13 or SNX14 depletion alters lysosomal recruitment of proteins involved in LLOMe-induced membrane damage sensing and repair. a, **c.** Scatter plot showing comparison of SNX13-KD/control and SNX14-KD/control log 2 abundance differences after 1 h LLOMe exposure (a) and 4 h after washout (c). Plots include proteins significantly altered in at least one knockdown (FDR < 0.05; 1,045 in A and 366 in C). Orange and blue indicate significantly higher or lower abundance, respectively, in both knockdowns; gray indicates the remaining proteins. The dashed diagonal marks equal effects. **b.** Bubble plots showing gene ontology (GO) of cellular component (CC) and biological process (BP) enrichment of concordantly increased (442; orange) and decreased (504; blue) proteins, defined by changes in the same direction in both knockdowns and significance in at least one. Bubble positions indicate enrichment ratio (observed/expected) and -log10 FDR; bubble area represents the number of contributing proteins. Enrichment was assessed using hypergeometric tests with Benjamini–Hochberg correction (see Methods). Selected terms are labelled. **d.** Line plots showing lysosomal protein trajectories in Control cells across baseline, 1h LLOMe and 4h after washout conditions. Thin gray lines show all 4,036 proteins relative to their baseline abundance; thick colored lines show medians of six patterns defined by significant pairwise changes (FDR < 0.05). **e.** Representative GO cellular component and biological process terms enriched among the six control patterns. Colors match D. Bubble positions indicate enrichment ratio (observed/expected) and −log₁₀ FDR; area represents the number of contributing proteins. Enrichment was assessed using hypergeometric tests with Benjamini–Hochberg correction (see Methods). P1/P2 represent acute increases/decreases followed by reversal; P3/P4 represent acute increases/decreases without detected reversal; P5/P6 represent later increases/decreases without a detected acute change. Reversal denotes an opposite-direction change after washout, without requiring a return to baseline. **f.** Line plots showing trajectories of 151 P1 proteins with a lower acute LLOMe response in both knockdowns and a significant response difference from control in at least one (FDR < 0.05). Thin lines represent individual proteins and thick lines their medians in control (gray), SNX13-KD (magenta) and SNX14-KD (purple), relative to each protein’s control baseline. Plasma-membrane proteins were overrepresented in this subset (99 observed; 70.2 expected; two-sided Fisher’s exact test with Benjamini–Hochberg correction, q = 3.28 × 10^-10^). **g, h.** Heatmaps of selected lysosomal damage sensors and sensing-associated proteins (G) and damage-response proteins (H). Values show fitted log₂ abundance differences relative to each protein’s control baseline, with colors capped at ±4. Columns within each stage show control, SNX13-KD and SNX14-KD. Pattern assignments follow D. NC indicates no statistically detected change; EP+/EP− indicates an increase/decrease between baseline and the post-washout endpoint without a resolved intermediate pattern. **i.** Experimental schematic for LLOMe treatment, washout, and fixation for CHMP4B staining. **j.** Representative images of HeLa cells with staining of lysosomes (LAMP2) and ESCRT-III component (CHMP4B) over experimental time. 60min and 240min recovery is measured after 10 minutes of 1mM LLOMe treatment. **k.** Box plots showing corrected CHMP4B intensity signal on LAMP2-positive areas over experimental time per cell, normalized to Control at 0 min (baseline) per experiment. n=40-75 cells per 2-3 independent experiments, analyzed by One-Way ANOVA followed by Dunnett’s test for correction of multiple comparisons. * p<0.05, ** p<0.01, *** p<0.001, **** p<0.0001.

Among proteins that were less abundant in KDs than in Controls at LLOMe and recovery were enzymes of the lysosome lumen (e.g. PLD3, PSAP and NEU1), amino acid transporters (e.g. SLC3A2, SLC1A5 and SLC38A2) and sphingolipid metabolism regulators (e.g. CERK, SERINC1 and SGMS2) (Figure 5A,C), indicating this loss is likely due to persistent lysosome membrane damage in *SNX13-*KD and *SNX14-*KD. At the LLOMe treatment condition, both KDs also showed reduced abundance of ER-lysosome associated tethering proteins VAPA and VAPB, which support lipid-mediated lysosomal membrane repair, as well as proteins involved in lipid handling and trafficking (SLC44A1, SLC27A2 and SERINC3) (Figure 5A). These differences with LLOMe treated Control cells could reflect impaired lipid-mediated repair of damaged lysosomes, potentially contributing to the persistent lysosome membrane damage signature in *SNX13-*KD and *SNX14-*KD.

### Loss of SNX13 or SNX14 alters recruitment trajectories of lysosome damage response proteins

The preceding analyses identified how SNX13 or SNX14 depletion altered lysosomal protein abundance at individual stages. However, these comparisons did not establish which groups of proteins followed a common course through damage and recovery, or how SNX13 or SNX14 depletion altered these temporal patterns. To resolve this, we next analyzed the lysosome recruitment trajectories of proteins upon LLOMe exposure and recovery in Control cells (Figure 5D,E). Proteins were grouped according to the direction and statistical significance of their abundance changes across the treatment conditions (FDR < 0.05). Of the analyzed proteins, 60.5% were assigned to six main trajectories, whereas 34% showed no detected abundance change. The remaining 5.5% differed between baseline and the post-washout endpoint without a resolved intermediate pattern. Patterns P1 and P2 comprised acute increases and decreases, respectively, followed by reversal after washout. Patterns P3 and P4 comprised acute increases and decreases without detected reversal. Patterns P5 and P6 comprised later increases and decreases without a detected acute change. Together, these trajectories indicate that lysosomal damage induces extensive remodeling through increases and decreases in lysosome-associated proteins. During the 4 h following washout, this remodeling was partly reversed and partly retained, while additional changes also emerged.

GO enrichment analysis linked these temporal patterns to distinct cellular components and processes (Figure 5E). For example, proteins that increased and subsequently reversed (P1) were enriched for the ESCRT complex proteins, autophagosome and lysosomal membranes, V-type ATPase complexes and vesicle-mediated transport (Figure 5E). Proteins that decreased then reversed (P2) were enriched for the endoplasmic reticulum lumen, protein folding, peroxisomal matrix and membrane-coat complexes (Figure 5E). In contrast, decreases without detected reversal (P4) were enriched for lysosomal luminal proteins and processes involved in glycoprotein, proteoglycan and lipid catabolism, whereas increases without detected reversal (P3) were associated with ribosomes, RNA processing and regulation of proteasomal protein degradation (Figure 5E). Later increases (P5) were enriched for vesicle tethering and Golgi-associated transport, while later decreases (P6) involved membrane-associated proteins, endocytosis and transmembrane transport (Figure 5E). Taken together, these findings indicate that repair-associated proteins declined after their acute enrichment, while luminal and catabolic proteins showed more sustained reductions. Additional changes in trafficking-associated proteins during the 4 h after washout suggest that lysosomes continued to reorganize as the acute damage response subsided.

We next analyzed how SNX13 or SNX14 depletion altered these temporal patterns. Because changes in lysosomal protein abundances were largely concordant between *SNX13-*KD and *SNX14-*KD across all three treatment conditions, we focused on identifying shared departures from Control-defined trajectory patterns with significant difference in at least one KD (FDR<0.05). In general, both KDs attenuated acute LLOMe-induced changes in subsets of proteins, often accompanied by baseline abundance shifts toward the LLOMe treated control state (Figure 5F, S5E). Notably, the clearest shared divergence between Controls and KDs occurred within P1, comprising proteins that transiently increased following LLOMe in Control cells. Instead, this group of proteins was slightly elevated at baseline and showed a blunted increase following LLOMe in KD lysosomes (Figure 5F). This P1 divergence pattern was shared by 151 proteins across both KDs (median Log2 acute accumulation: Control = +1.47, *SNX13-*KD = +0.37 and *SNX14-*KD = -0.54). Membrane-associated proteins were the only significantly overrepresented functional annotation within this KD departing group (O/E=1.41 q = 3.28 x 10^-10^) (Figure 5F). These included V-ATPase complex proteins (ATP6V0D1, ATP6V1C1), Ragulator components (LAMTOR1, LAMTOR2) and cytoskeletal proteins (RAC1), with direct significance in both KDs. Indeed, three ESCRT-associated proteins (CHMP2A, CHMP5 and STAM2) also showed an attenuated or reversed P1 pattern, although these changes reached significance only in *SNX14-*KD (Figure 5F-H). This supported a model where SNX13 or SNX14 loss perturbed recruitment of damage response factors to lysosomes.

The association of some of these membrane-associated proteins with lysosome quality control, and the persistent lysosome membrane damage of KD lysosomes, prompted us to perform a focused analysis of lysosome membrane sensing and response protein trajectories. Among the literature-curated list of 32 lysosome damage sensors and 75 response proteins, 22 and 51 were identified respectively in our trajectory datasets (Table S10). In Control cells, some of these proteins increased during LLOMe exposure and declined after washout (P1), others decreased and recovered (P2), and others remained increased (P3) or decreased later (P6) (Figure 5G, H). Additionally, some damage sensors showed evidence for altered patterns in KDs. For example, ANXA7, a calcium dependent phospholipid binding protein proposed to function in lysosome membrane damage sensing and repair^49^, was significantly higher in *SNX13-*KD and *SNX14-*KD lysosomes at baseline but departed from the P3 pattern in Control lysosomes, to P1 pattern in KDs (Figure 5G). SPART, a damage-associated lipid-packing sensor^50^, did not show significant differences in the trajectory pattern although baseline levels were slightly higher in both KDs (Figure 5G).

Importantly, ESCRT proteins also showed several distinct responses. The ESCRT-I subunit TSG101 retained a visible acute rise and subsequent fall (P1) in Control lysosomes (Figure 5H). Although differences did not reach significance, it changed from P1 to P6 in both KDs. PDCD6IP/ALIX decreased during 1h LLOMe and partially returned during recovery in Control cells (P2). Its acute decrease was attenuated in both KDs leading to a pattern of not detectable change (NC) (Figure 5H). CHMP2A followed a P1 pattern in Control cells but its rise in *SNX14-*KD was significantly smaller than in Controls during LLOMe treatment (Figure 5H). VPS4A/B were higher in both KDs at baseline and showed variable response during LLOMe and recovery. Whereas VPS4A remained in the same pattern in both KDs, VPS4B changed pattern assignments without significantly affecting abundance changes at LLOMe or recovery (Figure 5H). Similar to ESCRT protein changes, proteins of the PITT-mediated repair pathway also showed concordant changes in both KDs, with VAPB exhibiting the clearest shared change. Its LLOMe-induced acute rise was absent in the two KD patterns (*SNX13-*KD, −1.444; q = 0.0315 and *SNX14-*KD, −1.131; q = 0.0367) (Figure 5H). VAPA also showed a smaller acute LLOMe-induced response in both KDs than in Controls, although the difference was significant only in *SNX14-*KD (Log2FC = -1.361; q = 0.0211) (Figure 5H). In Control lysosomes OSBP, OSBPL9 and OSBPL11 were decreased followed by rebound (P2) and their pattern assignment changed in KDs (Figure 5H).

Altogether, these analyses indicate that SNX13 or SNX14 depletion alters the temporal remodeling of the lysosomal proteome, with shared attenuation of acute responses among damage-sensing and repair associated proteins. These findings, together with the altered proteomic of SNX13 and SNX14 KD lysosomes at baseline, reflects a diminished damage-sensing and repair response, further revealed by the induction of additional lysosome membrane damage with LLOMe.

### Loss of SNX13 or SNX14 alters the temporal recruitment pattern of CHMP4B to LLOMe-damaged lysosomes

LysoIP proteomics indicated altered LMP repair protein recruitment to lysosomes in SNX13 and SNX14 depleted cells. To validate this finding with orthogonal methods and evaluate repair protein recruitment to lysosomes with more temporal resolution, we treated cells with LLOMe and monitored the levels of ESCRT-III subunit CHMP4B on lysosomes. We focused on CHMP4B because, in contrast to other repair subunits that displayed blunted lysosome recruitment in *SNX13-*KD or *SNX14-*KD (Figure 5H), CHMP4B appeared be recruited to lysosome at 1h LLOMe, in line with previous studies indicating its strong recruitment to damaged lysosomes^7^. We thus reasoned that CHMP4B represented a detectable ESCRT repair factor across lysosome samples we could monitor over time to determine how SNX13 or SNX14 loss impacted repair protein dynamics. Samples received a 10min LLOMe treatment followed by a washout of 10 min, 1h, or 4h for recovery before fixation and immunostaining for CHMP4B and LAMP2 (Figure 5I). Consistent with previous reports^7^, CHMP4B was quickly recruited to Control lysosome membranes at 1 min after LLOMe exposure (Figures 5J, K, S5F). Of note, *SNX13*-KD HeLa cells exhibited significantly elevated CHMP4B lysosome signal even before LLOMe exposure (Figure 5J,K, S5F). Interestingly, although lysosomal CHMP4B intensity increased in SNX13-KD at 1min of LLOMe exposure (Figure 5J, K), this increase was lower than in Control cells (Fig S5G), suggesting an early blunting of CHMP4B recruitment following acute LLOMe damage. CHMP4B lysosome signal further increased after 10min of LLOMe, indicating that prolonged exposure causes more lysosome damage and additional CHMP4B recruitment. Notably, CHMP4B accumulation in lysosomes peaked at 1h of recovery following the 10min LLOMe pulse and washout, suggesting CHMP4B recruitment continues during the recovery phase to promote lysosome repair. Of note, CHMP4B profiles were different for both KDs compared to Control at 1h recovery, with SNX13-KD again showing blunted CHMP4B lysosome signal compared to baseline, and SNX14-KD displaying a boosted change from its baseline (Figure S5H). By 4h recovery, CHMP4B levels returned to near baseline levels in Control cells, consistent with the lysosome repair process (Figure 5J,K). However, CHMP4B levels remained higher in SNX14-KD lysosomes than in Control consistent with incomplete repair (Figure 5J, K, S5I). Collectively, this indicates that SNX13 or SNX14 loss alters CHMP4B lysosome recruitment during LLOMe-induced damage and repair, consistent with their roles in maintaining lysosome homeostasis.

### SNX13/14 homolog NVJ3 binds phospholipids in vitro

Having established that loss of SNX13 and SNX14 impacts lysosome homeostasis and recruitment trajectories of repair proteins, we next investigated the molecular mechanism. Several recent studies indicate that lipid transport proteins (LTPs) enrich at ER-lysosome contacts to promote lipid movement to lysosomes and aid LMP repair^19,21,22^. In line with this, ours and other previous work implicate SNX13 and SNX14 in interorganelle lipid crosstalk^15, 18, 24^. Notably, AlphaFold guided structural modeling suggests the PXA and PXC domains of SNX-RGS proteins, including SNX13 and SNX14, interact intramolecularly to form a conserved module, defined by a large hydrophobic cavity, ranging from ∼2000-6000 Å^31^ and similar to known hydrophobic LTP lipid binding pockets^11^. However, it is still unknown whether this putative LTP module binds or transports lipids.

To experimentally test this, we took advantage of the yeast ortholog of SNX13/14 called nucleus-vacuole junction protein 3 (NVJ3), which naturally encodes only the putative lipid binding PXA:PXC domains predicted to form the LTP module (Figures 6A and S6A). AlphaFold structural predictions indicate that this module forms a large hydrophobic cavity similar to SNX13 and SNX14, that could in principle bind to lipids (Figure 6B and S6A). Thus, NVJ3 provides a naturally existing model for studying the PXA:PXC module in vivo, while leveraging well characterized lipid metabolic pathways in yeast^11^.

**Fig. 6.**
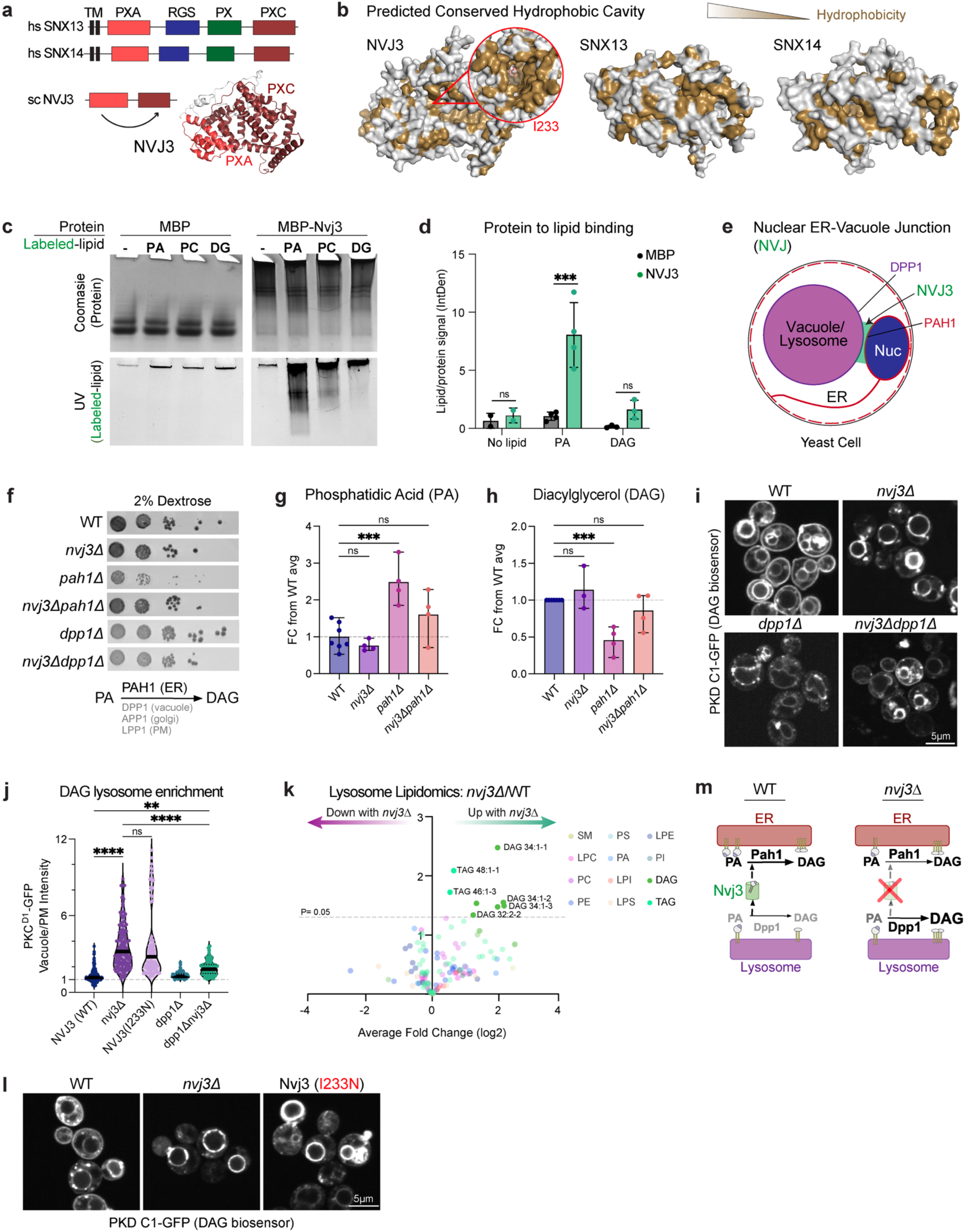
SNX13/14 ortholog NVJ3 binds PA *in vitro* and modulates PA/DAG metabolism at ER-lysosome contact sites in yeast. **a.** Domain architecture of SNX13, SNX14, and NVJ3, with predicted structure from interaction between distal PXA and PXC domains. **b.** Molecular structure of the predicted hydrophobic cavity formed by PXA:PXC interaction in NVJ3, SNX13, and SNX14. Hydrophobic regions are marked in brown. Hydrophobic core mutation I233N is enlarged and marked in red. **c.** Protein-lipid binding assay on a native gel, including MBP and purified MBP-Nvj3, binding to NBD-labeled phosphatidic acid (PA), phosphatidylcholine (PC), and diacylglycerol (DAG); protein stained with Coomassie, and lipid fluorescence detected under UV light. **d.** Bar graphs showing mean ±SD of lipid binding intensity measured as lipid fluorescent signal relative to protein Coomassie signal intensity. n=2-4 independent experiments. Statistical analysis was performed by multiple unpaired two-sided t-tests with Holm-Šidák correction. **e.** Schematic of the Nuclear ER-Vacuole Junction (NVJ) in yeast, showing localization of NVJ3 to the junction (green), Pah1 to the ER-side of the NVJ (red), and DPP1 to the vacuole/lysosome (purple). **f.** Serial dilutions of labeled yeast strains grown on synthetic dextrose media at 30C. Below, a schematic showing the conversion of PA to DAG by PA phosphatases and their respective localizations. **g, h.** Relative lipid quantification from thin layer chromatography of whole yeast cells. Bar graphs show PA levels relative to wildtype, quantified using Egg PA standard (A) and DAG levels relative to wild type. n=3-7 independent experiments analyzed by one-way ANOVA followed by Dunnett’s test for correction of multiple comparisons. **i.** Representative images of yeast transformed with PKD C1-GFP DAG biosensor in yeast with denoted gene knockouts. Scale bar: 5 µm. **j.** Violin plots of DAG biosensor enrichment on the lysosome per cell, quantified as ratio of signal on the lysosome divided by the signal on the plasma membrane. n=100-150 yeast from 3 independent experiments, analyzed by one-way ANOVA followed by Dunnett’s test for correction of multiple comparisons. **k.** Volcano plot of lipid species from isolated yeast lysosomes, showing mean fold-changes in the Nvj3-KO vs wildtype yeast in n=3 independent cultures. Lipid species are color-coded by major lipid groups, consistent with legend. The dotted line represents P<0.05, as analyzed by two-tailed t-test. **l.** Representative images of yeast transformed with PKD C1-GFP DAG biosensor with wildtype Nvj3, Nvj3-KO, and Nvj3 replaced with Nvj3 (I233N) at its endogenous locus. Scale bar: 5 µm. **m.** Model for Nvj3-mediated transport of PA from lysosome to ER, and PA metabolism at both membranes. Statistical significance is noted as: ns=not significant (p>0.05), ** p<0.01, *** p<0.001, and **** p<0.0001.

To test whether NVJ3 directly interacts with lipids *in vitro*, we generated recombinant NVJ3 protein tagged with maltose binding protein (MBP) (Figure S6B) and conducted *in vitro* lipid binding assays. After incubating MBP-NVJ3 (or MBP alone as a negative control) with different phospholipids conjugated with the fluorophore nitrobenzoxadiazole (NBD), we ran the reactions in native PAGE gels and analyzed if the fluorescent lipid signal co-migrated with the recombinant NVJ3 protein. Notably, whereas MBP did not co-migrate with any NBD-lipids we tested, MBP-NVJ3 exhibited clear co-migration with the NBD-conjugated charged lipid phosphatidic acid (PA), but not with diacylglycerol (DAG) (Figure 6C,D). This observation indicated that NVJ3 interacts with PA *in vitro*, and may be involved in some aspect of PA and/or phospholipid metabolism.

### NVJ3 maintains yeast PA and DAG pools and their cellular distribution

To further investigate the role of NVJ3 in lysosome lipid homeostasis, we conducted a large-scale candidate-based genetic interaction screen in budding yeast. We focused on genes involved in lipid metabolism, lipid transport, and inter-organelle contact, and queried whether they genetically interacted with *NVJ3*. Upon deletion of *NVJ3* alone or in combination with numerous candidate proteins, we cultured yeast on synthetic complete media containing 2% dextrose, and monitored their growth rates on solid agar media (Figure S6C). NVJ3 deletion alone mildly affected growth rate at elevated temperatures (data not shown), and other combined deletions modestly worsened this phenotype (Figure S6C). Notably, our screening revealed an interaction between NVJ3 and the PA phosphatase *PAH1* (Figure 6E, F). PAH1 is a master regulator of PA metabolism in yeast, and its deletion results in elevated PA and decreased DAG, which perturbs cell growth and division^51, 52^. Our screen revealed that deletion of *NVJ3* together with *PAH1* (*nvj3Δpah1Δ)* rescued the growth defect of *pah1*Δ yeast (Figure 6F). From this, we hypothesized *NVJ3* deletion may correct the cellular PA:DAG lipid imbalance caused by *PAH1* loss. Strikingly, biochemical lipid measurements indeed revealed deletion of *NVJ3* corrected the PA:DAG imbalance in *pah1*Δ yeast (Figure 6G, H). Furthermore, this effect was specific to PA:DAG metabolism as *NVJ3* deletion did not rescue PE or PC phospholipid imbalances detected in *pah1*Δ yeast (Figure S6E-F). This specificity suggested that NVJ3 selectively regulates PA metabolism by directly influencing PA and/or DAG pools *in vivo*.

To mechanistically dissect how NVJ3 influences PA/DAG balance in cells, we imaged yeast expressing the fluorescent DAG biosensor PKD C1-GFP^56^. As expected, wildtype yeast displayed DAG pools at both the cell surface and yeast vacuole (defined as the lysosome hereon), but *nvj3*Δ yeast exhibited significant DAG accumulation on the lysosome surface and reduced cell surface DAG signal (Figure 6I, J). To quantitatively validate lysosome DAG accumulation, we purified yeast lysosomes from wildtype and *nvj3*Δ yeast, and after confirming lysosome isolation (Figure S6H), we analyzed them by LC-MS/MS lipidomics. Consistent with imaging, lysosomes purified from *nvj3*Δ yeast showed elevations of most DAG species compared to wildtype lysosomes (Figure 6K, L Figure S6I). In conclusion, NVJ3 is required for maintaining yeast lysosome lipid composition, potentially similar to mammalian SNX13 or SNX14.

### The lipid-binding interface of NVJ3 is required for ER-Lysosome lipid crosstalk

Recent studies have shown that NVJ3, PAH1 and PA/DAG metabolism are spatially localized to the nucleus-vacuole junction (NVJ), akin to the yeast ER-lysosome interface (Figure 6E, S6G)^53–55^. Combined with our finding that NVJ3 binds PA *in vitro*, these data suggested that NVJ3 could maintain lysosome lipid homeostasis through its lipid binding activity, and potentially function as a lipid transporter between the ER network and lysosome surface. To test this model, we mutated the hydrophobic residue isoleucine-233 (I233) within the NVJ3 hydrophobic cavity to asparagine (N), predicted to disrupt lipid binding and NVJ3 function (Figure 6B). Importantly, when expressed on its endogenous promoter, the I233N-NVJ3 mutant still localized to the ER-lysosome interface (the NVJ) like wildtype NVJ3 (both tagged with the fluorescent protein mNG), indicating that the I233N mutation does not disrupt NVJ3 localization (Figure S6J). Strikingly, replacing endogenous NVJ3 with the I233N-NVJ3 mutant phenocopied the lysosome DAG accumulation observed in NVJ3 knockout yeast (Figure 6L), indicating that the hydrophobic cavity is required to maintain NVJ3 function despite normal localization.

These findings prompted us to investigate how loss of NVJ3-lipid binding activity leads to lysosomal DAG accumulation. Because recombinant NVJ3 binds PA, we hypothesized that NVJ3 promotes PA exchange between the lysosome and ER network. In this model, loss of NVJ3 would result in PA accumulation on lysosome membranes, where it could be gradually converted to DAG by the lysosome-localized PA phosphatase DPP1^57,58^ (Figure 6F,M). To test this prediction, we deleted *DPP1* in combination with *NVJ3,* and compared lysosome DAG levels between NVJ3 knockout and hydrophobic mutant I233N-NVJ3-expressing yeast. As a control, we validated that *DPP1* deletion alone did not significantly alter the DAG distribution ratio between the cell surface and vacuole. Strikingly, dual deletion of *NVJ3* and *DPP1* returned yeast lysosome/PM DAG distribution to near baseline levels (Figure 6I,J). These results demonstrate that NVJ3 lipid binding maintains lysosome PA and DAG homeostasis, potentially by facilitating PA transport across lysosome-ER contact sites (Figure 6M).

Collectively, our study suggests an evolutionarily conserved role for SNX-RGS proteins in preserving lysosome composition through non-vesicular lipid transfer, a mechanism that may underlie the role of SNX13 and SNX14 in maintaining lysosome membrane integrity and promoting repair following damage in neurons.

### Loss of SNX13 or SNX14 in human neurons perturbs lysosome homeostasis and LMP repair

Despite the recognized vulnerability of neurons to disrupted lysosome homeostasis^9, 48, 56^, how neurons safeguard lysosomal integrity remains unclear. Emerging evidence suggests that canonical LMP sensing and repair pathways are not fully conserved in neurons^57, 58^, raising the possibility of distinct protective mechanisms. To determine whether SNX13 and SNX14 contribute to these mechanisms, we generated human pluripotent stem cell (hPSC)-derived neuronal models by depleting *SNX13* or *SNX14* with CRISPRi and inducing their differentiation to neurons by transient *NGN2* overexpression^59^. We confirmed depletion of *SNX13* and *SNX14* in hPSCs and in neurons (Figure S7A-B), and validated expression of pluripotency markers and efficient differentiation to neurons (Figure S7C-E). We then stained neurons with LAMP1 and noted that *SNX14*-KDs but not *SNX13-*KDs exhibited significantly enlarged lysosomes (Figure 7A-C) consistent with our TEM observations in HeLa cells and previous reports^15, 20^.

**Fig. 7.**
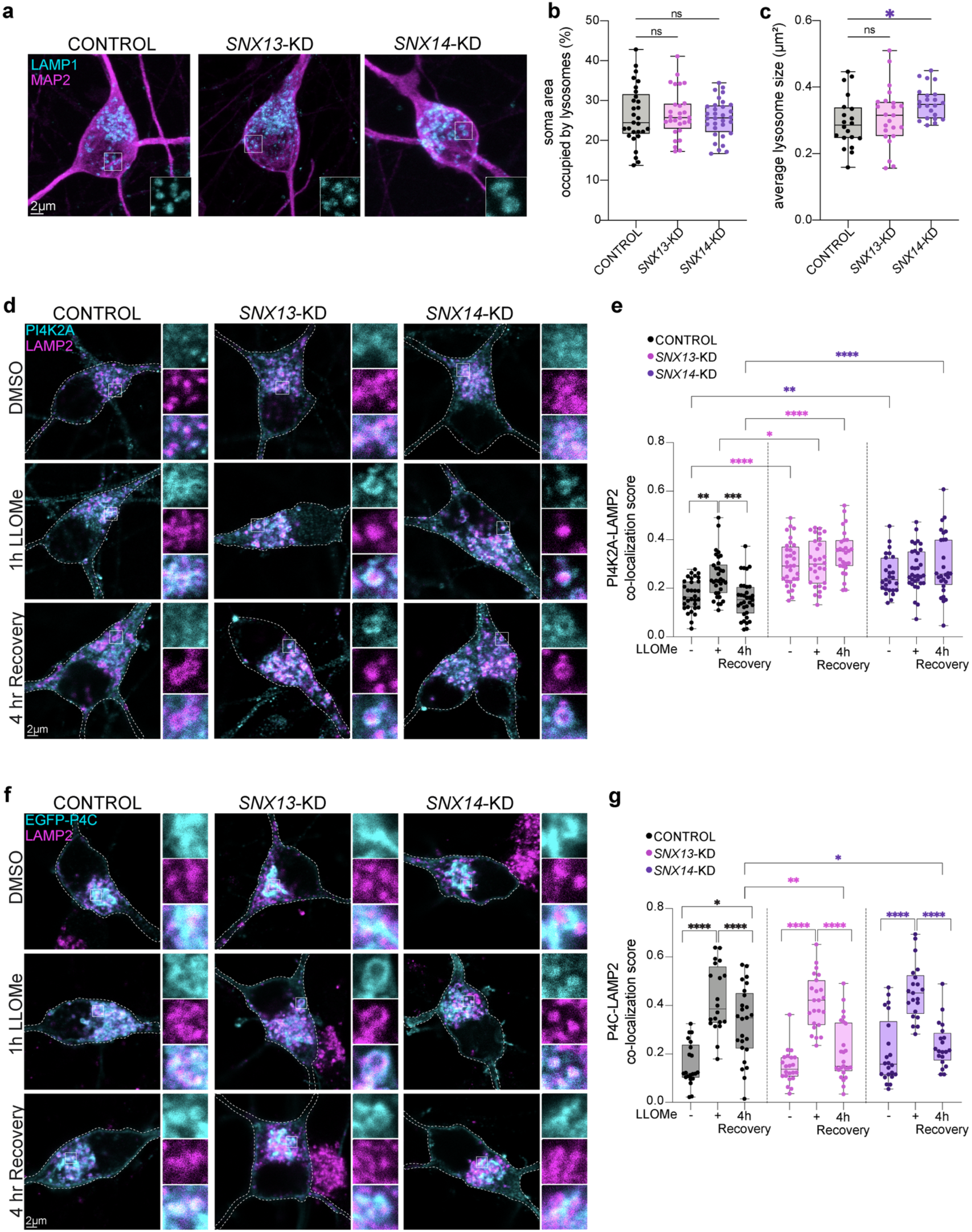
SNX13 or SNX14 deficient human neurons exhibit altered lysosome morphology and PI4K2A-mediated membrane damage response. **a.** Representative images of anti-LAMP1 (cyan) and anti-MAP2 (magenta) in Control, *SNX13*-KD, and *SNX14*-KD D21 neurons. **b.** Box plots showing the percentage of soma area occupied by LAMP1+ lysosomes in n=20-22 neurons from 3 independent experiments. Statistical analysis was performed using ordinary one-way ANOVA followed by Dunnett’s correction for multiple comparisons. **c.** Box plots showing average size of LAMP1+ lysosomes in n=28-32 neurons from 3 independent experiments. Statistical analysis was performed using ordinary one-way ANOVA followed by Dunnett’s correction for multiple comparisons. **d.** Representative images of anti-PI4K2A (cyan) and anti-LAMP2 (magenta) in Control, *SNX13*-KD, and *SNX14*-KD D21 neurons treated with DMSO (control) or 0.5 mM LLOMe for 1h and 4h washout for recovery. **e.** Box plots showing colocalization score of PI4K2A with LAMP2 in n= 27-33 neurons from 3 independent experiments. Statistical analysis was performed using two-way ANOVA followed by Sidak’s correction for multiple comparisons across six families. **f.** Representative images of EGFP-P4C_SidC_ biosensor signal (cyan) and anti-LAMP2 (magenta) of Control, *SNX13*-KD, and *SNX14*-KD D21 neurons treated with DMSO (control), 0.5 mM LLOMe for 1h or 4h recovery **g.** Box plots showing colocalization score of EGFP-P4C_SidC_ and LAMP2 in n=19-25 neurons from 3 independent experiments. Statistical analysis was performed using two-way ANOVA followed by Sidak’s correction for multiple comparisons across six families. Statistical significance is noted as: ns or nothing= not significant, * p<0.05, ** p<0.01, *** p<0.001, and **** p<0.0001. Scale bar: 2 μm.

Next, we examined whether *SNX13-*KD or *SNX14-*KD neurons showed signs of LMP or altered recruitment of repair machinery. Since human cortical neurons do not express Gal3, and ESCRT-III repair components, such as CHMP4B, are undetectable by immunofluorescent staining even after damage induction^57, 58^, we adopted two alternative approaches to evaluate LMP and repair. First, we quantified lysosomal localization of HGS, an ESCRT-0 protein previously shown to accumulate on damaged lysosomes in human neurons^56, 58^. As expected, acute LLOMe exposure increased HGS recruitment to LAMP1^+^ lysosomes in Control neurons, which returned to baseline levels after 4h of recovery following LLOMe washout (Figure S7F, G). Of note, SNX13-KD neurons exhibited reduced HGS accumulation on lysosomes at 1h LLOMe, but the signal continued accumulating during the recovery period and remained significantly elevated compared with Control neurons at 4h after LLOMe washout (Figure S7F,G).

Second, we assessed the lysosomal recruitment of PI4K2A, a lipid kinase which generates PtdIns4P on damaged lysosomes to initiate the PITT repair pathway^10^. Notably, at baseline, *SNX13-*KD and *SNX14*-KD neurons exhibited increased colocalization of PI4K2A with LAMP2^+^ lysosomes, reaching levels comparable to those observed in Control neurons upon acute LLOMe exposure (Figure 7D, E). Furthermore, lysosomal PI4K2A levels remained elevated in *SNX13-*KD *and SNX14*-KD neurons after LLOMe washout, whereas it returned to baseline levels in Control neurons (Figure 7D, E). These findings are consistent with persistent LMP in *SNX13-*KD or *SNX14*-KD neurons.

Given that PI4K2A lysosome pools were boosted in KDs, we next asked whether this increased lysosomal PI4K2A was accompanied by PtdIns4P enrichment on lysosome membranes. To monitor this, we transfected neurons with a plasmid expressing the PtdIns4P sensor EGFP-P4C_SidC_^60^. As expected, EGFP-P4C_SidC_ localized predominantly to the Golgi apparatus in Control neurons at baseline and redistributed to lysosomes following acute LLOMe treatment, indicating the formation of a lysosomal PtdIns4P pool (Figures 7F, G and S7H). LLOMe-induced lysosomal recruitment of EGFP-P4C_SidC_ was also observed in *SNX13-*KD and *SNX14-*KD neurons (Figure 7F, G). Surprisingly, despite increased PI4K2A levels on lysosomes, the lysosomal EGFP-P4C_SidC_ intensity was lower in *SNX13-*KD and *SNX14*-KD neurons than in Control cells, particularly at 4h recovery (Figure 7F, G). These data suggest that loss of SNX13 or SNX14 results in neuronal failure to sustain lysosomal PtdIns4P pools, despite elevated PI4K2A presence on lysosomes. We speculate that some aspect of the PtdIns cycle is perturbed from SNX13 or SNX14 loss, causing blunted PtdIns4P lysosome signaling. Of note, PtdIns itself can be generated from PA^61^. Together, this data suggests that alterations in lysosomal membrane lipid composition caused by the loss of SNX13 or SNX14-mediated ER-lysosome contact and lipid transfer, may lead to perturbed recruitment and function of lysosome membrane repair machinery.

## Discussion

Neurodegenerative diseases are increasingly linked to defects in organelle homeostasis, a particular challenge for long-lived cells, like neurons. Lysosomes are central to this vulnerability as their degradative function requires lifelong maintenance of membrane integrity. In this study, we identify a novel cerebellar neurodegenerative syndrome linked to SNX13 deficiency which is recapitulated in a mouse model we generated. We demonstrate that human SNX-RGS proteins SNX13 and SNX14 are required to maintain lysosome function. Mechanistically, we find their loss alters lysosome acidification and perturbs LMP repair in multiple cell types, including hPSC derived neurons. By applying Lyso-IP proteomics we reveal the dynamics of lysosome proteome remodeling during membrane damage and recovery, and identify perturbed recruitment of lysosome repair proteins in SNX13 and SNX14 deficient cells. Lastly, leveraging the yeast SNX13/SNX14 ortholog NVJ3, we provide experimental evidence these proteins belong to a family of highly conserved lipid transporters with ancestral roles in the maintenance of lysosome membrane composition. Collectively, our findings link SNX-RGS proteins to neurodegeneration and uncover a novel mechanism by which LTPs at ER-lysosome junctions promote lysosome integrity and help maintain lysosomal phosphoinositide signaling, ultimately promoting neuron health.

For lysosome quality control (LQC), recruitment of LTP proteins is proposed to aid repair by delivering fresh lipids to “patch” holes on damaged lysosome membranes^10^. Our work proposes that the PXA and PXC domains of SNX13 and SNX14 aid in this lipid delivery. Given the conserved role for SNX-RGS proteins in localizing to interorganelle junctions, a reasonable model is that SNX-RGS proteins can localize to ER-lysosome junctions and facilitate lipid transport from the ER to lysosome surface in order to maintain lysosome membrane composition and aid in LMP repair upon damage^15, 18, 23, 24, 62^. In support of this, SNX13 has previously been shown to localize to ER-lysosome junctions, as well as influence lysosome cholesterol egress, though it is unclear whether this egress is direct or indirect^24^. Furthermore, previous work indicates SNX14 co-purified with isolated lysosomes and ER membranes, consistent with its role in inter-organelle crosstalk^15, 17^. We and others have also observed SNX13 and SNX14 localize at other membrane contacts, such as ER-LD junctions, during periods of LD biogenesis and growth, suggesting these proteins may function as LTPs in other interorganelle contexts, potentially in a cell or metabolic context dependent manner^18, 24^. Notably, this is similar to other LTPs which are observed at other interorganelle junctions involving the ER in different metabolic contexts^63^. Furthermore, using hPSC neurons, we find loss of SNX13 or SNX14 leads to defective PtdIns4P lysosome pools following membrane damage. We speculate these PtdIns4P perturbations could be due to defective lipid transport by SNX13 or SNX14, including PA, consistent with our findings in NVJ3. We also cannot rule out the possibility that SNX13 or SNX14 may serve as direct transporter of PtdIns or other phospholipids such as phosphatidylserine, thus supplying lysosomes with PtdIns for PtdIns4P biosynthesis. In this model, SNX13 and SNX14 help maintain a PIP cycle at lysosomes, ensuring PIP-dependent lysosome responses to damage.

Our work also indicates NVJ3 and its homologs may play a role in PA/DAG metabolism that may underlie lysosome membrane homeostasis. We show that yeast NVJ3 binds to phospholipids including PA *in vitro*, and that NVJ3-deficient yeast display elevated DAG at lysosomes. Similarly, *NVJ3* deletion can rescue the DAG:PA imbalances of *PAH1* depleted yeast. NVJ3 has also recently been shown to promote the remodeling of lipids at the ER to support LD biogenesis, further emphasizing its role as a LTP involved in neutral lipid metabolism^62^. Of note, DAG is a non-bilayer forming lipid and its accumulation in membranes favors a negative curvature which can disrupt or even rupture organelles like the lysosome^64, 65^. Moreover, DAG and PA metabolism are increasingly associated with neurological disease. For example, a recent multi-omics report indicates that genetic variation among phosphatidic acid phosphatases can modify the severity of Niemann Pick type C disease^66^. Similar to PA/DAG metabolism, lysosomal dysfunction and LMP are emerging hallmarks of neurodegeneration^4, 5, 9, 48^. Our work suggests SNX13 and SNX14 are additional factors linking lysosomes and lipid metabolism to neurological disease.

Importantly, here we establish SNX13 deficiency as the cause of a previously unrecognized cerebellar ataxia syndrome characterized by progressive neurodegeneration. This is supported by a previous report describing a child with global developmental delay and reduced cerebellar volume carrying a homozygous frameshift variant in *SNX13,* considered at the time to be a variant of uncertain significance^67^. Furthermore, our longitudinal imaging demonstrates progressive cerebellar tissue loss, establishing cerebellar neurodegeneration as a hallmark of SNX13 deficiency. Similar to the loss of *Snx14*, *Snx13* loss in mice results in embryonic lethality^33^, suggesting that SNX13 deficiency may also disrupt embryonic development in humans. Consistent with this possibility, the number of identified patients remains small despite our worldwide efforts to identify additional individuals with biallelic loss-of-function *SNX13* variants. As our search for additional patients continues, the *Snx13^PCKO^* mouse model provides functional evidence that SNX13 is required to maintain cerebellar PC function and survival.

SNX13 and SNX14 join a growing list of ER-associated proteins involved in lysosome and lipid homeostasis and neurological disease. Lysosome storage diseases (LSDs), like Niemann Pick type C, are increasingly linked to defects in ER-lysosome crosstalk or lysosome damage repair, suggesting convergent organelle-level mechanisms underlying neurodegeneration^68, 69^. PCs are one of the largest neurons in the brain, and the ER extends throughout the entire neuron from the soma to distant compartments that form many contacts with organelles^70–72^. Notably, SNX13 and other LTPs like SNX14, VPS13D, and NPC1 are linked with cerebellar ataxias, suggesting cerebellar PCs are particularly vulnerable to disturbances in lysosomal and lipid homeostasis^15, 73, 74^. Although it should also be noted that individual loss of SNX13 or SNX14 result in distinct diseases, both share early onset cerebellar ataxia as a prominent clinical feature. Considering this along with the distinct cellular phenotypes caused by depletion of either, we suggest that SNX13 and SNX14 perform nonredundant but partially overlapping roles for maintaining lysosome and lipid homeostasis.

In closing, our work establishes new factors that influence lysosome damage and repair in neurodegenerative diseases. It builds on others in the field revealing ER and lysosome associated proteins as critical modulators of neuron health, including, recent genetic studies that associate SNX13 mutations with early-onset Parkinson’s disease (PD)^75^. Of interest, PD related genes *GBA1*, *LRRK2*, and *VPS13C* are all associated with lysosomal and lipid dysfunction, similar to SNX13 and SNX14^76–78^. A recent study showed that VPS13C acts as an early responder to lysosomal damage potentially to aid in bulk lipid transport from the ER to lysosomes^13, 79^. Our work reveals that SNX13 and SNX14 belong to their own class of non-vesicular LTPs that facilitate lysosome membrane repair, reminiscent of VPS13C or the PITT pathway. Future studies will further dissect how these different players inter-relate in lysosome health, and whether they display distinct cell-type or tissue-specific roles that could influence specific disease states.

## Resource availability

### Lead contacts

Request for information and resources should be directed to the lead contacts, Mike Henne and Naiara Akizu.

### Materials availability

All data needed to evaluate the conclusions stated in the paper are presented in the paper, the supplementary materials and/or the key resources table.

### Data and code availability

Mass spectrometry proteomics data from LysoIPs will be deposited in PRIDE and made available for everyone upon publication of this paper. Yeast vacuole lipidomic data will be deposited in LIPID MAPS and made available for everyone upon publication of this paper. The analysis code for the behavioral and histology data will be made available at Zenodo.

## Data Availability

All data produced in the present study are available upon reasonable request to the authors

## Acknowledgements

We thank members of the Akizu, Henne, and Prinz labs for help preparing the manuscript; the Behavioral Evaluation of Advanced Model Systems (BEAMS) facility in the Center for Brain Research in Development, Genetics, and Engineering (BRIDGE) at Children’s Hospital of Philadelphia for their contributions to behavior data generation; Alexandre Toulmay and Subhra Banerjee of the Prinz lab for conceptual advice; Michael S. Marks for his feedback and for sharing the EGFP-P4C_Sidc_ plasmid with permission of Tamas Balla; Michael E. Ward for the hPSCs engineered to express *NGN2* under doxycycline-inducible system and *dCAS9* constitutively; and HelmohltzZentrum Munchen for the sperm of *Snx13* first-knock out allele mice. K.G. and W.M.H. are supported by NIH NIDDK R01DK126887, NIGMS R35GM119768, and the Ara Parseghian Foundation. V.B.S is supported by NIH F31NS130942 and NINDS T32 NS007413. C.B. is supported by a NIH T32 GM153602-02. N.A. is supported by NIH NINDS R01NS119699, National Ataxia Foundation and Eagles Autism Foundation pilot grants. B.M.C is supported by an NHMRC Investigator Grant 2016410 and ARC Discovery Project Grant 230100552. A.J.E. is supported by NIH NIMH R01MH129970 and W.A.P. by NIH R35GM153315. The Nikon spinning disk microscope at the core facility in UTSW was purchased with a Shared Instrumentation grant from NIH 1S10OD028630-01 to K. Luby-Phelps.

## Author contribution

K.G. and V.B.S. developed and performed experiments. W.M.H. and N.A. conceived and supervised the study. K.G., V.B.S., W.M.H. and N.A. conceptualized the study. E.R., C.B., T.J., S.Y. and S.S. performed experiments. S.Y. and A.J.E. designed, analyzed and interpreted behavioral studies. S.Y. and T.J. performed behavior and histology experiments. K.G., T.R. and J.F.B. performed formal analysis. J.F.B contributed to proteomic data interpretation and visualization. I.C. and D.S. performed lipidomic analysis. J.G.G., U.T., A.P., C.N., J.L., B.K. and A.A., recruited, phenotyped and performed genomic studies in patients. A.J.E., O.S., X.O-G., C.M., R.D., W.P., J.MD. contributed to experimental design and interpretation. B.P. and B.M.C. performed and interpreted structural modeling. K.G., V.B.S., J.F.B., W.M.H., and N.A., wrote the manuscript. All authors read and approved the final version of the manuscript (with exception of U.T. who passed away before the completion of this study).

## Declaration of generative AI and AI-assisted technologies in the manuscript preparation process

During the preparation of this work, the authors used ChatGPT (OpenAI) in order to improve the clarity, grammar and readability of the manuscript. After using this tool, the authors reviewed and edited the content as needed and take(s) full responsibility for the content of the published article.

## Methods

### Human subjects

#### Ethical regulations and study participants

Patients included in this study were identified by connections through GeneMatcher^80^ and national and international collaborations. Patient recruitment and participation were covered by Institutional Review Board approvals at the respective institutions. Ethical approval and informed consent were obtained for participation, phenotyping, sample collection including peripheral blood sample DNA. The authors also confirm that human research participants provided written informed consent for publication of the images in Figure 1 and Supplementary Figure 1. Since this study is focused on gene identification, all patients with confirmed diagnosis are included in the study, regardless of their gender and origin.

#### Identification of pathogenic variants

Pathogenic variants were identified by exome or genome sequencing performed on blood genomic DNA extracted from the probands. When available, blood DNA from parents and siblings was also sequenced or Sanger sequenced. Data analysis was performed to assess quality of sequence reads and variants filtered according to mode of inheritance, evolutionary conservation, pathogenic prediction scores and frequency in the population based on the minor allele minor allele frequency (MAF) collected from gnomAD v4.1.2^32^. Relatively common variants were excluded based on MAF threshold of 0.1%, and functional annotation such as synonymous, non-exonic, and non-splicing-altering were also included. Variants were reported according to standardized nomenclature recommended by the Human Genome Variation Society^81^ on *SNX13* transcript GenBank: https://www.ncbi.nlm.nih.gov/nuccore/NM_001350862.1].

### Mice

#### Animals and Husbandry

All procedures were approved by the Institutional Animal Care and Use Committee (IACUC) at CHOP and were performed in accordance with the Association for Assessment and Accreditation of Laboratory Animal Care (AAALAC) and National Institutes of Health (NIH) guidelines for the care and use of laboratory animals (CHOP, Philadelphia, PA; AAALAC Accreditation #000427; PHS Animal Welfare Assurance D16-00280 [OLAW A3442-01]). Our reporting adheres to the ARRIVE 2.0 guidelines^82^.

Snx13^PCKO^ mice were generated via a gene targeting strategy. The knockout-first targeting allele (Snx13^tm1a^(EUCOMM)^Hmgu^) contained exons 2-4 of the *Snx13* coding sequence, the LacZ reporter and Neo selection cassettes flanked by FTR sites followed by the exon 5 flanked by LoxP sites. A C57BL/6N female was inseminated with sperm of mice carrying the above targeting allele and the offsprings were bred with either Actb:FLPe (Jax #005703) or Pcp2:Cre (Jax #010536) mice. *Snx13*^+/-^; *Pcp2:Cre* females were crossed with *Snx13^flox/flox^* males for generation of experimental mice. In the absence of Cre recombinase, the floxed *Snx13* allele expresses the wild-type sequence is expressed; in the presence of Cre recombinase, there is a Cre-mediated excision of exon 5 leading to an interruption of the *Snx13* gene expression. *Snx13*^PCKO^ mice (*Snx13*^flox/-^; *Pcp2*:Cre) and Control littermates (*Snx13*^flox/+^; *Pcp2*:Cre, *Snx13*^flox/+^ and *Snx13*^flox/-^) of both sexes were generated at Children’s Hospital of Philadelphia (CHOP) as described above. Mice were group housed (2 to 5 per cage) in a HEPA-filtered, closed airflow vivarium system (Enviro-Gard A; Lab Products Inc.) under a 12:12 h light/dark cycle (lights on at 06:15) at 20 to 23°C and 30% to 40% humidity, with standard rodent chow (Standard Rodent Chow 5015; LabDiet) and water available ad libitum. At each cage change, cages were given a nestlet square and a hut; no other enrichment was provided. The genotype of all mice was determined via PCR performed on tail snips obtained at P1-P7 and was re-confirmed for each mouse after they were sacrificed for immunohistochemistry.

### Behavior analysis

#### Experimental Design

Behavior analysis was performed with six cohorts of Control and *Snx13^PCKO^*males and females at 4 months of age. Each cohort contained mixed genotype and sex of animals. Behavior tests were performed in the following order: accelerating rotarod, ataxia scoring battery (ledge test, hindlimb clasping, gait and kyphosis) and vertical pole test. All mice were tested on the accelerating rotarod, and mice from cohorts 2 to 6 were also assessed with the ataxia scoring battery and the vertical pole test. Mice were group housed on a 12 h light/dark cycle with food and water available ad libitum, and all tests were performed during the light phase (8:30 to 11:30 am). Mice were transferred to the testing room at least 30 min before each test. The apparatus was cleaned with 70% ethanol between mice, and with disinfectant wipes between male and female mice, to remove olfactory cues. Investigators were blinded during scoring of behavioral assessments. Whenever possible, offline analysis by computer software was utilized to enhance rigor.

#### Accelerating Rotarod

Motor coordination and motor learning were assessed on an accelerating rotarod with a 3 cm diameter rod (Mouse RotaRod, model 47650; Ugo Basile, Gemonio, Italy) as described^83^. On day 1, mice were habituated to the rotarod rotating at a constant 4 rpm for 2 minutes. This was immediately followed by a trial where rotation was programmed to rise from 4-40rpm in 300 seconds. After a 30-minute intertrial interval (ITI), a second trial was performed, followed by another ITI and a third trial. Three additional trials were performed on the next 2 consecutive days, for a total of 9 trials. A trial was terminated when a mouse fell, made one complete revolution while hanging onto the rod, or after 300s. Mice that fell within the first 20 s were placed back on the rod and the trial was restarted. Latency to fall (time stayed until falling or riding the rod for a single revolution) was determined.

#### Vertical Pole Test

Measures collected include T-turn quality, T-turn time, and time to descend the pole. The vertical pole test was performed as previously described^84, 85^. Briefly, a pole 50 cm tall and 1 cm in diameter, wrapped in non-slip material, was secured upright with its base inside a plexiglass cage filled with bedding. Mice were tested on three consecutive days, no more than 24 h apart, with three trials per day, an inter-trial interval of at least 60 s and 30 s of free exploration at the base after each descent. On each trial, the mouse was placed head-up at the top of the pole and allowed to turn and descend. On days 1 and 2, mice that attempted to climb over the top of the pole were gently redirected downward; no guidance was given on day 3. All trials were video recorded. T-turn time was measured from placement until the mouse had turned fully head-down with its body aligned with the pole, and descending time was measured from the completion of the turn until all four paws touched the base. T-turn and descending times were scored live, and T-turn quality was scored from the videos only at day 1 to capture the naïve response before learning, by an observer blinded to genotype as follows: 0, smooth complete turn; 1, clumsy or hesitant turn, or sideways turn; 2, slip during the turn; 3, fall or no turn. Data of the three trials run per day per mouse were averaged for statistical analysis and representation.

#### Ataxia Test

Ataxia battery test included the ledge test, hindlimb clasp test, gait assessment, and kyphosis assessment as previously described^39^. Ataxia composite score is the sum of the ledge, hindlimb clasp, gait and kyphosis subscores (each scored 0-3 and averaged across repeated observations; range 0-12); the ledge and kyphosis scores in Figure 2F-G are therefore components of the composite rather than independent measures.

#### Ledge Test

In the ledge test, the mouse was placed on the ledge of a conventional mouse cage and observed as it walked along the ledge and lowered itself into the cage (0, walks along the ledge without losing balance and lowers itself into the cage gracefully using its paws; 1, loses footing while walking along the ledge but otherwise appears coordinated; 2, does not effectively use its hind legs, or lands on its head rather than its paws when descending; 3, cannot complete the ledge walk, falls, or requires repeated attempts to descend).

#### Hindlimb Clasp Test

The mouse was suspended by the base of the tail for 10 s (0, both hindlimbs consistently splayed outward, away from the abdomen; 1, one hindlimb retracted toward the abdomen for more than 50% of the suspension; 2, both hindlimbs partially retracted toward the abdomen for more than 50% of the suspension; 3, both hindlimbs entirely retracted and touching the abdomen for more than 50% of the suspension).

#### Gait Assessment

Gait was observed as the mouse walked on a flat surface (0, normal gait, moves fluidly with no ataxic features; 1, slight imbalance or irregular stride pattern; 2, wide-based or staggering gait, or hindlimb dragging; 3, unable to walk normally, crawls, or cannot right itself).

#### Kyphosis Assessment

Kyphosis was scored while the mouse walked or sat on a flat surface (0, easily straightens the spine while walking; 1, mild kyphosis but able to straighten the spine when moving; 2, unable to straighten the spine completely and maintains a persistent mild kyphosis; 3, maintains a pronounced kyphosis while walking or sitting).

#### Composite Ataxia Scoring

Each item of the ataxia test (ledge test, hindlimb clasping, gait, and kyphosis) was scored from 0 (normal) to 3 (severe) on three trials, the three scores were averaged and the four averages were summed (range 0 to 12), as described in^39^.

### Sex as a biological variable

Given that SNX13 and SNX14 deficiencies affect both males and females, we examined animals of both sexes in our study. The main text of the manuscript focuses on describing the effect of the genotype (*Snx13^PCKO^*) in behavior and cerebellar integrity, as our primary goal was to determine whether SNX13 deficiency causes cerebellar neurodegeneration. However, statistical models included sex as a factor, and full results are reported in Table S2. Genotype effects were generally similar in direction in males and females. For turn quality and kyphosis the effect was in the same direction in both sexes, however the male comparisons did not reach significance at n = 16, which reads as limited power rather than a sex difference. For the composite ataxia score, the genotype effect differed by sex although it reached statistical significance in both sexes. Sex differences were also observed in rotarod performance and improvement across trials, regardless of genotype.

### Rationale for pooling Control genotypes

Parents of individuals with SNX13 deficiency are heterozygous and clinically unaffected so far. We therefore did not expect substantial differences among the three Control genotypes, each of which retains at least one functional *Snx13* allele (like heterozygous parents): *Snx13*^flox/+^; *Pcp2*:Cre, *Snx13*^flox/+^ and *Snx13*^flox/-^. Consistently, in our pilot analysis, vertical pole and ledge scores did not differ significantly among these genotypes (p = 0.38 and p = 0.19, respectively). All control mice scored 0 for kyphosis, and neither Purkinje cell density nor molecular layer thickness differed significantly among control genotypes in lobules II and III. In lobule X, Purkinje cell density differed among control genotypes (p = 0.02), with the highest density in *Snx13^flox/+^*mice (∼45) and the lowest in *Snx13^flox/−^* mice (∼36.5). Molecular layer thickness in this lobule did not differ significantly (p = 0.06). Overall, although the small group sizes limits the conclusions of this pilot study, we did not observe overt differences between Control genotypes..

### Immunofluorescence (IF) labeling

Mice were deeply anesthetized with isoflurane and underwent transcardial perfusion with 15 ml of 1X PBS followed by 15 mL of 4% paraformaldehyde (PFA) in PBS. Whole brains were extracted and postfixed for 24 hours in 4% PFA at 4C and washed 3 x 10 mins in 1X PBS. Parasagittal brain sections were cut at 50 μm intervals using a vibratome (Leica). On the day of staining, sections were washed in 1X PBS and incubated in blocking solution (4% normal donkey serum (NDS) (Sigma-Aldrich, D9663), 0.5% Triton X-100 in PBS) for 1 hour at room temperature. The sections were transferred into primary antibody solution (2% normal donkey serum, 0.1% Triton X-100 in PBS) and incubated at 4C overnight. Next, sections were washed 3 times with 0.1% Triton X-100 in PBS and incubated for 2 hours at room temperature in secondary antibody solution (2% normal donkey serum, 0.1% Triton X-100 in PBS). Finally, sections were washed 3 times with 0.1% Triton X-100 in PBS and were mounted on glass slides using homemade Mowiol (Sigma 81381)-DABCO (Sigma D27802) mounting medium. Images were acquired using a confocal microscope (Leica TCS SP8 X) and image processing was performed with ImageJ software.

### Human pluripotent stem cells

#### Maintenance

WTC11 hPSCs engineered to express NGN2 under a doxycycline inducible system in the AAVS1 safe harbor locus and pC13N-dCas9-BFP-KRAB targeted to the human CLYBL intragenic safe harbor locus were cultured on Matrigel (Corning, #354277) coated plates in mTeSR1 or mTeSR Plus media (STEMCELL Technologies, #85850 or #100-0276, respectively). The medium was changed every day and cells were passed when they reached 70% confluence, approximately every 5–7 days, using ReLeSR (STEMCELL Technologies, #100-0484) at a 1:20 ratio. Structural variant analysis with an Illumina Infinium GSA-24v3 BeadChip identified a single CNV covering chr7:69282553-70158495, known to be carried in the parental WTC11 hPSCs. Cells were tested monthly for potential mycoplasma contamination by PCR using DNA isolated from hPSC pellets.

#### Human pluripotent stem cell transduction

20bp sgRNA sequences targeting either SNX13, SNX14, and Non-Targeting Control were designed using the Broad Institute’s CRISPick software, and were cloned into the lenti-sgRNA-blast backbone (Addgene, Plasmid #104993. For lentiviral production, HEK293-T cells were plated at 1×106 cell/well density in a well of a 6-well plate with 2 mL of media (1X DMEM (Gibco #11995073), 10%FBS (Tissue Culture Biologicals #101), and 1%Penn/Strep (Gibco #15140122). The following day, cells were transfected using Lipofectamine 2000 (Invitrogen #11668027) with 1.06 μg MDLg/RRE (gag/pol expression plasmid), 0.57 μg MD2.G (VSV-G envelope expressing plasmid), 0.4 μg pRSV-Rev (rev expression plasmid), and 1.6 μg of transfer vector (sgRNAs) per well. 6-8 hours post-transfection, media was removed and cells were washed with 500 μL of mTeSR and then replenished with 2 mL of mTeSR. 48 h later 2 mL of media with lentiviruses were collected and filtered through 0.45 μm pore filters. 100-200 μL of filtered lentiviral media was introduced into CRISPRi hPSCs via lentiviral delivery using LentiBlast (ozbiosciences; #LBPX500) according to the manufacturer’s protocol. Cells were selected with 4 μg ml−1 blasticidin HCl (Gibco #A1113903) for 2–4 days until WT non-transduced cells die.

#### Differentiation to iNeurons

WTC11 NGN2 hPSCs were differentiated using doxycycline-induced expression of NGN2 based on previously described methods^59^. Day 0 iPSCs were collected using Accutase by aspirating media, washing 2x with DPBS, and then incubating with Accutase at 37C for 10 min. Accutase was then diluted with pre-warmed DMEM/F12, iPSCs were pelleted, and then resuspended in fresh mTeSR Plus with 10 μM Y-27632 for counting. iPSCs were plated in Induction Medium: DMEM/F-12, 1x N2 supplement, 1X MEM Non-Essential Amino Acids (MEM-NEAA) (ThermoFisher #11140-050), and 1X GlutaMAX, supplemented with 10nM Y-27632 and 2ug/mL doxycycline hydrochloride (Sigma-Aldrich #D3072). iPSCs were plated at a concentration of 1×10^6^ cells/well on pre-coated GFR Matrigel plates. On Day 1, Y-27632 was removed. On Day 3 or 4, pre-differentiated cells dissociated using Accutase, counted with Countess II Automated Cell Counter, and either cryopreserved in ATCC StemCell Freezing Media (ATCC #ACS3020) or plated on pre-treated plates in Maturation Media: Neurobasal Plus Medium (ThermoFisher # A3582901), 1x B27 Plus supplement (ThermoFisher #A3582801), 1x CultureONE supplement (ThermoFisher #A3320201), 1X MEM-NEAA, and 1X GlutaMAX, supplemented with with 10 μM Y-27632, 2ug/mL doxycycline hydrochloride, 10ng/mL NT-3 (PeproTech, 450-03), 10ng/mL BDNF (R&D Systems #248-BDB), 10ng/mL GDNF (R&D Systems # 212-GD), and 100 μM L-ascorbic acid (Sigma-Aldrich #4034-100). Thereafter, neurons were fed every 4–5 days by replacing 50% of the medium volume.

### Hela and HEK293T cell culture

#### Maintenance

Hela and HEK293T cells (Lenti293T, Takara Bio) were cultured in DMEM (Sigma-Aldrich #D5796) supplemented with 10% fetal bovine serum (Sigma-Aldrich #F4135), 1% penicillin streptomycin solution (Corning #30-002-Cl), and 25 mM HEPES (Sigma-Aldrich #H0887).

### siRNA-based gene knockdown

RNAi was performed using Lipofectamine RNAiMAX (ThermoFisher #13778150). For SNX13 and SNX14 knockdowns, pooled siRNAs or Silencer Select negative control siRNA #1 were diluted in 250 μL Opti-MEM and then into full media with Lipofectamine RNAiAX used at a concentration of 3 μL/mL. Pooled siRNAs (Dharmacon ON-TARGETplus Smartpool siRNA) were used at 30nM for SNX13 knockdown and 10nM for SNX14 knockdown. siRNA diluted in Opti-MEM (ThermoFisher #31985062) was added dropwise to Lipofectamine RNAiMAX diluted in Opti-MEM and incubated for 5 min before adding to cells in growth media. Media was changed after 24 h, and the total knockdown time was 48hrs.

1mM L-Leucyl-L-Leucine methyl ester monohydrochloride (LLOMe; Sigma #L7393) was used for all LLOMe-induced lysosomal damage experiments. Briefly, Hek293T or HeLa cells were treated with either DMSO (vehicle control) or LLOMe for a desired timepoint (i.e., 1min > 1h) in complete media, followed by a complete washout with either 1x DPBS or DMEM and replaced with complete media for 1 or 4 hours to allow for recovery. After each treatment, cells were fixed with 4% PFA and IF was performed.

#### LLOMe-induced lysosomal damage

1mM or 0.5mM L-Leucyl-L-Leucine methyl ester monohydrochloride (LLOMe; Sigma #L7393) was used for all LLOMe-induced lysosomal damage experiments in Hela/HEK293T or hPSC derived neruons respectively. Cells were treated with either DMSO (vehicle control) or LLOMe for a desired timepoint (i.e., 1min > 1h) in complete media, followed by a complete washout with either 1x DPBS or DMEM and replaced with complete media for 1 or 4 hours to allow for recovery. After each treatment, cells were fixed with 4% PFA and IF was performed

### Immunofluorescence (IF) labeling

Neurons were fixed with prewarmed 37 °C 4% paraformaldehyde (PFA)/PBS for 10–15 min at RT, washed 3x with PBS and were subsequently stored at 4 °C or processed for IF. All antibodies used can be found in key resources table.

#### Saponin-Based

The majority of neuronal lysosomal stainings were performed using a saponin-based staining protocol. In short, neurons were blocked in a blocking buffer (1.5% Glysine, 3% BSA, 0.01% Saponin in 1X PBS) for 1h at RT. Neurons were incubated in primary antibody diluted in antibody diluent (0.1% BSA, 0.01% Saponin in 1× PBS) on a shaker overnight at 4 °C. After washing with 1X PBS for three times, AlexaFluor – conjugated secondary antibodies (Invitrogen # A31570 or A78947) along with 1μg/uL DAPI diluted in antibody solution were added to neurons for 2h at room temperature. Finally, neurons were washed with 1X PBS 3 times. Coverslips were mounted with Mowiol-DABCO-based mounting medium.

#### Tween-Based

Briefly, hPSCs were plated onto Matrigel coated glass coverslips (Electron Microscopy Sciences #72290-04). Cells were fixed with 4% PFA for 10 min at room temperature, permeabilized and blocked in blocking buffer (4% goat serum (Sigma-Aldrich, G9023) and 0.1% Tween-20 in PBS) for 1 h at room temperature. Cells were immunostained overnight at 4 °C with the primary antibodies in 2% goat serum. Next day, cells were incubated for 2h at room temperature with Alexa Fluor – conjugated secondary antibodies and 1 μg/ml DAPI and mounted with Mowiol-DABCO-based mounting medium.

HeLa cells were grown in glass-bottom dishes for imaging (Cellvis #D35-14-1.5-N) and fixed with 4% paraformaldehyde in PBS (ThermoFisher #J61899.AK) for 15 min at RT. Cells were washed with PBS and permeabilized with 0.5% Triton X-100 (Sigma-Aldrich) in PBS for 5 min. After washing twice with PBS, the cells were blocked in IF buffer (3% BSA in PBS) for 30 min. Cells were incubated with primary antibody in IF buffer for 1 hr or overnight, washed 3x 5-min with PBS, incubated with secondary antibody in IF buffer for 1 hr, and washed 3x 5-min with PBS. Primary antibodies to visualize lysosomes in HeLa cells include: anti-TMEM192 (Abcam #AB185545) and anti-LAMP2 (BD Biosciences #555803). Lysosome damage and repair was assessed with anti-Galectin3 (BD Biosciences #556904) and anti-CHMP4B (Proteintech #13683-1-AP). LDs were visualized by 20 minute incubation with 1:1000 Lipi-Blue (Dojindo #LD01). Cells were kept in PBS at 4°C prior to imaging.

### Microscopy and Image analysis

HeLa or hPSCs images were acquired on a Nikon spinning disk camera at 40x or 60x magnification, a step size of 0.5 μm, and a total volume of 10 μm. Neuron images were acquired with Leica SP8 confocal microscope at 63x magnification, a step size of 0.3 or 0.5 μm. All images were processed and analyzed using ImageJ (NIH, https://fiji.sc).

#### Lysosome measurements

Max-projections were used to manually draw cell or soma boundaries to generate a region of interest (ROI). LAMP1/2 was thresholded and binarized (default) within these ROIs to achieve lysosomal area. The cell/soma area occupied by lysosomes was obtained by dividing the lysosomal area by the cell/soma area. In addition, the brightness (raw integrated density) of max-projected lysosomes were also measured. Notably, to obtain the average size of lysosomes in neurons, the elliptical tool was used to manually outline individual (>20) lysosomes per neuron.

#### Co-localization

Raw, unprocessed, non-overlapping images were imported into ImageJ. A mid-z plane was used to manually draw a boundary around the soma to generate an ROI, followed by background subtraction using a 50 pixel rolling ball radius. Individual channels were thresholded independently to exclude background and non-specific staining noise and converted to binary masks. Co-localization between lysosomes (LAMP1/2) and the marker of interest (HGS, P4C, or PI4K2A) was determined using the “AND” function using the image calculator. Data are plotted as the fraction of lysosomes that are positive for the marker of interest.

#### Lysosome-nucleus distance measurements

Distance between thresholded lysosomes and the center of the nucleus were calculated using the published ImageJ macro by FNSoria (Zenodo DOI:328500157). Using brightfield, boundaries around the cell and nucleus were manually drawn. Max projections were performed on lysosome images, and the images were enhanced for segmentation using the default sigma on a Difference of Gaussians filter. The average distance between lysosomes and the nucleus center are reported per cell.

#### Relative pH of acidic compartments

LysoSensor Yellow/Blue DND-160 (Invitrogen, #L7545) was incubated with live cells at 50µM in warm FluoroBrite DMEM (Gibco #A18967-01), then incubated at 37 °C for 5 minutes. Cells were washed twice warm PBS, and replaced with warm FluoroBrite for imaging. Cells were then imaged on the Nikon Spinning disk with the dual camera feature, in a 37 °C incubation chamber. Blue and yellow images were taken simultaneously. For quantification, images were sum-projected and background was subtracted using the rolling ball set to 50. Cell outlines were drawn manually using brightfield and collected as ROIs, and the mean gray value was taken for each channel. The values for the yellow channel were divided by the blue channel, and the resulting values were normalized to the control cells per replicate to give relative acidity.

#### Galectin-3 (GAL3) recovery

To measure damage at each timepoint, a ratio of cytoplasmic-to-nuclear signal was measured. The cytoplasmic compartment was defined using the XOR operation between the whole cell and nuclear ROIs. To account for background noise and cell size variability, the corrected total cell fluorescence (CTCF) was calculated: CTCF = Integrated density - (area x mean background). The nuclear to cytoplasmic ratio was determined by dividing the nuclear CTCF by the cytoplasmic CTCF. Background was determined by averaging the mean grey value of two regions of interest without visible GAL3 signal per field of view. Since the CTCF ratios were statistically different at baseline between KDs, we also compared the magnitude of damage vs the magnitude of recovery (percent recovery).

#### CHMP4B recovery assay

To quantify the amount of CHMP4B intensity on lysosomes, a custom macro was used in Fiji. To segment the lysosomes, background subtraction and median filtering was used to reduce noise on the LAMP2 channel. Then, automated thresholding was used to create a binary mask of the lysosomal regions. For each cell, nuclear and whole-cell ROIs were drawn using the brightfield channel. The cytoplasmic ROI was defined by applying the XOR function between whole cell and nuclear ROIs. The AND function was used between the whole-cell ROI with the total lysosomal mask to isolate the lysosomal compartments of each cell. For each image, background subtraction was determined by measuring the mean grey value of two ROIs without visible signal in the CHMP4B channel. The mean background value was calculated by averaging these measurements per field of view. The mean intensities were obtained for all ROIs. Background-corrected values were calculated by subtracting the mean background intensity from the raw mean intensity. The values were reported in different ways:

1. *Normalized intensity:* Background-corrected values of all timepoints and KDs were normalized by the average background-corrected value of the control cells at timepoint 0 (baseline), per replicate. These values would represent the amount of CHMP4B recruited on lysosomes.
2. *Induction rate:* For each KD background (including control), background-corrected values of all timepoints were normalized by the average background-corrected value of timepoint 0 (baseline), per replicate. These values would represent a rate/change; how CHMP4B recruitment has changed through the given timepoint.

### TEM preparation and imaging

Cells were fixed on MatTek dishes with 4% paraformaldehyde, 2.5% (v/v) glutaraldehyde+ 0.2% picric acind in 0.1M sodium cacodylate buffer then transferred to the Electron Microscopy Core Facility for further processing. After five rinses in 0.1 M sodium cacodylate buffer, samples were post-fixed in 1% osmium tetroxide and 1.5% K3[Fe(CN)6] in 0.1 M sodium cacodylate buffer for 1 hour at 4 degrees C. Cells were rinsed with water and en bloc stained with 0.5% aqueous uranyl acetate in 25% methanol overnight at 4 degrees C. After five rinses with water, specimens were stained with 0.0M lead nitrate in 0.03M L-aspartate acid in 60 degree C oven for 30 minutes. After 5 water washes, samples were dehydrated with increasing concentration of ethanol, infiltrated with Embed-812 resin and polymerized in a 60oC oven overnight. Embed-812 disc were removed from MatTek plastic housing by submerging the dish in liquid nitrogen. Pieces of the disc were glued to blanks with super glue and blocks were sectioned with a diamond knife (Diatome) on a Leica Ultracut UCT (7) ultramicrotome (Leica Microsystems) and collected onto copper grids, post stained if needed with 2% Uranyl acetate in water and lead citrate. Images were acquired on a JEM-1400 Plus transmission electron microscope equipped with a tungsten source operated at 80 kV using an AMT-BioSprint 16M CCD camera.

### RNA extraction, cDNA generation, and qPCR

For hPSCs and neurons, total RNA was extracted from samples using the RNeasy Plus Mini Kit (Qiagen #74034). For knockdown analysis, directed reverse transcription was performed using 0.5μg of total RNA with SuperScript™ III First-Strand Synthesis System (Invitrogen #18080051). For quantification of transcripts by RT-qPCRs, 0.5μg of total RNA was retrotranscribed with oligodTs and SuperScript™ III First-Strand Synthesis System following standard protocol (Invitrogen # 18080051). Next, qPCR was performed in a 10µL reaction volume containing 4.6µL of cDNA sample, 0.5 µM of forward and reverse primers (key resources table) and 1X Power SYBR Green PCR Master Mix (Applied Biosystems #4367659) in a Biorad CFX384 Touch Real-Time PCR. Primers were designed using Benchling’s Molecular Suite Primer3 program or NCBI Blast.

For HeLa and HEK293T cells, RNA was extracted from cultured cells using Trizol (ThermoFisher #15596018). Briefly, cell pellets were washed with PBS and centrifuged at 400xg for 5 min. 500μL Trizol was added, mixed by pipetting, then incubated for 5 min at room temperature. 100μL chloroform was added, and the extraction mixtures were shaken for 15 s before RT incubation for 3 min and centrifugation at 12000xg for 15 min at 4°C. The upper aqueous fraction was transferred into a new tube, and isopropanol was added in a 1:1 ratio. The tubes were inverted 4x before being incubated at room temperature for 10 min and centrifuged at 12000xg for 10 min at 4°C. The supernatant was removed, and the pellet was washed with 75% ethanol, vortexed, centrifuged at 7500xg for 10 min at 4°C, and the final supernatant was aspirated. The pellet was dried, resuspended in nuclease-free water, and incubated at 55°C for 10 min before being stored on ice. To synthesize cDNA, 1 μg of RNA was combined into a 20 μL 1x mixture of iScript Reverse Transcription Supermix for qRT-PCR. Cycler conditions were 5 min at 25°C, 20 min at 46°C, 5 min at 95°C, then 4°C hold. For qRT-PCR, cDNA was diluted to 500 ng/μL, and mixed with SsoAdvanced Universal SYBR Green Supermix and 1.6 μM primers with 1 μg of cDNA per 20 μL reaction. qRT-PCR was performed on a BioRad CFX96 Real-Time system, and mRNA expression data was normalized to expression of 36B491,93.

### Yeast

#### Generation of Yeast strains

Saccharomyces cerevisiae was constructed in the genetic background of W303 (ade2-1; leu2-3; his3-11, 15; trp 1-1; ura3-1; can1-100). Endogenous deletion and tags of yeast genes were created using PCR-based homologous recombination via lithium acetate transformation. All knockouts and knockout combinations were generated using pFa6a plasmid cassettes for gene replacement with natMX6, kanMX6, and/or hygMX6. Yeast endogenously expressing Nvj3-mNeonGreen (mNG) and Nvj1-mCherry were generated using longtine PLT plasmid cassettes: PLT-mNG-Hygromycin (hygMX6) and PLT-mCherry-G418 (kanMX6). Strains used for vacuole purification, also expressed Vph1-mCherry, generated using PLT-mCherry-G418. To express the mutated Nvj3 (I233N) from its endogenous locus, Nvj3’s open reading frame was replaced by a Nvj3 (I233N)-NAT cassette. Proper chromosomal integrations were validated by PCR with primers homologous to the selection marker, and/or flanking the open reading frame. We received the DAG biosensor (PKC D1-GFP) from Alexandre Toulmay (William Prinz Lab), and expressed it exogenously in yeast, via a modified lithium acetate transformation for plasmid expression. To analyze localization, Nvj3-mNG and Nvj3 (I233N)-mNG were also expressed on plasmids with the endogenous promoter (1000 bp upstream), from pRS315 plasmids.

Plasmids were generated via HiFi Assembly (NEB #E2621). To create the plasmid used to integrate Nvj3 (I233N) into the chromosome: 1) Nvj3 was endogenously tagged with mNeonGreen, 2) genomic DNA was extracted from Nvj3-mNG tagged strain, 3) Nvj3-mNG was cloned into pBp73G using XbaI and XhoI cutsites, 4) overlap extension PCR was used to mutagenize pBp73G-Nvj3-mNG to create pBp73G-Nvj3(I233N)-mNG, and 5) Nvj3(I233N) was amplified and AscI and BglII was used to clone into the longtine pFa6a series plasmid pFa6a-natMX. To create pRS315-Nvj3-mNG-LEU and its I233N mutant, Nvj3-mNG from genomic DNA was cloned into pRS315 using XhoI and SphI and overlap extension PCR was performed for mutagenesis.

#### Growth Assay

Yeast were grown in 2% dextrose medium to logarithmic growth phase, washed twice with water, and adjusted to an OD_600_ of 0.25. Serial dilutions were prepared in water (4x, 10-fold) and spotted onto 2% dextrose plates (YPD or SCD). The plates were incubated for 2.5 days at 30°C and images were taken with white light setting on the BioRad ChemiDoc Touch Gel Imager.

#### Lipid extraction, thin-layer chromatography (TLC), and quantification

Yeast was grown in synthetic complete media (SCD) to late stationary phase. Approximately 100 OD_600_ units were collected, and the wet weight of the pellets were normalized to the lowest value. Lipid extraction was performed using a modified Folch method^86^. Cells were resuspended in water with 0.5-mm acid-washed glass beads (Sigma-Aldrich, #G8772-500G) and lysed by three 1-min cycles on a MiniBeadBeater. Chloroform and methanol were added to the lysates, achieving a 2:1:1 ratio of chloroform:methanol:water. Samples were vortexed and centrifuged to separate organic and aqueous phases, and the organic phase was collected. This procedure was repeated twice. The organic solvent phases were combined and washed twice with 1 mL of 1.0 M KCl. Lipid samples were dried under argon gas and resuspended in chloroform to a final concentration of 1 μl of chloroform per 1 mg of cell pellet wet weight.

Extracted lipids were spotted in duplicate onto heated glass-backed silica gel 60 plates (Sigma-Aldrich, #1057210001). Each plate contained at least 5 spots of different concentrations of lipid standard, either: neutral lipid reference standard mixture (Nu-Chek Prep; no. 18-5C, prepared in chloroform to 10 mg/ml and diluted to 1 μg/μl before loading onto the plate) or a custom standard mix for phospholipids-2.5 μg/mL of PA (Avanti #A84128), PE (Avanti #A84021), and PC (Avanti #A84051) in chloroform. Neutral lipids were separated in an 80:20:1 hexane:diethyl ether:glacial acetic acid solvent. Phospholipids were separated with two subsequent solvents: 65:25:4 chloroform:methanol:water, followed by a secondary solvent of 100:20 isohexane:ethylacetate. All TLC bands from both standards and samples were visualized by spraying dried plates with cupric acetate in 8% phosphoric acid and baking at 145°C for at least 30 min for neutral lipids and ∼1hr for phospholipids.

Baked TLC plates were scanned and processed for quantification using Image J. Using the rectangle and measure tools, the band integrated density was taken from the standards to create a standard curve where the x-axis was used to calculate the lipid mass in micrograms. Sample band intensities were then plotted on these standard curves to calculate the mass of each lipid in each sample. The neutral lipid standards did not contain DAG, thus relative DAG abundances were calculated by normalizing the value of lipids in experimental samples to wild type samples.

#### Fluorescent Imaging and analysis

Yeast were grown to logarithmic growth phase at 30°C in the appropriate synthetic medium, either with complete amino acids or amino acid drop-out for plasmid retention. After washing once with water, cells were imaged on glass slides on the Nikon Spinning Disk microscope at 100x in room temperature. Z-series were captured at a step size of 0.4μm and a total volume of 5.0μm. To visualize yeast lysosomes (vacuoles), cells were incubated with a final concentration of 8μM FM-464 dye (ThermoFisher, #T13320) for one hour during the logarithmic phase. Linear adjustments for visualization purposes were made using ImageJ (NIH). For analysis of DAG vacuole enrichment, 2 line scans were performed for each cell and the average signal from the vacuole was divided by the average signal from the plasma membrane.

#### Vacuole Purification by density centrifugation

Cultures in 1 mL YPD were prepared and grown in a 30°C shaker ∼18hr overnight. The next morning, the cultures were diluted to 0.1 OD in 10mL SCD. After ∼8 hr incubation, yeast were diluted to ∼0.004OD in 1L synthetic complete media into a 2L non-baffled flask shaking at 200 rpm. Yeast were collected in logarithmic growth phase after ∼18h (OD∼0.7). Out of the liter, 50mL was collected for whole cell lipidomics, and 950 mL were left for processing for vacuole purification. The cells were harvested by benchtop centrifugation (3,000 *g* for 30 min) and weighed, yielding ∼2g of wet weight (wwt) cell pellet. Pellets for whole cell lipidomics were flash frozen and stored in -80 °C.

Vacuole isolation and purification was performed similar to that previously described in^87^, where the initial vacuole purification was expanded from Wiederhold *et al*.^88^ with several modifications. The cell pellet was resuspended in 10 ml of 100 mM Tris-HCl, pH 9.5, then 100 μL of 1M DTT (Fisher Scientific #BP172) was added. After incubation at 30°C for 10 min, cells were centrifuged at 4,000×*g* for 5 min, washed with 10 ml of water, washed with 10 ml of 1.1 M sorbitol, then resuspended in 5 ml of 1.1 M sorbitol. To generate spheroplasts, 6 mg Zymolyase 20T (Sunrise Science #N0766391) per gram wwt cells was dissolved in 1 ml of 1.1 M sorbitol, then added to the cell suspension. After 45 minute incubation at 30°C (with manual swirling every 15 min), all procedures were conducted on ice or 4 °C. Spheroplasts were washed by centrifuging through a 7 ml layer of 7.5% Ficoll 400 (VWR) in 1.1 M sorbitol at 4,000×*g* for 20 min. The spheroplasts were resuspended in 7 ml 12% Ficoll 400 in 10 mM Tris-MES, pH 6.9. For lysis, the suspension was transferred to a dounce homogenizer (15 ml, Wheaton) and homogenized with 15 strokes of a tight B pestle. The suspension was transferred to a centrifuge tube (Beckman Coulter #344059) and 4 ml of 12% Ficoll 400 in 10 mM Tris-MES was layered on top. Centrifugation was performed at 50,000×*g* for 35 min in a SW 41 Ti rotor (Beckman Coulter). Crude vacuoles were isolated from the top layer and collected using an inoculation loop as well as a 1mL pipet; and diluted in 4 ml 12% Ficoll 400 in 10 mM Tris-MES.

Crude vacuoles were then further purified to rid of membrane contaminants, especially LDs. Crude vacuoles were lightly homogenized in a dounce homogenizer by stroking 5 times with the B pestle. 4 ml of vacuole suspension was transferred to a centrifuge tube, then layered with 4 ml 8% Ficoll 400 in 10 mM Tris-MES and 4 ml 4% Ficoll 400 in 10 mM Tris-MES. The vacuoles were further separated from the mixture through density centrifugation at 50,000×*g* for 35 min in a SW 41 Ti rotor. After centrifugation, the top layer was collected as before. Vacuoles were then diluted in 2mL of 0.6 M sorbitol in 5 mM PIPES-KOH, pH 6.8. They were then purified through a sorbitol/sucrose gradient: two volumes (2 ml) of vacuole suspension were layered on top of one volume (1mL) of 0.4 M sorbitol, 0.2 M sucrose in 5 mM PIPES-KOH, pH 6.8, which was layered on top of one volume (1mL) of 0.36 M sorbitol, 0.24 M sucrose in 5 mM PIPES-KOH, pH 6.8. After centrifugation at 3500 x g for 30 min, vacuoles sedimented at the bottom of the tube. The supernatant was removed and the vacuoles were resuspended in 400uL of 0.6M sorbitol buffer. Vacuole integrity was assessed by viewing large intact particles that were positive for Vph1-mCherry. Imaging was done on the EVOS FL Cell Imaging System, with a 60x oil objective. Vacuoles were then flash frozen and kept at -80 °C before lipidomics analysis.

#### Lipidomics Analysis of Yeast and Vacuole Compartments

All solvents were either HPLC or LC/MS grade and purchased from Sigma-Aldrich. All lipid extractions were performed in 16 × 100 mm glass tubes with PTFE-lined caps. Glass Pasteur pipettes and solvent-resistant plasticware pipette tips (Mettler-Toledo) were used to minimize leaching of polymers and plasticizers.

For yeast, the pellets were re-suspended in 500uL PBS and transferred to a 2mL bead beating tube containing glass beads and they were beaten on an Omni bead-ruptor, and transferred to glass tubes along with another 500uL PBS prior to lipid extraction. Lipids from yeast and isolated vacuoles were extracted using MTBE method^89^. The samples were suspended in Methanol (1mL) mixed with 0.1% HCl (diluted from 50mM HCl in Methanol) and pipetted into a silanized glass tube (16×100 mm) with a PTFE-lined cap (Fisher Scientific, Pittsburg, PA). MTBE (2mL), and PBS (1mL) were added. The tube was vortexed vigorously after each addition. The sample was then centrifuged at 3500K for 5 mins. The upper layer was collected in a fresh silanized tube, and the solvent was evaporated under N2. After evaporation, 20uL of 1:5 diluted Splash II Lipidomics Mass Spec Standard (A83709 Avanti Research) was added to it and the lipids were re-suspended in 3:1 MeOH:DCM (500uL). LC-MS/MS was done using a Shimadzu LC system equipped with a SCIEX Triple quad 6500+ mass spectrometer on MRM mode.

Phospholipids were chromatographically separated using a HILIC chromatography on Phenomenex Luna NH2 (150mm x 2.0mm) 5um column. Mobile phase A: 9:1 Acetonitrile: Dichloromethane (v/v) and mobile phase B: 1:1 Acetonitrile: water (v/v) + 20mM ammonium acetate. The column was maintained at 35 C temperature with a constant flow rate of 0.4 mL/min, and an injection volume of 5uL. After injection, the percentage of B was maintained at 5% for 1.5 mins, and then was gradually increased to 10, 20, and 70 at 1.6 mins, 10mins, and 15 mins respectively. The B percentage was kept constant at 70% for 5 mins, and then brought down to 5% at 20.1 mins. The chromatographic run was completed at 25 mins after a re-equilibration period at 5% B. Mass spectrometer settings were as follows: Ion source gas1-50 psi; gas 2 – 55psi; CAD gas 9psi; curtain gas 30 psi; source temperature-300 C; ion spray voltage for positive ion mode-5500 kV; negative ion mode-4500 kV. Sphingomyelins (SM) and lyso-phosphatidylcholines (LPC) were detected in positive ion mode as protonated ions ([+H]), with the head group fragment as the product ion (184.0 in both cases). All the phospholipids other than Phosphatidylcholine (PC) were detected in negative ion mode having the deprotonated ion ([-H]) as the precursor ion and the fragment corresponds to the fatty acid. PCs were detected as their formate adduct with the same fragmentation pattern.

Neutral lipids were separated using reversed phase chromatography on an Agilent Poroshell 120 EC-C18 (150mm x 2.1 mm) 2.7 um column. Mobile phase A was 1:1 Acetonitrile: water (v/v) + 20mM ammonium acetate and B was 9:1 Isopropanol: Acetonitrile +5mM ammonium acetate. The column was maintained at 40 C temperature with a constant flow rate of 0.4 mL/min, and an injection volume of 5uL. The mobile phase B gradient went as follows: from 0 to 0.5 mins – 20%; 0.5 to 2 mins – 20 to 70 %; 2 to 15 mins – 70 to 100%; 15 to 20 mins – maintained at 100%; 20 to 20.1 mins – 100 to 20%; 20.1 to 25 mins – maintained at 20%. The mass spectrometer was set to gas 1-20psi; gas2-50psi; CAD gas 9psi; curtain gas 30 psi; source temperature-250 C; ion spray voltage-5500 kV. Both TAGs and DAGs were detected as their ammonium adducts in the positive ion mode and fragmented by the neutral loss of fatty acids.

Mass spectra were analyzed using SCIEX OS software by comparing the response of the endogenous species to internal standard response. Lipid values represent area under the curve for each lipid detected through a particular solvent/method. These values were normalized by the sum of these areas for each method per sample.

### Recombinant protein synthesis and purification

Plasmid pET15b-6xHIS-MBP-(codon optimized) Nvj3-StrepII was generated using HiFi assembly and a geneblock (Twist Bioscience). For protein production, the plasmid was transformed to BL21 DE3 cells (NEB #C2527) on LB+Ampicillin (RPI, #A40040), and AMP addition was kept for growth periods. The next morning, colonies were inoculated into a 150 mL culture of LB broth (RPI, #L24065).The culture was grown until the logarithmic growth phase (0.6 OD) and 1L cultures of Auto-induction media (Formedium, #AIMLB0260) in 2L flasks were inoculated at 1%. 6-8 flasks were grown at 18°C at 200-225RPM for 40-48 hours. After this step, all proceeding steps were on ice or at 4°C. Cells were pelleted in 1L bottles (Beckman Coulter #c31597) at 4000RPM for 20 minutes in a JLA-8.1000 rotor. The supernatant was decanted and pellets were resuspended in PBS (Sigma #D8537) and 2 pellets were collected per 50mL conical tubes. The cells were spun at 4000 RPM, the supernatant was discarded, and the pellets were stored at -80°C for later processing.

A protease inhibitor tablet (ThermoFisher #A32963) and 25mL of lysis buffer (20mM HEPES, 500mM NaCl, pH 7.4) were added for each pellet (thawed in an ice bath) and vortexed until smooth. 500µL of antifoam (Sigma #A6426) was added for every 50mL of cell suspension and tubes were inverted to mix. Up to 200mL was added into the Constant Systems Cell Disruptor, set to continuous flow without pump, and a pressure of ∼31kPa. Lysis was collected, weighed to ensure equal weight, and spun at 20,000 xg for 30 minutes. During the spin, 1000uL of TALON bead slurry (Takara #635502) was prepared for every liter of bacteria in a 20 mL gravity column (BioRad #7321010); washing 3x with water, 3x with lysis buffer, and resuspended in lysis buffer, equal to the original volume. After the spin, supernatants were transferred to new falcon tubes and 1mL of prepared bead slurry was added into each tube. Tubes were rocked for 60-90 minutes and then centrifuged at 1000g for 2 minutes. The supernatant was decanted without drying the beads and each pellet was resuspended with 50 mL of lysis buffer. This solution was poured into the column to collect the beads and the following washes were done: additional 50mL Lysis buffer, 50 mL ATP wash (20 mM HEPES pH 7.4, 500 mM NaCl, 10 mM ATP, 10 mM MgCl2), 100 mL Wash 1 (20 mM HEPES pH7.4, 500 mM NaCl, 5mM Imidazole), and 100 mL Wash 2 (20 mM HEPES pH7.4, 150 mM NaCl). 5mL of elution buffer (20 mM HEPES pH7.4, 150 mM NaCl, 200mM Imidazole) was used to resuspend the beads, and the solution was allowed to incubate for 5 minutes. The column was then uncapped, and the eluted protein was collected by gravity. This elution step was repeated, resulting in 10 mL of TALON eluate.

As a second step to this purification, the TALON eluate was used for affinity purification with a Strep column (IBA-LifeSciences #21240001) on a GE AKTAPurifier. The strep column was prepared by washes with 10 column volumes of water then 10 column volumes of Wash 1 at 2mL/min. The TALON eluate was loaded onto the column at 0.5mL/min. Protein from the strep column was eluted into 500µL fractions using 20mL of Wash 2 +2.5mM Desthiobiotin. Samples under the major peak were collected and ran through a 100 kDa concentrator (Milipore Sigma #UFC505024). Protein samples were flash frozen or stored at 4C for immediate use. Protein samples from final and intermediate steps were run on SDS-PAGE with Coomassie staining to assess for purity.

### Lipid binding assay on Native gel

Fluorescently labeled lipids dissolved in chloroform (Avanti #810176C, #810133C, 810297P) were transferred to cleaned glass tubes, evaporated with nitrogen gas, and resuspended in methanol. Nvj3 protein concentration was measured by Bradford Assay (ThermoFisher #23246) and purified MBP was purchased from Abcam (#167888). Purified proteins and lipids were mixed in a 1:20 protein:lipid molar ratio, and allowed to incubate on ice for 2hrs. Samples were mixed with Native Sample Buffer (Bio-Rad #1610738), and loaded on 4-20% TGX Mini-PROTEAN gels under native conditions to run for 90 minutes at 100V. Using the UV channel on the BioRad ChemiDoc Touch Gel Imager, lipids were visualized. The gels were then Coomassie stained for 20min and allowed to de-stain overnight. Coomassie bands were visualized on the BioRad ChemiDoc Touch Gel Imager.

### Generation of stable line for lysosome immunoprecipitation

HEK293T cells (LentiX293T, Takara Bio) were plated to reach 90% confluency for transfection. Cells were cotransfected using Lipofectamine 3000 with 5ug of the transfer vector (pLJM1-TMEM192-RFP-3xHA Addgene#134631 or pLJM1-TMEM192-FLAG Addgene#112742), 3.75ug of psPAX2 packaging plasmid (Addgene 12260), and 1.25ug pMD2.G envelope plasmid (Addgene 12259). The media was replaced after 6 hours and replaced the following morning. 72 hours post-transfection, viral supernatants were collected and purified through a 0.45um low protein absorbance PES filter. The virus was concentrated using the Lenti-X Concentrator at a 1:3 ratio, incubated for 2 hours at 4C, and centrifuged at 1,500xg for 45 minutes at 4°C. The viral pellet was resuspended in complete medium, aliquoted, and stored at -80°C. Target HEK293T cells were seeded in 6 well plates with concentrated virus and 8ug/ml of polybrene in antibiotic-free media. Cells were selected with 2 ug/ml puromycin 24 hours post-transduction and cultured until cell death of WT cells. Expression of the TMEM192-mRFP-3xHA and its localization was validated by immunofluorescence (data not shown).

### Lysosome Immunoprecipitation (Lyso-IP)

HEK293T cells stably expressing TMEM192-RFP-3xHA (Addgene #134631) or TMEM192-FLAG (Addgene #112742) were grown to 80-95% confluency in 15cm dishes (Fisher #FB012925). Dishes were 1) treated with 1mM DMSO for 1 hr, 2) treated with 1mM LLOMe 1hr, or 3) treated with 1mM LLOMe for 1hr and allowed to recover in washout for 4 hrs. After treatment, all following steps occurred on ice or at 4 °C. Cells were harvested using a scraper in 5mL cold DPBS and a transfer pipette (VWR #414004-024) was used to move cells to 15 mL conical tubes. Cells were gently pelleted at 1000xg for 2min. The supernatant was discarded and the pellet was washed once with 1mL of ice cold Lyso-KPBS + 1x Halt PPIC (136 mM KCl, 10mM KH2PO4, 50mM sucrose pH=7.25) and pelleted again at 1000xg for 2min. Cells were resuspended with 1mL of Lyso-KPBS + 1x Halt PPIC (ThermoFisher #78440) and lysed with 25-30 strokes with a 2 mL Dounce homogenizer. Using a pasteur pipette, the homogenate was harvested into low-binding microcentrifuge tubes. An additional 200 µL of ice-cold Lyso-KPBS was added to the homogenizer (Pyrex England, 2mL) and collected with a transfer pipette. Between replicates and experimental conditions, the homogenizer was washed thoroughly with Lyso-KPBS. Lysed cells were centrifuged at 1,000 xg for 5 min, and the post-nuclear supernatants (PNS) were transferred to new tubes. The PNS was spun again at 1,000 xg for 5 min, and transferred to the final PNS tubes. Using a 1mL pipette with a cut tip, α-HA magnetic beads (ThermoFisher #88836, 90µL each sample) were washed 3x with 1 mL Lyso-KPBS buffer and then resuspended in an equal volume of Lyso-KPBS. 90µL of resuspended bead slurry was added to each PNS, and samples were incubated for 20 min with gentle rotation. Beads were separated from the lysate with a magnetic stand (ThermoFisher #MR02), and the flow through was collected for western blot analysis. Continuing to use the magnetic stand, beads were washed twice with 1 mL of ice-cold high salt Lyso-KPBS buffer (150 mM NaCl in KPBS + 1x Halt PPIC). Beads were transferred to a new tube and resuspended with 1 mL ice-cold Lyso-KPBS with inhibitors. For lipidomics analysis, 750µL of the suspension was moved to another tube where the magnetic stand allowed for the final Lyso-KPBS solution to be aspirated and replaced with 400uL of MeOH. For proteomics analysis, 250µL of the suspension was moved to another tube where the magnetic stand allowed for the final Lyso-KPBS solution to be aspirated and replaced with 30µL 0.5% NP-40 in KBPS with inhibitors. Proteomics samples were then rotated gently for 30 min at 4 °C to elute the protein from the beads. Using the magnetic stand, the eluate was collected into the tube. Beads were then resuspended with an additional 10µL 0.5% NP-40 in KBPS with inhibitors and the magnetic stand was used to collect additional eluate, which was combined with the previous.

### Western blotting

Proteins were separated on a precast Mini-PROTEAN TGX 10% SDS-PAGE gel (4561034; BioRad) and then transferred to a 0.45-μm nitrocellulose membrane in SDS transfer buffer (25 mM Tris, 192 mM glycine, 20% methanol, and 0.05% SDS; pH 8.2) using a Criterion tank blotter with plate electrodes (1704070; BioRad) set to 70 V constant for 1hr. Membranes were blocked with 5% milk dissolved in Tris-buffered saline + Tween (TBS-T) buffer, cut with clean scissors, and primary antibodies were allowed to bind overnight at 4°C. Primary antibodies used for determining human lysosome purity are as follows anti-: HA (ThermoFisher #26183), NPC1 (ThermoFisher #PA1-16817), LAMP2(BD Biosciences #555803), CTSD (ThermoFisher #MA532127), Calnexin (Abcam #AB22595), EEA1(BD Biosciences #610456), TOM40(Santa Cruz Biotechnology, #365467), PLN3(Atlas #HPA006427). Primary antibodies used for determining yeast vacuole purity are as follows anti-: Vph1 (received as a gift from Prinz Lab), Pma1 (Abcam #AB4645), Pdi1 (Abcam #AB4644), Por1 (Thermo Fisher #459500), Pet10 (ProSci, custom-made antibody, received from Joel Goodman).

Immunoblots were developed by binding HRP-conjugated anti-rabbit IgG (ab6721;1:5,000; Abcam) or anti-mouse IgG (ab6728; 1:1,000; Abcam) secondary antibodies to the membrane for 1hr in the presence of 5% milk in TBS-T, followed by four washes in TBS-T and developing with ECL substrate (1705061; BioRad). Blot signal was captured using the Chemiluminescence acquisition setting on a BioRad ChemiDoc Touch Gel Imager.

### Proteomics Preparation

For trypsin digestion and label-free MS, 25% of each Lyso-IP sample was loaded into 10% TGX Mini-PROTEAN 50 μL well gels and run 0.5 cm into the gel. Gels were stained with 0.1% Coomassie BB R250 in 10% acetic acid and 50% methanol for 15 min, then destained for 1.5 h in 10% acetic acid and 50% methanol. Destain was then exchanged for milliQ H2O. Protein bands were cut from each lane, diced, and stored at 4°C for delivery to the Proteomics Core Facility at UT Southwestern Medical Center.

Samples were digested overnight with trypsin (Pierce) following reduction and alkylation with DTT and iodoacetamide (Sigma–Aldrich). The samples then underwent solid-phase extraction cleanup with an Oasis HLB plate (Waters) and the resulting samples were injected onto an Orbitrap Fusion Lumos mass spectrometer coupled to an Ultimate 3000 RSLC-Nano liquid chromatography system. Samples were injected onto a 75 μm i.d., 75-cm long EasySpray column (Thermo) and eluted with a gradient from 0 to 28% buffer B over 90 min. Buffer A contained 2% (v/v) ACN and 0.1% formic acid in water, and buffer B contained 80% (v/v) ACN, 10% (v/v) trifluoroethanol, and 0.1% formic acid in water. The mass spectrometer operated in positive ion mode with a source voltage of 2.0 kV and an ion transfer tube temperature of 275°C. MS scans were acquired at 120,000 resolution in the Orbitrap and up to 10 MS/MS spectra were obtained in the ion trap for each full spectrum acquired using higher-energy collisional dissociation (HCD) for ions with charges 2–7. Dynamic exclusion was set for 25 s after an ion was selected for fragmentation.

Raw MS data files were analyzed using Proteome Discoverer v.2.4 SP1 (Thermo), with peptide identification performed using Sequest HT searching against the reviewed human protein databases from UniProt. Fragment and precursor tolerances of 10 ppm and 0.6 Da were specified, and three missed cleavages were allowed. Carbamidomethylation of Cys was set as a fixed modification, with oxidation of Met set as a variable modification. The false-discovery rate (FDR) cutoff was 1% for all peptides. Proteins that were identified with >1 peptide were considered for analysis. If more than one protein in a group had the same score, an equal number of peptide spectrum matches, and an equal number of peptides, the protein with the longest sequence was designated as the master protein. Protein abundances were calculated using the areas under the curve of corresponding peptides.

### Proteomics Analysis

#### Protein abundance and statistical analysis

Analyses used the set of 4,036 proteins supported by at least two total peptides. Two outlier baseline samples were excluded, leaving 25 Lyso-IP samples: three baseline controls, two baseline samples per knockdown, and three samples per genotype after 1 h LLOMe and 4 h after washout. Observed abundances were log2 transformed and median normalized within each sample; missing values were not imputed.

Differential abundance was analyzed using proDA 1.2.0^90^, with siRNA condition (perturbation), experimental state (treatment) and their interaction in a single model. Two-sided contrasts compared time points within each genotype, knockdowns with controls at matched time points, and changes between time points across genotypes. Response contrasts compared the change from each genotype’s own starting time point. Benjamini–Hochberg (BH) correction was applied across all 4,036 proteins separately per contrast; significance was defined as FDR < 0.05 without a minimum fold-change threshold. Mock-IP samples were modeled separately to assess nonspecific recovery.

#### Lyso IP specificity and global sample structure

Specificity was assessed against a published reference of 213 core lysosomal proteins^48^. For proteins detected in at least two baseline control Lyso-IP replicates and both baseline mock replicates, enrichment scores were calculated as the difference between group medians of sample-median-centered log2 abundances. Core and other proteins were compared by a one-sided Wilcoxon rank-sum test. Principal component analysis used the 2,209 proteins observed in all 25 retained samples, centered by protein without variance scaling. Pearson and Spearman correlations summarized concordance between fitted knockdown effects descriptively.

#### Gene Ontology enrichment

Gene Ontology (GO) cellular component and biological process enrichment used human UniProt-GOA annotations archived in July 2026, including annotations propagated to parent terms. Overrepresentation was tested by one-sided hypergeometric tests against the 4,036-protein analysis background, with BH correction and FDR < 0.05. Control LLOMe sets were divided into significant increases and decreases. Concordant basal and LLOMe-endpoint sets required significance in at least one knockdown and matching effect signs in both. Shared post-washout sets required significance in both knockdowns and matching signs. Differences between control LLOMe and basal knockdown effects required significant direct comparisons with both knockdowns and matching difference signs.

For each biological comparison, BH correction was applied jointly across the two directional protein sets, separately for CC and BP. All eligible tested terms entered correction before display filtering. Bubble plots show observed/expected enrichment, GO FDR and contributor count, displaying terms with FDR < 0.05, at least three contributors and at most 200 background members, except for “lipid metabolic process” in the LLOMe-endpoint comparison, which had 304 background members.

#### Rank Enrichment for the LLOMe response

Proteins were ranked by basal knockdown/control log2 fold change, and the control LLOMe-increased and LLOMe-decreased sets were evaluated using unweighted directional enrichment scores from cumulative hit-minus-nonhit curves. Significance was assessed against a fitted simulation null that removed alignment between LLOMe and knockdown effects while retaining the paired knockdown structure, shared-control covariance and data-estimated correlated measurement noise. LLOMe-response sets were reselected in each simulation. Empirical calibration used 10,000 null datasets with latent-model refitting and 499 inner simulations per dataset, with the same simulation resolution for the observed-data probabilities. Calibrated P values were Holm-adjusted across the four directional tests and the procedure was checked in 2000 independent null datasets.

#### Trajectory classification and shared changes

Proteins detected in at least two replicates at each stage were classified independently in each genotype using the direction and significance of the baseline-to-LLOMe, LLOMe-to-washout and baseline-to-washout contrasts. Among 2,969 eligible control proteins, six main patterns distinguished acute increases or decreases followed by reversal, acute changes without detected reversal, and later increases or decreases without a detected acute response. Reversal required a significant opposite-direction change after washout, not return to baseline.

We compared protein trajectory assignments between control and knockdown cells, including only proteins meeting the classification criteria in both conditions. For proteins assigned to different patterns, we assessed whether direct statistical comparisons supported the reassignment. This required a significant difference in the acute response (LLOMe minus baseline) or the washout response (post-washout minus LLOMe), with FDR < 0.05 and a direction consistent with the reassignment.

To identify response changes shared by both knockdowns, we selected proteins whose response differed from control in the same direction in SNX13-KD and SNX14-KD, with FDR < 0.05 in at least one knockdown. Proteins could meet this criterion even if their trajectory assignment remained unchanged.

For visualization, each protein’s control baseline was set to zero, and all other measurements were expressed relative to that value. This preserved differences already present in knockdown cells before LLOMe treatment.

GO enrichment of control patterns and altered memberships used control-eligible or mutually eligible proteins as background, with BH correction across the full trajectory enrichment group. Representative significant control terms were selected for interpretation without recalculating their FDRs. GO annotations in the shared P1 response subset were tested for over- or underrepresentation using two-sided Fisher exact tests, with the 301 proteins eligible for P1 comparison across control and both knockdowns as background. BH correction was applied across 12 predefined P1 GO terms.

#### Lysosomal damage sensors and response proteins

Proteins implicated in lysosomal damage sensing and downstream damage responses were manually curated from published literature. Existing fitted abundance estimates and trajectory assignments were displayed relative to each protein’s control baseline in a heatmap, without row scaling or additional statistical testing.

#### Software and packages used

R: 4.0.2 and 4.6.1. R packages include: proDA (1.2.0), readxl (1.4.0), ggplot2 (3.3.2), ggrepel (0.8.2), pheatmap (1.0.12), gridExtra (2.3), patchwork (1.0.1) and viridisLite (0.4.0). Python: 3.9.6 and 3.12.14. Python packages include: NumPy (2.3.5), pandas (2.2.3), SciPy (1.18.1), openpyxl (3.1.5), ReportLab (4.4.9), pypdf (6.10.0).

### Statistical Analysis

#### General

Unless otherwise stated, statistical analyses were performed using GraphPad Prism 11 (GraphPad Software, Inc.) or Python (for behavior analyses). When possible, data was analyzed blind to the genotype. Sample size for each experiment was determined based on similar studies. To compare the means of groups where normal distribution and similar variance between groups was confirmed, two-tailed Student’s *t*-test (for two samples), one-way ANOVA (for more than two samples) or two-way ANOVA followed by Sidak’s or Tukey’s post hoc test (for multiple variables) was used. A *P* value less than 0.05 was considered statistically significant.

Behavioral and Purkinje cell histology data (Fig 2 and Fig S2) were analyzed in Python (version 3.13.15) on Google Colaboratory using statsmodels (version 0.15.0)^91^, SciPy (version 1.16.3)^92^, pandas (version 2.2.3), NumPy (version 2.1.3) and matplotlib (version 3.10.0)^93^. Both sexes were included in all analyses, and sex was treated as a biological variable. Because mice were tested in six cohorts, cohort was included as a random effect in all linear mixed models.

#### Mouse Behavior specific

Measures obtained once per mouse and treated as continuous (composite ataxia score, and PC density and molecular layer thickness in each lobule) were analyzed with linear mixed models that included genotype, sex and their interaction as fixed effects and cohort as a random intercept. Models were fitted by restricted maximum likelihood with sum-to-zero contrasts, and fixed effects were tested with Wald χ² tests (Type III). Repeated measures were analyzed at the level of individual trials. Rotarod latency to fall was modeled with genotype, sex, trial (nine trials) and their interactions as fixed effects, and vertical pole T-turn and descending times with genotype, sex, day and their interactions as fixed effects; both models included random intercepts for mouse and cohort. Pole test times were right-skewed and were log-transformed, log(1 + x), before analysis; data are presented on the original scale. PC density and molecular layer thickness across lobules II, III and X were analyzed on section-level data with genotype, lobule and their interaction as fixed effects and random intercepts for mouse and cohort; values plotted for each mouse are the mean of two sections per lobule. Ordinal scores (ledge test, hindlimb clasping, gait, kyphosis and turn quality) were compared between genotypes with two-sided Mann-Whitney U tests, with sexes pooled and within each sex.

Post hoc comparisons of Snx13PCKO and Control mice (with sexes pooled, within each sex, on each day, or within each lobule) were performed only when the genotype main effect or an interaction involving genotype was significant. They were estimated as linear contrasts from linear mixed models fitted separately to each day (pole test, log-transformed daily means), each trial (rotarod) or each lobule (histology), with genotype, sex and their interaction as fixed effects and cohort as a random intercept, and p values were Bonferroni-corrected for the number of comparisons within each panel. For ordinal scores, within-sex Mann-Whitney U tests were performed only when the pooled test was significant and were Bonferroni-corrected for the two comparisons. The relationship between PC density in lobule II and behavioral scores was assessed in mice with both measures by Pearson correlation and least-squares linear regression, with Spearman rank correlation as a nonparametric check; to test whether PC density predicted behavior beyond the effect of genotype, genotype was added as a covariate to the regression model. For these data, statistical significance was set at p<0.05, and 0.05≤p<0.10 is indicated as a trend (#). Box plots show the median, interquartile range and minimum to maximum values, with each mouse plotted; line graphs show mean ± S.E.M. Sample sizes and statistical results for each panel are given in the figure legends.

## Supplementary Figures

**Figure S1.**
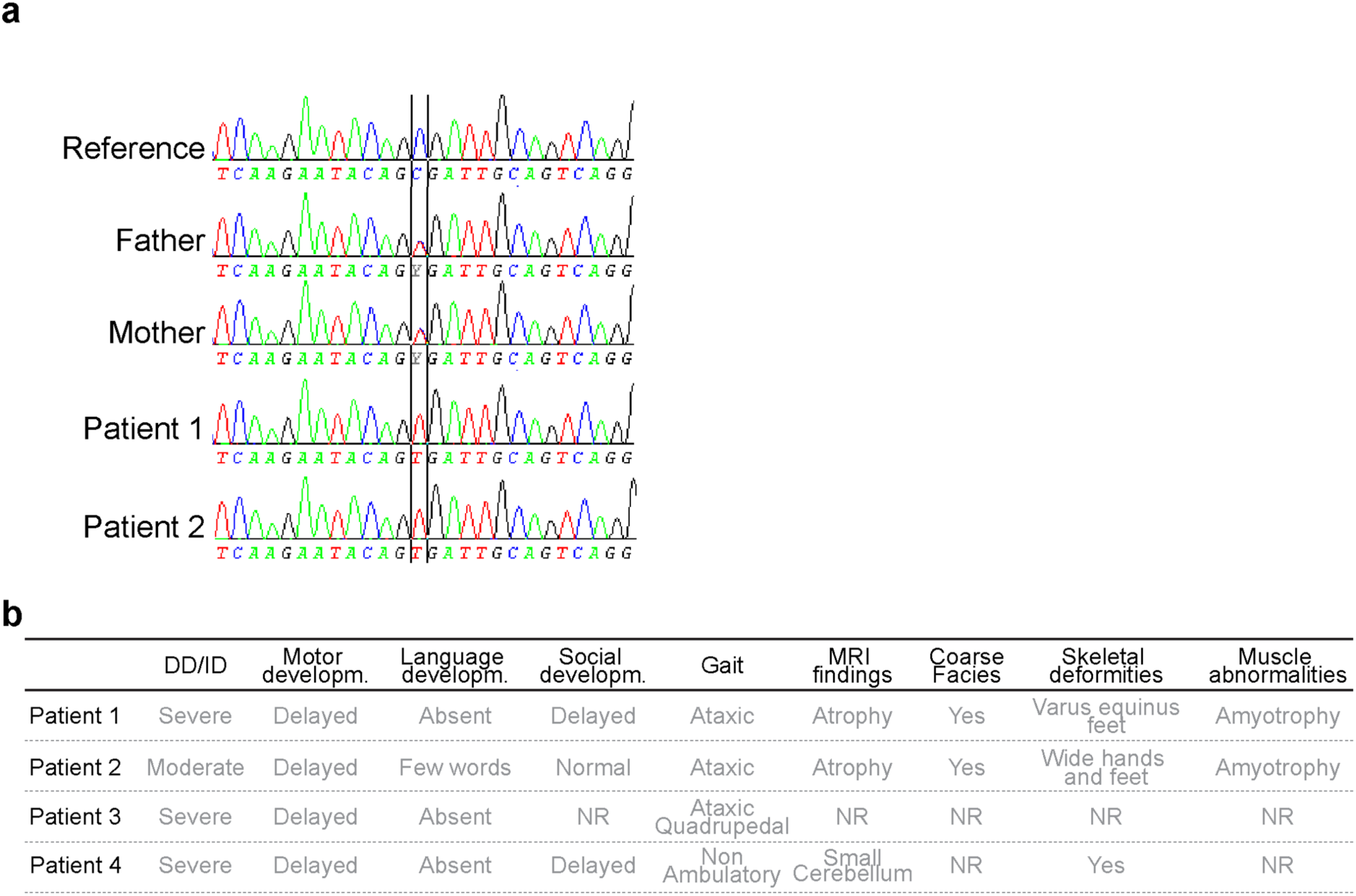
Chromatogram and photo of individual carrying homozygous *SNX13* nonsense variant. **a.** Sanger sequencing confirms that patients P1 and P2 are homozygous for the *SNX13* c.1048C>T variant and parents heterozygous. A sequence from an unrelated individual is shown as reference in the top. **b.** Table with major clinical features of patients with homozygous SNX13 loss of function variants. DD; developmental delay; ID: intellectual disability; NR: not reported.

**Extended data Fig. 2.**
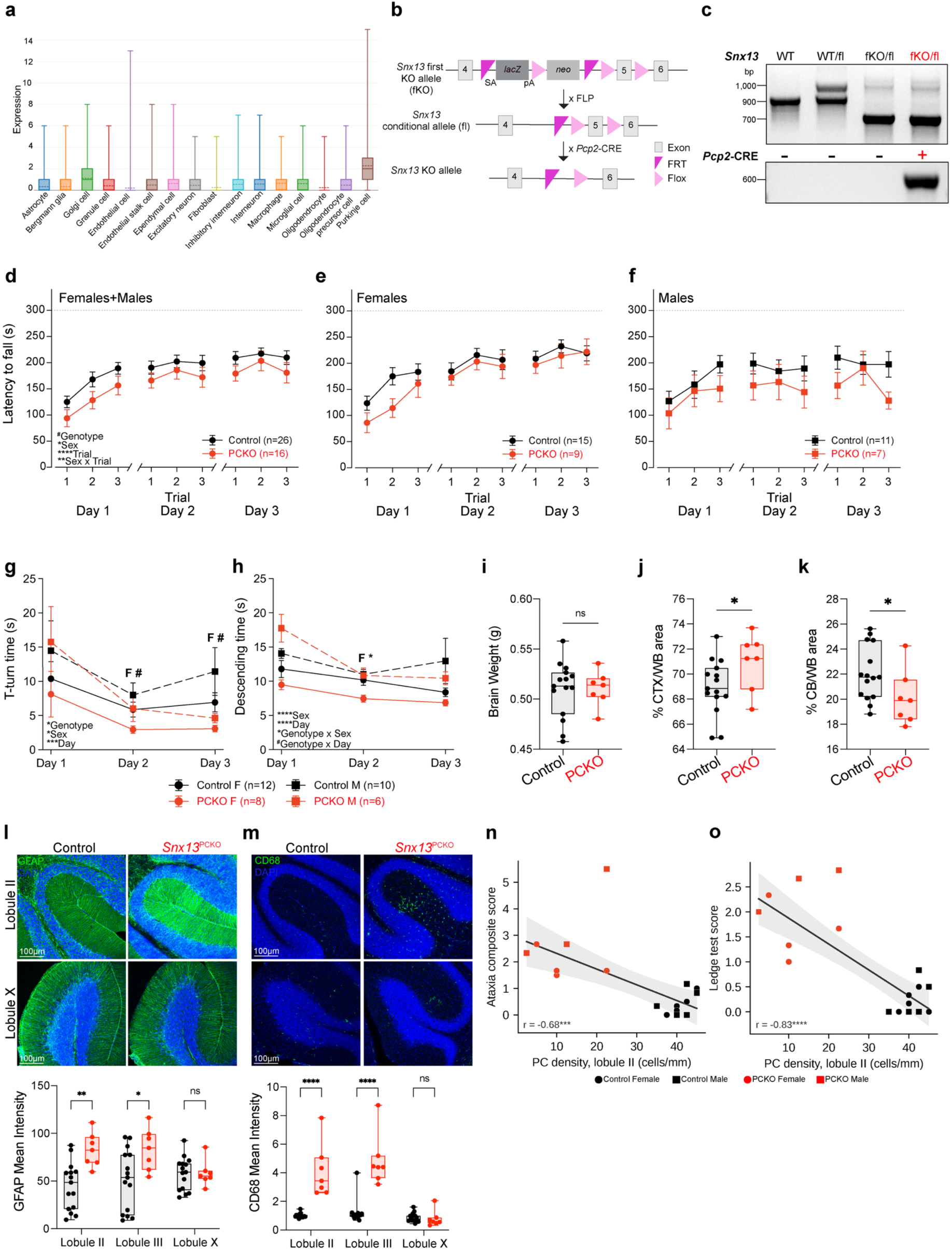
*Snx13* is expressed in Purkinje Cells of the cerebellum and its specific deletion in *Snx13^PCKO^* leads to signs of neurodegeneration in mice. **a.** *Snx13* expression levels in the adult mouse cerebellum from single nuclei RNAseq deposited in Broad Institute Single Cell Portal. **b.** Schematic of *Snx13* alleles used to generate conditional Purkinje cell (PC) specific *Snx13* KO mice (*Snx13*^PCKO^). **c.** Representative image of agarose gel from PCR genotyping to identify Control (*Snx13*^flox/+^; Pcp2-Cre or *Snx13*^flox/-^) from *Snx13*^PCKO^ mice. **d-f.** Quantification of latency to fall on the accelerating rotarod on three trials per day over three consecutive days, for all mice (D; n=26 control, n=16 *Snx13*^PCKO^), females (E; n=15 control, n=9 *Snx13*^PCKO^) and males (F; n=11 control, n=7 *Snx13*^PCKO^); dotted line, 300 s cutoff. Statistical analysis was performed using omnibus fixed effects with Wald χ^2^ tests in linear mixed models (genotype, lobule and their interaction fixed; cohort random intercept; animal added for repeated measures). Line plots show mean ± S.E.M. **g-h.** Quantification of t-turn time (G) and descending time (H) on the vertical pole in n=12 female and n=10 male control mice and n=8 female and n=6 male *Snx13*^PCKO^ mice. Statistical analysis was performed using omnibus fixed effects with Wald χ^2^ tests in linear mixed models (genotype, lobule and their interaction fixed; cohort random intercept; animal added for repeated measures). Line plots show mean ± S.E.M. **i-k.** Quantification of brain weight (I), percentage of cerebral cortex (CTX) (J) and cerebellum (CB) (K) relative to whole brain area (WB) of 4-month-old mice. Statistical analysis was performed with unpaired, two-sided Student’s *t*-test. Box plots show median and interquartile range and whiskers minimum and maximum values. **l-m.** Representative images and mean intensity quantifications of cerebellar lobule II, III, and X immunostained with the astrocytes marker anti-GFAP (l) and microglia marker anti-CD68 (m) antibody. Box plots show median and interquartile range and whiskers minimum and maximum values of n=15 control (7 females, 8 males) and n=7 *Snx13*^PCKO^ mice (3 females, 4 males). Two-way ANOVA followed by Tukey’s test. Scale bar: 100 μm. **n-o.** Correlation between PC linear density in lobule II and composite ataxia score (n) or ledge test score (o) in mice with both measures (n=11 Control, n=7 *Snx13*^PCKO^). Lines show least-squares fits across all mice with 95% confidence bands. Pearson correlation: n, r=-0.68; o, r=-0.83 with genotype included as a covariate. Because PC density and ataxia severity separate largely by genotype, and the number of Snx13PCKO mice with histology was small, these correlations should not be read as evidence that the extent of PC loss predicts ataxia severity within mutants. Statistical significance is noted as: non significant are ns or not shown, # = near significant, * p<0.05, ** p<0.01, *** p<0.001, and **** p<0.0001. Additional statistics are available in Table S2.

**Extended data Fig. 3.**
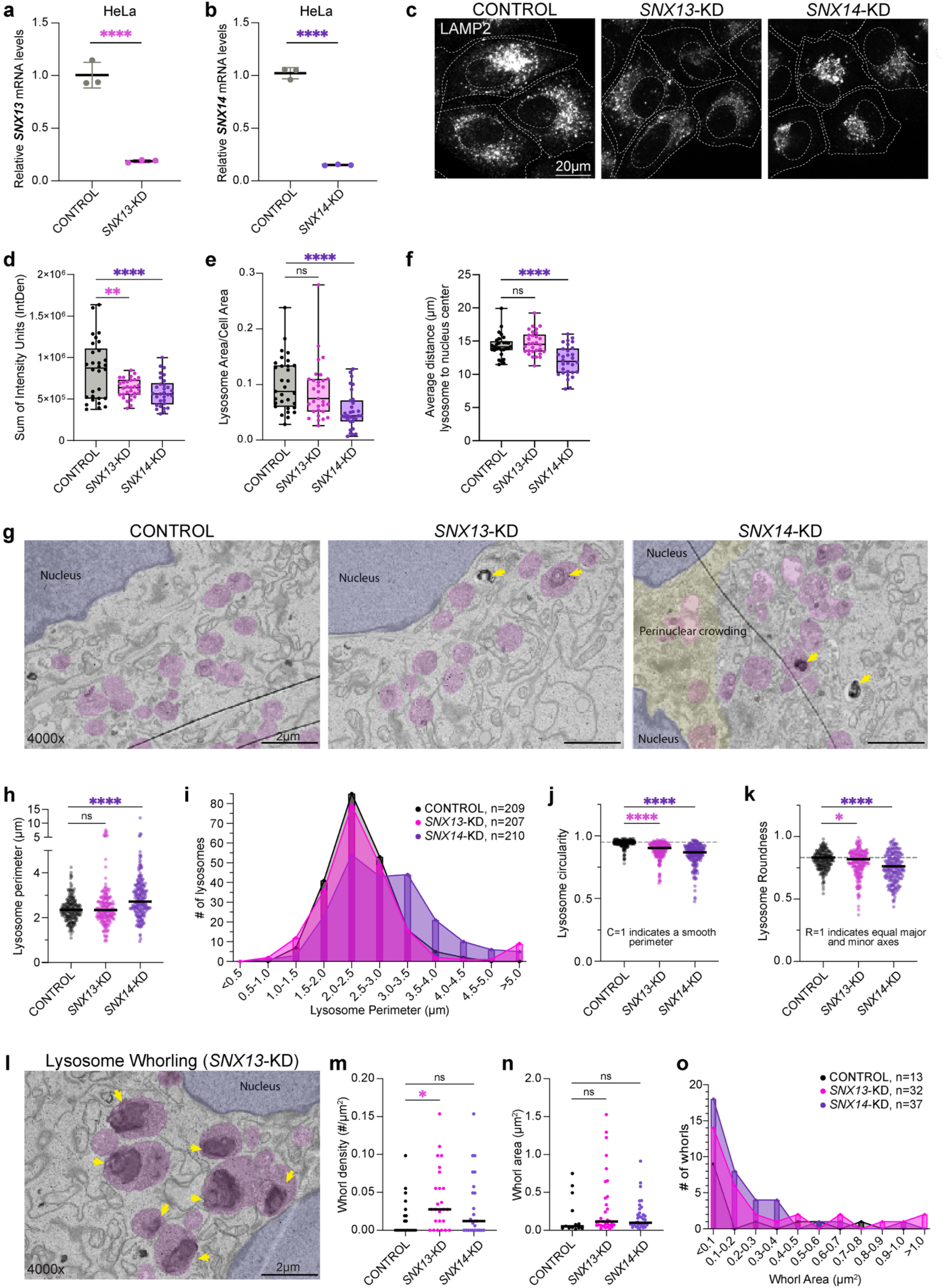
SNX13 and SNX14 siRNAs reduce their expression in HeLa cells and alter lysosomes. a, **b.** Relative *SNX13* (A) and *SNX14* (B) mRNA expression measured by qPCR from HeLa cells after 48h siRNA knockdown. n=3 averages of triplicates, analyzed by student’s t-test. **c.** Representative images of LAMP2 staining in fixed HeLa cells after siRNA knockdown. Cell boundaries are marked in dotted lines. Scale bar: 20µm. **d.** Integrated density of LAMP2 signal per cell. Value distribution shown by min-to-max box plot of n=30 cells from 3 independent experiments, analyzed by one-way ANOVA followed by Dunnett’s test for correction of multiple comparisons. **e.** Fraction of cell area taken by LAMP2 area. Lower fraction of area is consistent with clustering. Value distribution shown by min-to-max box plot of n=30 cells from 3 independent experiments, analyzed by one-way ANOVA followed by Dunnettt’s test for correction of multiple comparisons. **f.** Average distance (µm) of lysosomes from the nucleus center per cell to quantify peri-nuclear lysosome clustering. Value distribution shown by min-to-max box plot in n=30 cells from three independent experiments and analyzed by one-way ANOVA followed by Dunnett’s test for correction of multiple comparisons. **g.** Representative electron micrograph images of lysosomes (pink), relative to the nucleus (blue) at 4000x, including perinuclear crowding seen in SNX14 KD cells (yellow). Yellow arrows denote electron dense materials or whorling. Scale bar: 2µm. **h.** Perimeter of single lysosomes manually identified in TEM. Black line denotes mean of n=207-210 lysosomes from at least ten cells analyzed by one-way ANOVA followed by Dunnettt’s test for correction of multiple comparisons. **i.** Frequency distribution of lysosomes identified in panel H, binned by perimeter with size set to 0.1µm. **j, k.** Measures of single lysosomes identified in panel H, including circularity (J, smooth perimeter) and roundness (K, equal major and minor axes). Black line denotes mean of n=207-210 lysosomes from at least 10 cells, analyzed by one-way ANOVA followed by Dunnett’s test for correction of multiple comparisons. **l.** Representative electron micrograph image of lysosome whorling denoted by yellow arrows, in a *SNX13-*KD cell at 4000x. Includes lysosomes (pink) and the nucleus (blue). Scale bar: 2µm. **m.** Whorl density, calculated as the number of whorls (defined as 3+ closely encased layers) per unit area from TEM images. Black line denotes mean of n=25 images from at least 10 cells, analyzed by one-way ANOVA followed by Dunnettt’s test for correction of multiple comparisons. **n.** Area of single whorls manually identified in TEM. Black line denotes mean of n=13-37 from at least 15 cells, analyzed by one-way ANOVA followed by Dunnettt’s test for correction of multiple comparisons. **o.** Frequency distribution of whorls identified in panel N, binned by individual area with size set to 0.25µm^2^. ns = nonsignificant, *P<0.05, ***P<.001, and **** p<0.0001.

**Extended data Fig. 4.**
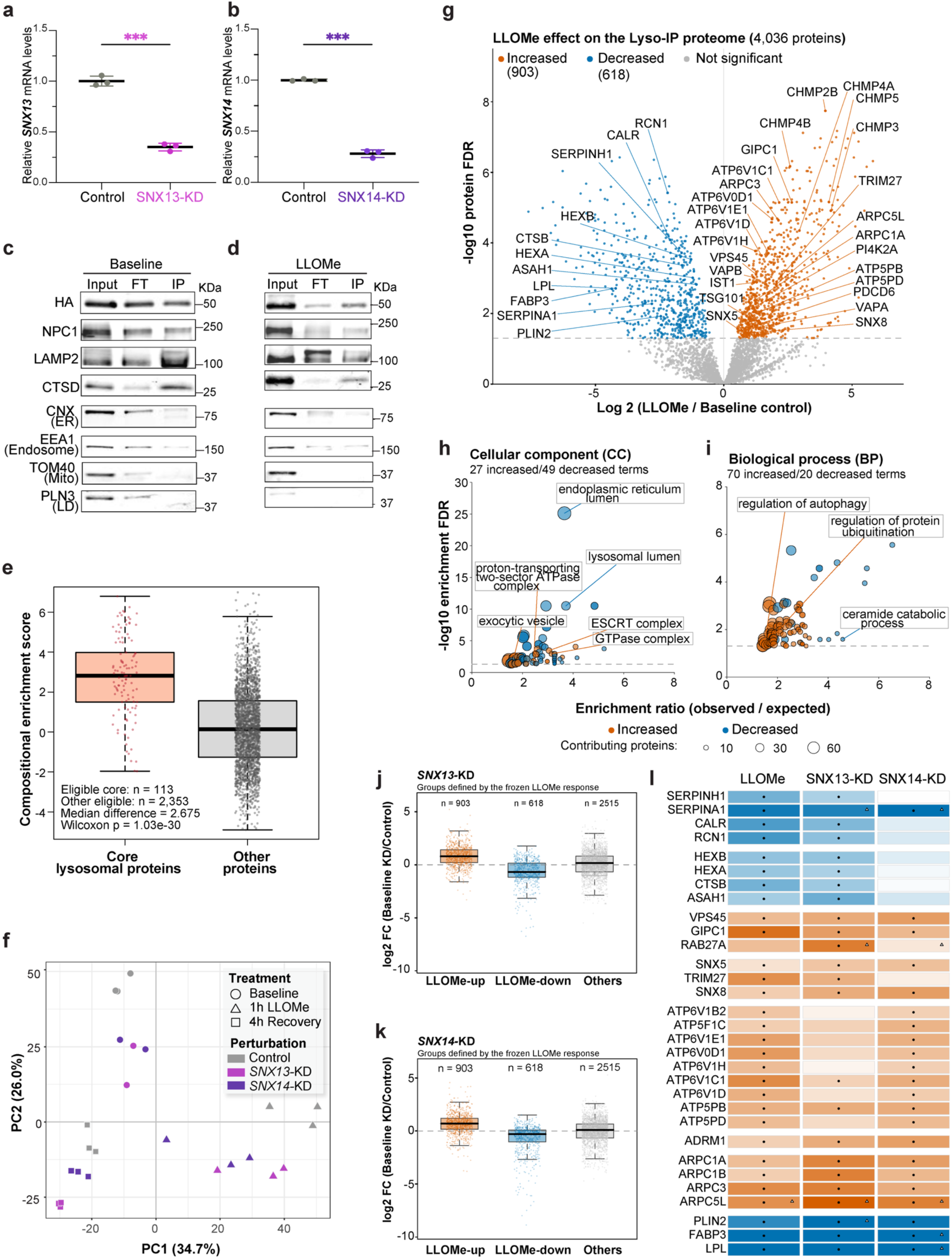
Lysosome-IP enriches for lysosomal proteins at baseline and reveals an altered proteome after LLOMe induced damage and in *SNX13-*KD or *SNX14-*KD cells. a, **b.** Relative *SNX13* (a) and *SNX14* (b) mRNA expression as measured by qPCR from HEK293T cells expossed to siRNAs for 48h. **c, d.** Western blot analysis of input, flow-through (FT), and lyso-IP fractions form HEK293T cells under baseline (c) or LLOMe treatment (d). HA detects tagged TMEM192; NPC1, LAMP2 and CTSD are lysosomal markers. Calnexin (CNX), EEA1, TOM40 and PLN3 assess endoplasmic-reticulum, endosomal, mitochondrial and lipid-droplet material, respectively. Molecular masses are indicated in kDa. **e.** Lyso-IP specificity assessed by comparing enrichment scores for 113 core lysosomal proteins and 2,353 other proteins. Scores represent normalized log₂ abundance differences between baseline control Lyso-IP and mock-IP samples (see Methods). Significance was assessed by a one-sided Wilcoxon rank-sum test (P = 1.03 × 10⁻³⁰). Points represent proteins; boxes show medians and interquartile ranges, with whiskers extending to the most extreme values within 1.5 interquartile ranges. **f.** Principal component analysis of 25 Lyso-IP samples using 2,209 proteins quantified in every sample. Points represent biological replicates; color indicates perturbation and shape indicates baseline, 1 h LLOMe or 4 h after washout. PC1 and PC2 explain 34.7% and 26.0% of the variance, respectively. **g.** Volcano plot of control-cell protein abundance changes after 1 h LLOMe relative to baseline. Orange and blue indicate significantly increased (903) and decreased (618) proteins, respectively; gray indicates the remaining proteins. The dashed line marks FDR = 0.05. Selected proteins are labelled. **h, i.** Bubble plots of GO cellular component (h) and biological process (i) enrichment of LLOMe-increased (orange) and decreased (blue) proteins. Bubble positions indicate enrichment ratio (observed/expected) and -log10 FDR; bubble area represents the number of contributing proteins. Enrichment was assessed using hypergeometric tests with Benjamini–Hochberg correction (see Methods). Dashed lines mark FDR = 0.05. **j, k.** Basal SNX13-KD/control (j) and SNX14-KD/control (k) log2 fold changes, grouped by the LLOMe response in control cells: increased, decreased or not significantly altered. Points represent proteins; box and whisker definitions are as in (e). **l.** Heatmap of log₂ fold changes for 31 selected proteins after LLOMe in control cells and at baseline in each knockdown, all relative to Control baseline. Colors are capped at ±3. Black dots indicate FDR < 0.05; open triangles indicate fewer than two detected replicates in Control baseline or the corresponding comparison group. Protein-level significance was assessed using proDA with Benjamini–Hochberg correction (see Methods).

**Extended data Fig. 5.**
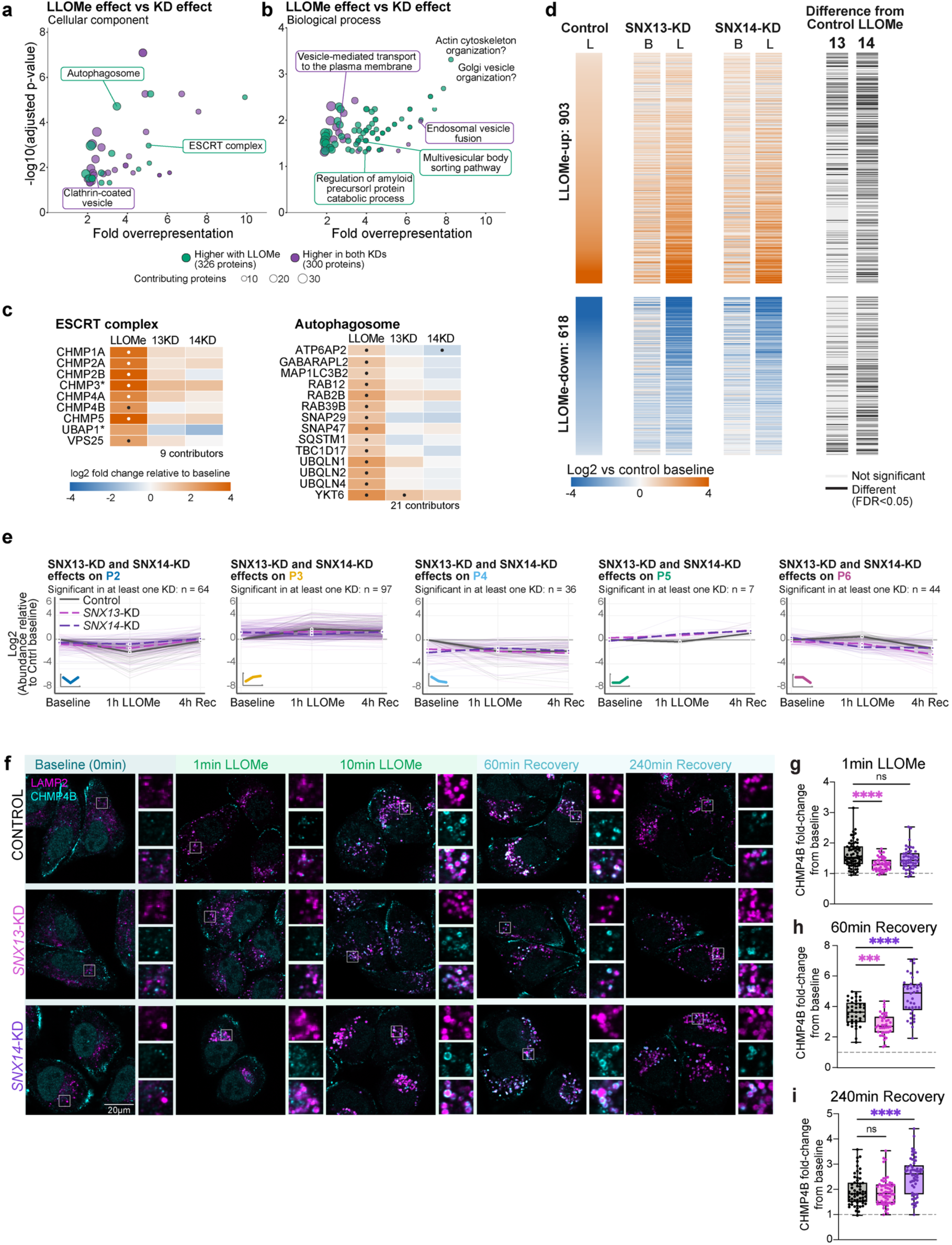
Isolation of lysosomes after damage and recovery reveal different patterns of protein recruitment/depletion after loss of SNX13 or SNX14. a, **b.** GO cellular component (a) and biological process (b) enrichment of proteins whose control LLOMe/baseline effects differ significantly from both basal KD/control effects (FDR < 0.05 in both direct comparisons). Teal indicates higher effects with LLOMe (326 proteins), and purple indicates higher effects in both basal knockdowns (300 proteins). Bubble positions indicate enrichment ratio (observed/expected) and −log₁₀ FDR; area represents the number of contributing proteins. Enrichment was assessed using hypergeometric tests with Benjamini–Hochberg correction (see Methods). **c.** Heatmaps of proteins contributing to the ESCRT-complex and autophagosome enrichments in A. Columns show control LLOMe and basal SNX13-KD and SNX14-KD effects, all relative to control baseline. Colors represent log₂ fold changes, capped at ±4. All proteins meet the direct-comparison criteria in A/B; dots indicate significance of the individual effects relative to control baseline (FDR < 0.05), with black or white used for visibility. Asterisks indicate fewer than two detected replicates in at least one relevant condition. Seven autophagosome contributors also belong to ESCRT and are displayed only in the ESCRT heatmap. **d.** Heatmap of the 903 LLOMe-increased and 618 LLOMe-decreased proteins across control LLOMe and both knockdowns at baseline (B) and after LLOMe (L). Values show log₂ abundance differences relative to control baseline. Black marks in the adjacent strips indicate significant differences between LLOMe-treated knockdown and LLOMe-treated control cells (FDR < 0.05); light marks indicate no significant difference. **e.** Trajectories of proteins from control patterns P2–P6 with acute-response differences in the same direction in both knockdowns and significance in at least one (FDR < 0.05), selected as in Fig. 5F. The most frequent direction within each pattern is shown: weaker acute decreases for P2/P4, lower acute responses for P3/P6, and higher acute responses for P5. Thin lines represent individual proteins and thick lines their medians in control (gray), SNX13-KD (magenta) and SNX14-KD (purple), relative to each protein’s control baseline. **f.** Representative images of HeLa cells with staining of lysosomes (LAMP2, magenta), and ESCRT-III component (CHMP4B, cyan), over experimental time. 60min and 240min recovery is measured after 10 minutes of 1mM LLOMe treatment. Insets show CHMP4B localization to the lysosome surface. **g-i.** Fold-change in CHMP4B signal on lysosomes per cell from each condition’s baseline to 1 min LLOMe (g), 60min recovery (h), and 240min recovery (i) from n=75 cells for control and n=50 cells for KDs from 3 and 2 independent experiments, respectively. Statistical analysis by one-way ANOVA followed by Dunnettt’s test for correction of multiple comparisons.

**Extended data Fig. 6.**
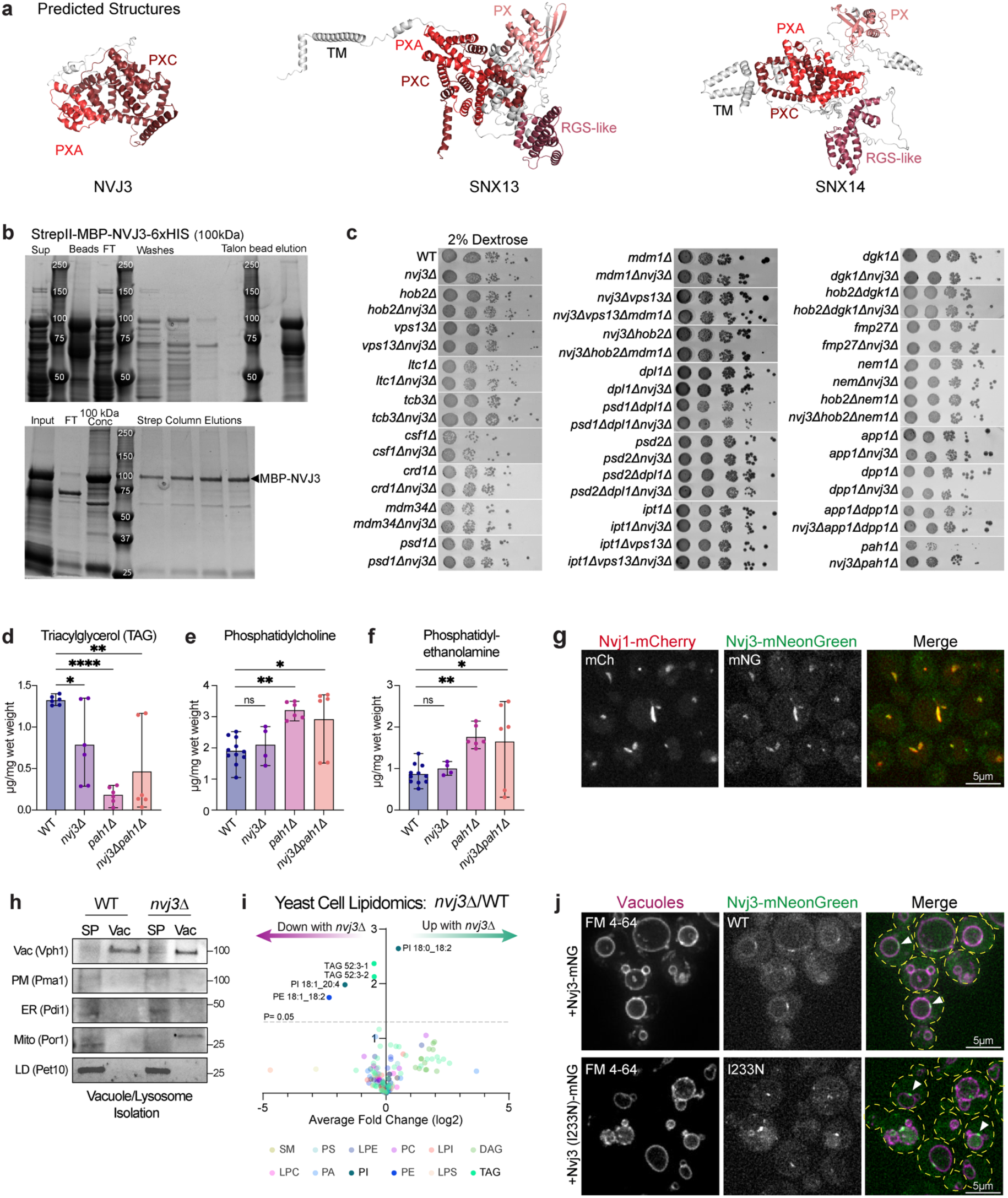
Nvj3 localizes at the NVJ and specifically modulates PA metabolism in a genetic screen. **a.** Predicted structures of NVJ3, SNX13, and SNX14 proteins, with color-coded and labeled domains, informed by AlphaFold3 and partial crystal structures. **b.** Coomassie staining of native acrylamide gels with washes and fractions of Nvj3 purification from BL21 DE3, using TALON and Strep column affinity. MBP-Nvj3 band is at 100 kDa. **c.** Genetic screen, testing interactions between Nvj3 and genes associated with lipid metabolism and other organelle contact sites, showing serial dilutions of labeled yeast strains grown on synthetic dextrose at 30C. **d-f.** Quantification of lipids from thin layer chromatography (TLC) of whole yeast cells (μg/mg of cell wet weight), using standards, in n=4-12 TLC analyses from 6 wild type and 3 knockout yeast cultures. Statistical analysis was performed with one-way ANOVA followed by Dunnett’s test for correction of multiple comparisons. **g.** Representative image of yeast with endogenously tagged Nvj1-mCherry (marks the NVJ, in red) and Nvj3-mNeonGreen (green), showing co-localization. Scale bar: 5 µm. **h.** Western blot analysis of spheroplasts (SP, yeast without cell wall) and purified vacuole fraction from wildtype and Nvj3-KO yeast, showing protein markers for intact yeast lysosomes and other organelle compartments. Blots were developed with ECL. **i.** Volcano plot of lipid species from whole cell yeast, showing fold-changes in the Nvj3-KO vs wildtype. Lipid species are color-coded by major lipid groups, consistent with the legend. The dotted line represents p<0.05, analyzed by two-tailed t-test from n=3 independent lipid extracts. **j.** Representative images of yeast transformed with Nvj3-mNeonGreen (top) or Nvj3(I233N)-mNeonGreen (bottom), showing localizations consistent with NVJ (white arrows). Yeast lysosomes are shown in magenta by FM4-64 dye, and Nvj3 is shown in green. Scale bar: 5 µm. Statistical significance is noted as: ns = nonsignificant, * p<0.05, *** p<0.001, and ****p<0.0001.

**Extended data Fig. 7.**
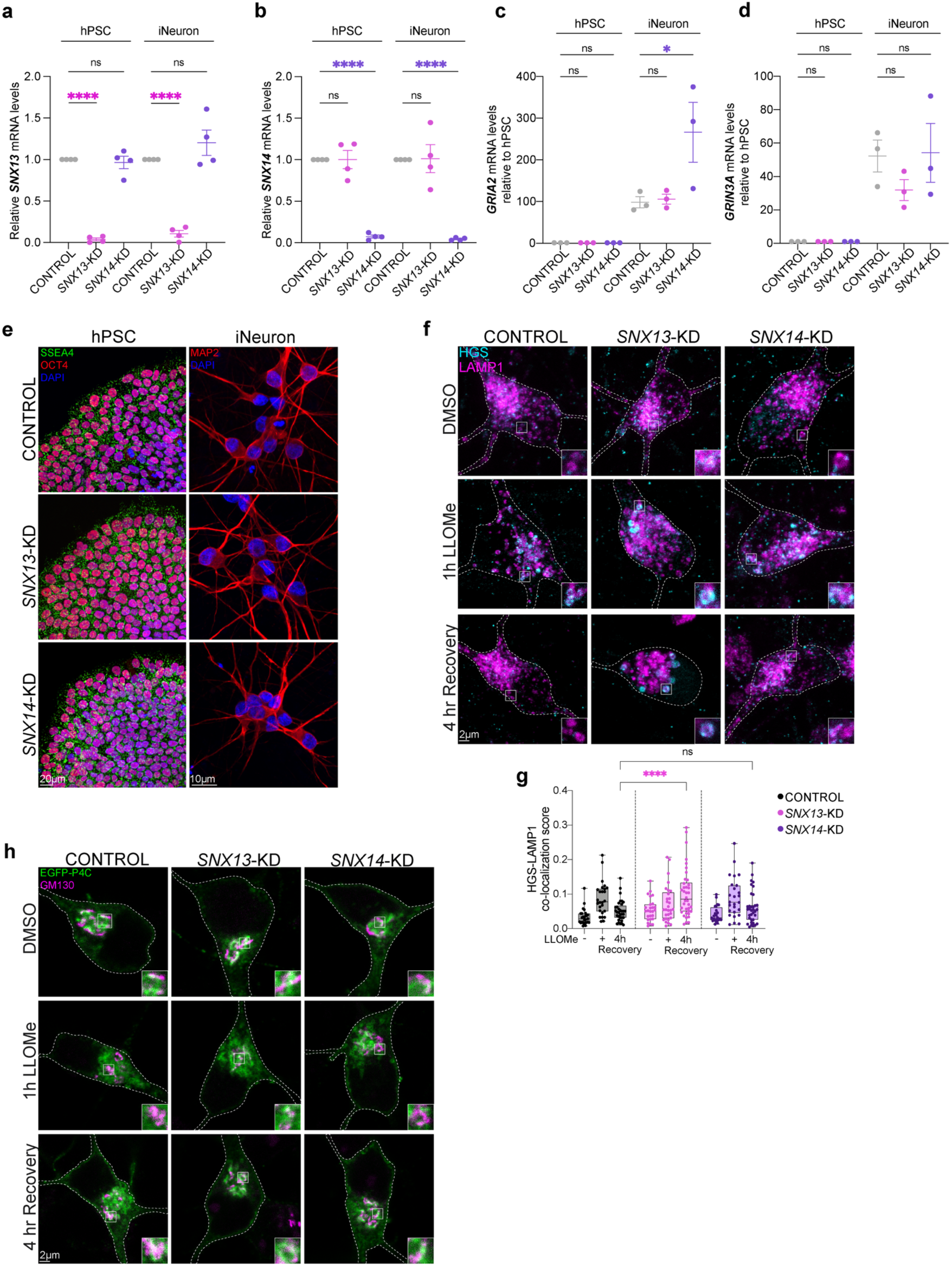
Knockdown of *SNX13* or *SNX14* in hPSCs does not affect differentiation to neurons that show signs of lysosome membrane damage. a, **b.** Quantification of relative *SNX13* (a) or *SNX14* (b) mRNA levels from hPSCs and D14 neurons. Graphs show mean ±S.E.M of n=4 independent hPSC cultures and differentiations. One-way ANOVA followed by Tukey’s test for correction of multiple comparisons. **c, d.** Quantification of relative *GRIA2* (c) and *GRIN3A* (d) expression levels quantified by qPCR in hPSCs and D14 neurons. Graphs show mean ±S.E.M of n=3 independent hPSC cultures and differentiations. One-way ANOVA followed by Tukey’s test for correction of multiple comparisons. **e**. Representative images of hPSCs stained with anti-SSEA4 and anti-OCT4 and neurons stained with anti-MAP2 at D14 in Control, *SNX13*-KD, and *SNX14*-KD. Scale bars: 20 μm and 10 μm. **f.** Representative images of anti-HGS and anti-LAMP1 in Control, *SNX13*-KD, and *SNX14*-KD D21 neurons treated with DMSO or 0.5 mM LLOMe for 1h and 4h washout for recovery. Scale bar: 2 μm. **g.** Quantification of co-localization ratio between HGS and LAMP1. Box plots show the median and interquartile range with whiskers from minimum to maximum of n=24-35 neurons from 3 independent differentiations. Two-way ANOVA followed by Dunnett’s test for multiple comparisons. **h.** Representative images of EGFP-P4C-SidC biosensor (green, localized in the trans-Golgi at baseline conditions) and anti-GM130 immunostaining (purple, cis-Golgi marker) in Control, *SNX13*-KD, and *SNX14*-KD D21 neurons treated with DMSO or 0.5 mM LLOMe for 1h and 4h washout for recovery. Statistical significance is noted as: ns = nonsignificant, * p<0.05 and ****p<0.0001.

